# Registration and reporting characteristics of trials investigating exercise therapy following total knee arthroplasty: A systematic review

**DOI:** 10.1101/2025.10.07.25337493

**Authors:** Birk M Grønfeldt, Rasmus S Husted, Rasmus H Brødsgaard, Line Holst, Thomas Kallemose, Cathy M Chapple, Carsten B Juhl, Thomas Bandholm

## Abstract

**Objectives:** Prospectively registering the primary trial outcome is important to reduce selective outcome reporting and increase trustworthiness of findings used to guide clinical practice. The objective of this systematic review was to explore and compare the reporting characteristics of prospectively and non-prospectively registered trials investigating exercise therapy following total knee arthroplasty.

**Design:** Trials comparing effects of exercise therapy after total knee arthroplasty due to osteoarthritis were sought in four databases from 2000 (clinicaltrials.gov launch) to 12^th^ of August 2024. Randomised controlled trials comparing different exercise therapy interventions were included. Primary outcomes were extracted using a pre-specified hierarchy to reflect each trial’s most consistently reported outcome. Risk-of-bias was assessed using the Cochrane Collaborations Risk-of-Bias tool version 2.

**Results:** Ninety-four trials (n = 9,396) were included: 13 prospectively registered, 33 retrospectively registered, and 48 unregistered. A single primary outcome was defined in 43.6% of trials. Four trials reported a primary outcome consistent with a prospective registration. Prospectively registered trials reported smaller effect estimates (SMD 0.06; 95% CI −0.03 to 0.16) than retrospectively registered (SMD 0.67; 95% CI 0.22 to 1.11) and unregistered trials (SMD 0.59; 95% CI 0.32 to 0.86), and more often defined a single primary outcome, reported sample size calculations, and followed intention-to-treat principles.

**Conclusion:** Prospective registration and clear primary outcome definition were uncommon in trials comparing exercise therapy after total knee arthroplasty. Trials without these elements had larger effect size estimates at higher risk of bias, suggesting that registration status is an important indicator of methodological quality.

**Registration:** https://doi.org/10.17605/OSF.IO/6WZRE

**Protocol and SAP:** https://osf.io/kwqcb/files/osfstorage

## Introduction

Numerous systematic reviews have synthesised the results of trials evaluating exercise therapy interventions after total knee arthroplasty (TKA) (1–8). Due to methodological challenges in the trials included in those reviews; the best exercise therapy approach has not yet been established. Without clear guidance there is limited consensus on what exercise therapy content provides best results after knee replacement (9), and only broad recommendations are presented in the few clinical guidelines available (such as NICE (10)). Randomised clinical trials (RCTs) are considered the gold standard for evaluating causal inference of interventions and are frequently synthesised in systematic reviews forming the foundation of recommendations in clinical guidelines. RCTs are considered more reliable and credible than other sources (e.g., observational studies), as they are considered largely free from most biases, because of the randomisation process, prospective registration, and pre-specified analysis plans (11,12). Prospective registration and consistent reporting of trial objectives and outcomes are central to combatting questionable research practices such as HARKing (hypothesising after results are known), P-hacking (manipulating analyses for statistical significance), or cherry-picking (selecting only supporting data) (13,14). By extension, the definition of a single, pre-specified primary outcome is essential to avoid selective outcome reporting and the risk of being misled by selectively highlighted or altered results (15). Given that clinical practice guidelines—and thus, everyday treatment decisions—often rely on evidence from RCTs, it is crucial to understand whether such core methodological principles are upheld in the trials that underpin them.

Prospective registration is the process of registering details on a trial’s intent, interventions, and outcomes, before enrolling the first participant (16). This practice is required by organisations, journals, and committees (e.g. anyone adhering to the Helsinki declaration or guidelines by the WHO or ICMJE (17–19)). A 2016 meta-epidemiological analysis of Cochrane reviews found that unregistered or retrospectively registered trials tended to show larger treatment effect estimates than prospectively registered trials (20). The authors recommended future review authors report included trials’ registration status and conduct sensitivity analyses to judge the influence of non-prospectively registered studies (20). Outcome switching and selective outcome reporting has been found prevalent even in high impact journals (21). In studies of cardiological trials (22) and samples from high impact journals (23) unregistered and retrospectively registered trials were more likely to report favourable findings than prospectively registered trials. Physical therapy trials published between 2008 and 2012 found only 27% of RCTs reporting registration details (24). More recently, a review of trials published, from 2016 to 2020, across 15 physical therapy journals concluded that the prevalence of post-randomisation bias could not be ruled out, as both prospective registration (only 36% of trials were prospectively registered) and detailed data analysis plans were rare (25). While several systematic reviews have synthesised results from trials of exercise therapy after TKA (1–8), none of them examined whether the included trials adhered to best practice in trial registration or whether timely registration influenced reported effects. Because these trials contribute directly to the evidence base used in clinical decision-making, a closer examination of their methodological transparency is warranted.

### Research aim

To 1) explore (primary) and 2) compare (secondary) the registration characteristics of prospectively registered and non-prospectively registered trials investigating the efficacy of exercise therapy following TKA.

## Methods

This prospectively registered systematic review (25^th^ Oct. 2019, osf.io/6wzre) was reported in accordance with the PRISMA 2020 reporting guidelines (26) (Appendix 1). The protocol document (4^th^ Oct. 2019) and Statistical Analysis Plan document (6^th^ Mar 2025) are publicly available via the Open Science Framework registration at https://osf.io/kwqcb/files/osfstorage [dataset] (27).

### Eligibility Criteria

We used the PICO(T) framework to define eligibility for the review (Figure 1). Participants were human adults who had undergone TKA followed by an exercise therapy intervention. This may have been the active intervention under investigation, or additional to the active intervention if the effects of exercise were reported separately. Comparators could be no exercise, or a type of exercise clearly different from the active group. Any trial outcomes or durations were acceptable for inclusion for the primary aim. Study design was RCTs published after 2000 (launch of clinicaltrials.org) with an accessible full text publication. Exclusion criteria were studies of TKA for reasons other than osteoarthritis, any RCT with multiple published reports of the same dataset and any report labelled “secondary analyses” to avoid double counting participants.

**Figure 1.**
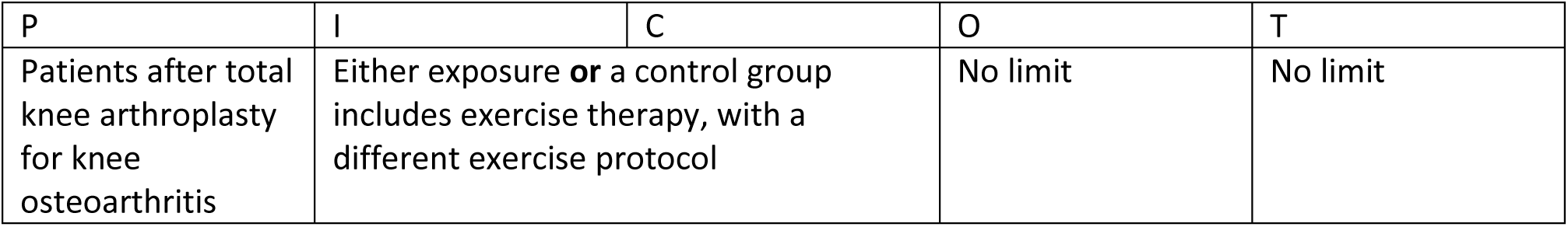
PICOT table. Description of components relating to Population, Intervention, Comparator, Outcome, and Timeframe of interest.

### Information Sources

Medline, EMBASE, CINAHL and the Cochrane Central Register of Randomized Controlled Trials (CENTRAL) were searched on 24^th^ July 2019 and last updated on 12^th^ of August 2024. The reference lists of included trials and relevant systematic reviews (published within the last 5 years) were checked for eligible studies and forward citation tracking carried out through Web of Science. No data was sought from other sources mentioned in the PRISMA checklist (e.g. registries). Any in-accessible reports were sought from corresponding authors via mail or researchgate.net.

### Search Strategy

Search strategies targeted trials reporting on knee arthroplasty and exercise therapy. The Cochrane highly sensitive search strings (adapted to not target drug therapy or animal studies) were applied to primarily identify RCT’s in MEDLINE and EMBASE databases (Cochrane Handbook 6.5 chapter 4.4.7). No limits were applied with built-in filters. Search strings are available in Appendix 2.

### Selection Process

Before screening, all searches were compiled and the Covidence.org duplicate screener was used to remove duplicates automatically. Manual removal of duplicates was carried out ad-hoc, by crosschecking trials as well as group values collected for participant data (BMI, age, n, sex distribution, etc.). Any record flagged as duplicate was verified by another reviewer before removal. Each record was assessed independently by two reviewers, first at the title and abstract level and second at the at full text level. Any reviewer disagreements were discussed until agreement.

Translations (to address the relevance of trials in languages unfamiliar to reviewers) were achieved through Google Translate and DeepL.com. There was no further use of automation tools, AI, or crowdsourcing, for eligibility screening.

### Data Collection Process

Data was extracted (using pilot tested forms) independently by two authors: BG, RB, LH, CJ, CC, TB, and RH. Disagreements were discussed until consensus. No missing data was sought from trial authors as we desired to synthesise publicly available data. Data points only available from figures were extracted and quantified using plotdigitizer.com.

### Data Items (outcomes)

Data extracted included: trial design (registration, multicentre, number of study arms, intervention and control content), publication details (author, year, location), population (age, sex, BMI, type of surgery), interventions (description, initiation, timing, duration, differentiation) and reporting clarity (sample size calculation, adherence to intention-to-treat principles (ITT), adverse events, dropouts). Data for primary endpoints were extracted (mean and standard deviation at baseline and follow-up). The primary outcome was defined using the hierarchy presented in Figure 2, prioritizing protocol-level registration data, supporting transparency, and avoiding post hoc outcome selection. Details of each extracted variable are given in the key table available in the data repository available through the osf.io registration [dataset] (27).

**Figure 2.**
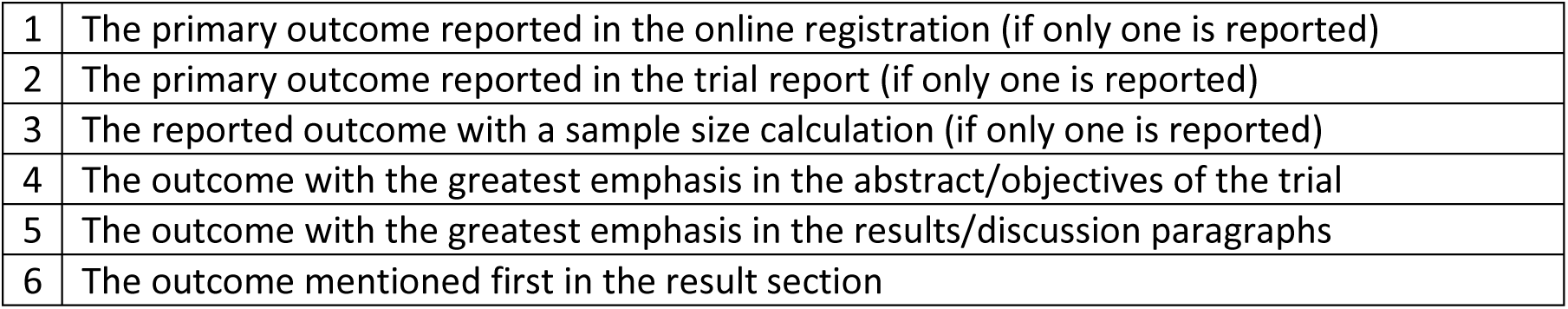
Primary outcome extraction hierarchy. Description of the hierarchy used to select a primary outcome for inclusion. Selection would start at level 1 and if conditions were not met then level 2, checking the below level, and so forth.

For trials with primary outcomes not presented within the domains of pain, disability, performance-based function, or a combination of them (such as a patient reported outcome questionnaire), the primary outcome score was not extracted. In studies with multiple follow-up timepoints, the one specified for the primary analysis was chosen. If not specified, the timepoint closest to the completion of the exercise intervention was chosen. If trials only reported standard errors or 95% confidence intervals and no standard deviations, conversion methods, and small sample size correction for trials with fewer than 60 participants, were used as described in chapter 6.5.2.2 in the Cochrane Handbook 6.5 (28).

### Study Risk of Bias Assessment

Risk of bias of the extracted primary outcome was assessed for the effect of assignment to the intervention (ITT) using the Cochrane Collaborations Risk of Bias 2 tool (RoB2) (29). A single RoB2 assessment was deemed adequate to assess both baseline- and follow-up values (i.e. the comparison between the two). Each study was assessed independently by two assessors and disagreements were discussed until consensus.

### Effect Measures and covariates

To evaluate the influence of registration status on primary outcomes, continuous outcomes extracted from available outcome measures within the domain of pain, disability, performance-based function, or a composite of these (such as a PROM) were used for statistical analyses.

Study characteristics and the RoB2 evaluation are reported according to registration status, trials were grouped based on whether the registration status was prospective, retrospective, or unregistered. Analysis was performed evaluating registration status and study characteristics listed in Table 1, excluding age, sex, BMI, and geographical location.

**Table 1.**
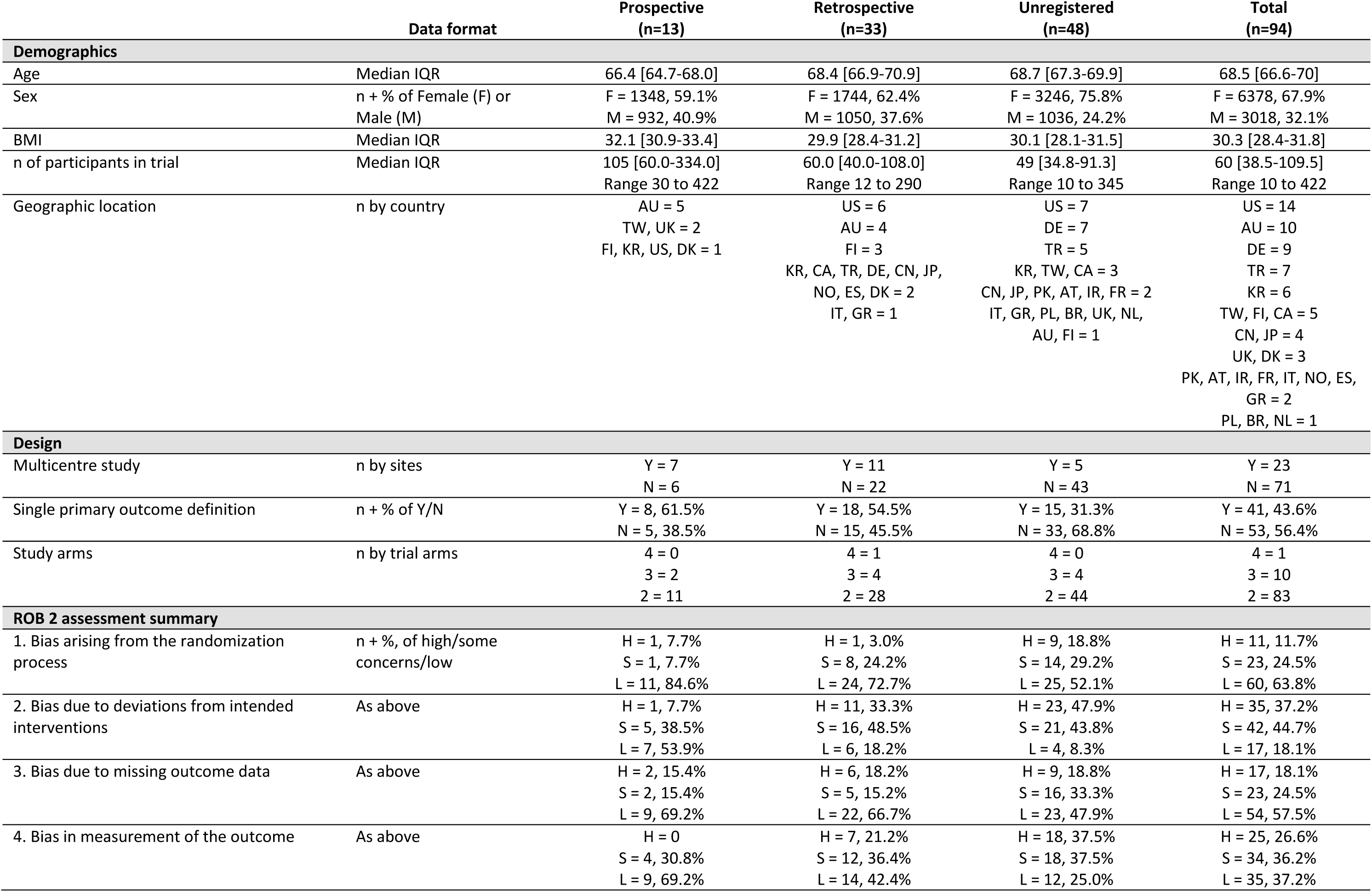
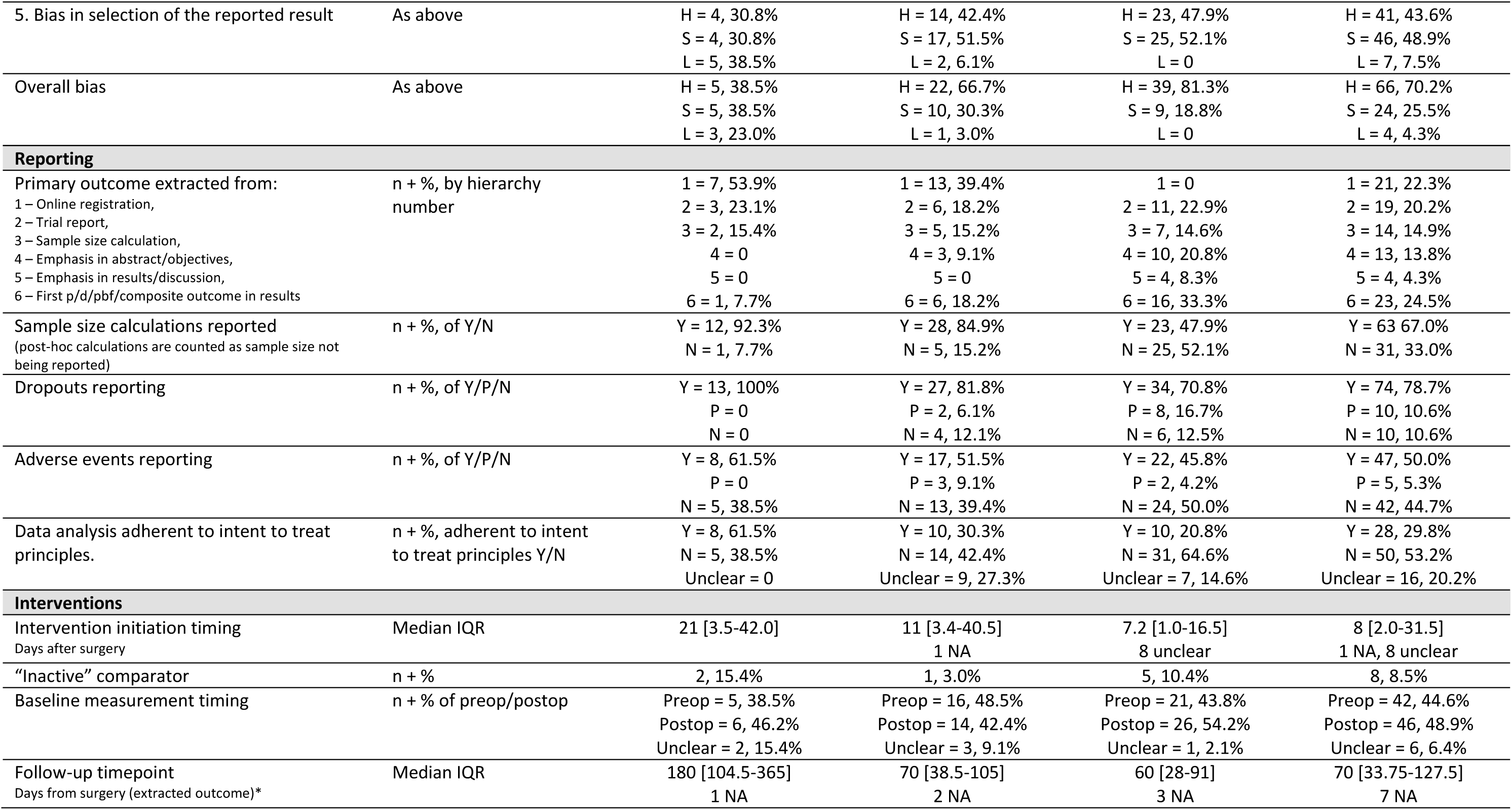
Trial reporting characteristics. Presentation of trial population, design, risk-of-bias, and intervention characteristics. Intention-to-treat definition: All randomised participants clearly included in assigned groups for analysis. * = Extracted either at primary outcome timepoint if specified - otherwise selected directly following end of planned exercise therapy intervention.

**Table 2.**
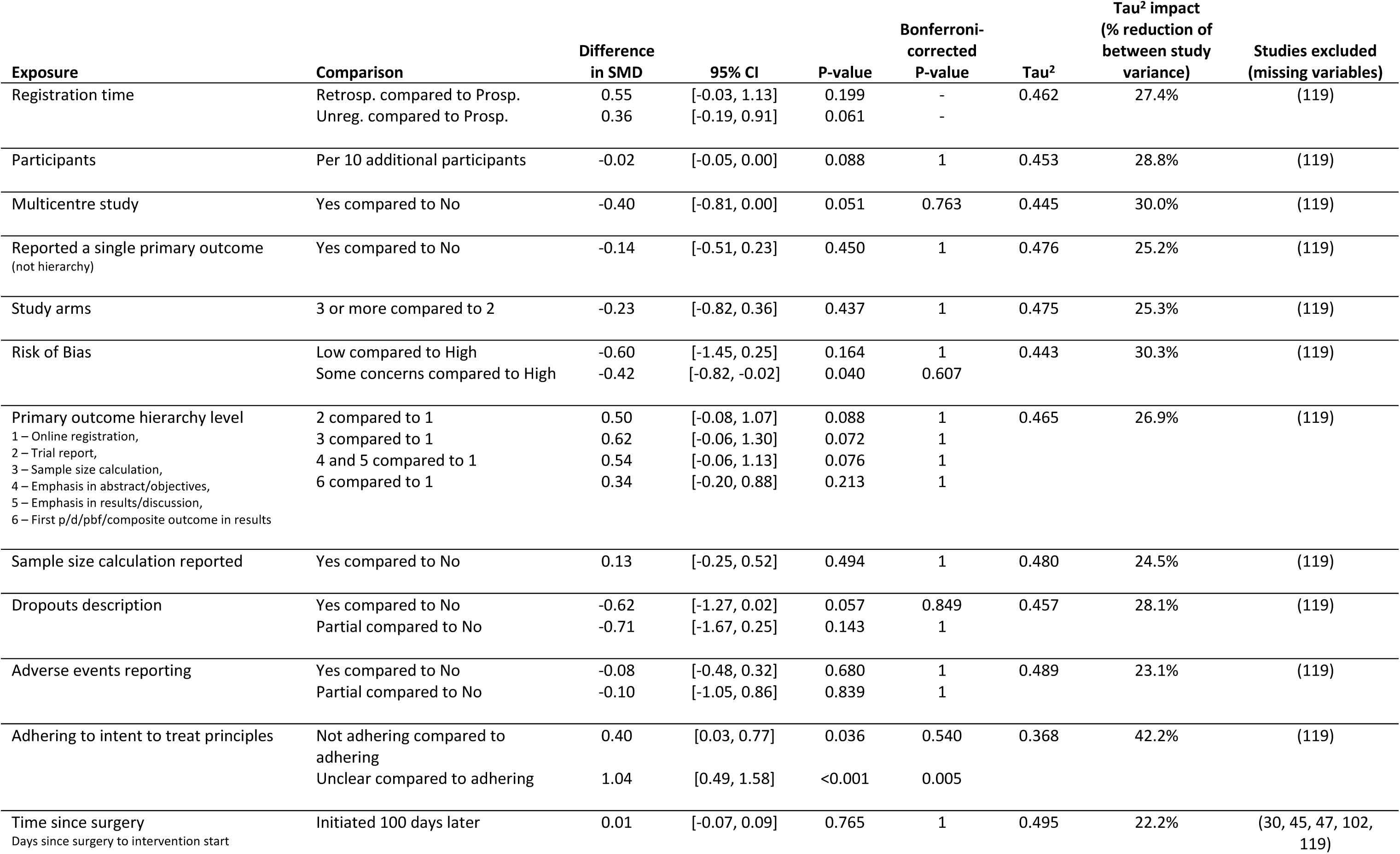
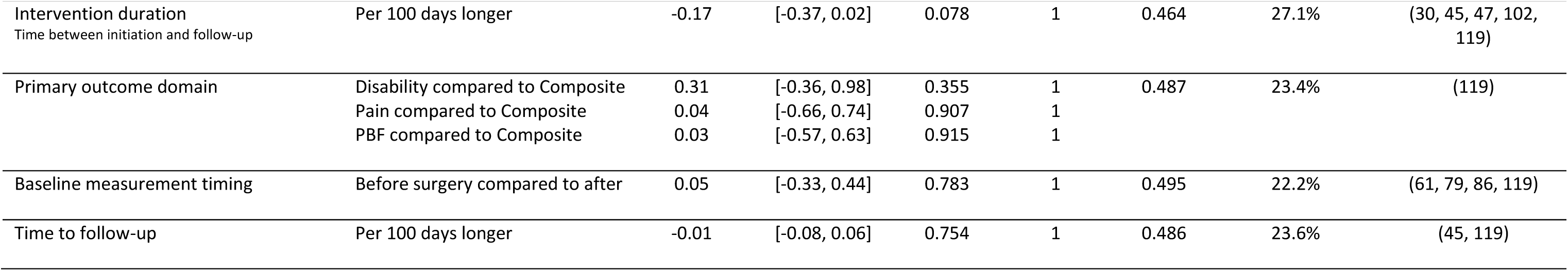
Meta regression analysis. Presentation of the influence of secondary covariates on SMD. Registration status was used as primary exposure for the analysis; hence Bonferroni correction was omitted for registration status. Primary outcome hierarchy levels: 1 = online registration, 2 = trial report, 3 = sample size calculation, 4,5 = emphases in abstract/objectives/results/discussion (grouped as there were too few), 6 = first outcome in results. PBF = Performance-based-function. Includes the 70 trials reporting extractable and comparable primary outcome measures, trials omitted from the analysis are referenced. Raw Tau^2^ = 0.6358.

### Statistics

The statistical analysis plan (SAP) was made publicly available on March 6^th^, 2025 (after latest search, before any analysis) at https://osf.io/kwqcb/files/osfstorage before any analyses were undertaken. For analysis, the primary outcome across the following domains were combined: self-reported pain, disability, performance-based function, and composites of these domains (such as total scores from WOMAC or KOOS questionnaires). The effect of exercise therapy compared to the comparator are expressed as the Hedges’ g standardized mean difference (SMD), a bias corrected adjustment of Cohens d, suggested by the Cochrane Collaboration, and analysed by meta-analysis using random effect modelling (Restricted Maximum Likelihood (REML)). SMD was applied to enable comparison across trials as the focus was on relative effect sizes rather than clinical interpretation per domain. Meta-regression analysis was used to estimate the effect of registration status on the pooled SMD, as well as the effect of the secondary covariates. Additionally, possible effect modification from the study characteristics on registration status was analysed by extending the meta-regressions to include an interaction term with registration status. All models were adjusted for baseline estimates. All extracted outcomes were “score inverted/normalised” so a higher score always indicated improvement/benefit). The overall pooled effects (SMD) were presented so positive outcome are in of favour the intervention group. Differences in effect between studies were expressed as inconsistency (I^2^) as proposed by Higgins et al. (i.e. percent of total variation across studies due to heterogeneity rather than chance) (30). Between study variance was expressed as Tau^2^. The magnitudes of the effect sizes were interpreted using the guidelines from Cohen (0.2 = small, 0.5 = moderate and 0.8 = large) (31). The impact of the secondary covariates in the meta regression was evaluated by their ability to reduce the between study variance (i.e. Tau^2^). All p-values from analysis of the secondary variables were presented with correction for multiple testing by Bonferroni correction, with p-values multiplied by the number of secondary variables. No missing data was imputed.

### Sensitivity analysis

The ICJME formulated specific requirements for prospective registration of trials for medical journals in 2004 (18). As it might not be reasonable to include studies before to this requirement was published, a sensitivity analysis (not pre-specified in protocol) was performed, using only studies after July 1st, 2005 (End of the ICMJE grace period). All sensitivity analysis were performed as described in the statistic section, with this sample.

## Results

As presented in Figure 3, 8,680 records were identified through database searches. After duplicates were removed, 5,222 records were screened, 440 full-text documents assessed, and 94 reports (32–125) included for main analysis, comprising 9,396 participants. Data from 70 (9 prospective, 25 retrospective, 36 unregistered) reports were included in secondary analyses (all in forest plots and meta-regression analyses) (32–34,36,37,41–44,46–49,52,54,58,60,62–70,72–85,87–89,91–94,96,98–101,103,104,107,109–112,114–119,121–125). Reasons for exclusion on full-text level are referenced in Appendix 3. No additional reports were found by searching references or cited-by lists. Figure 4 presents the included trials by publication year with their registration status.

**Figure 3.**
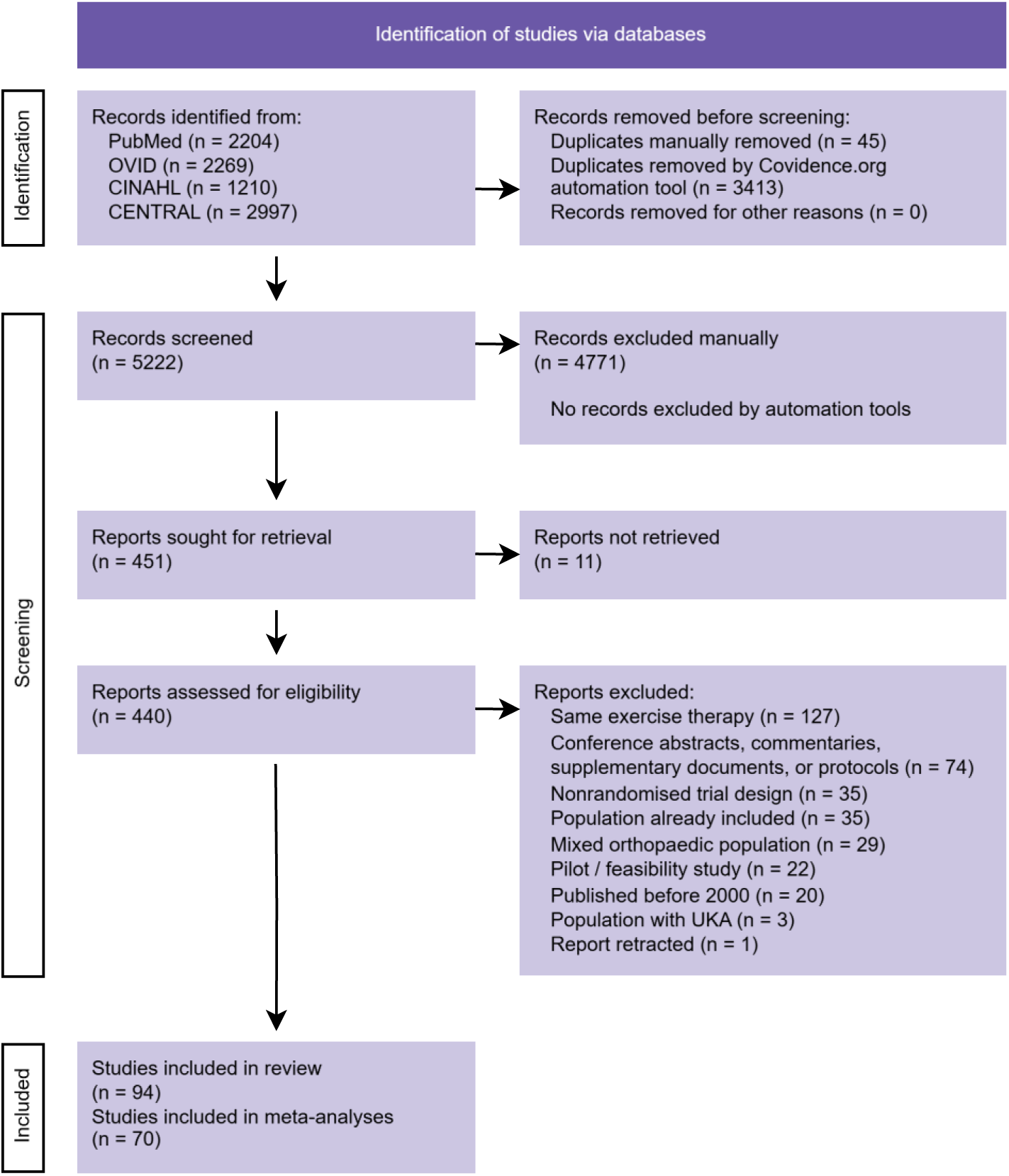
PRISMA flow diagram. Presentation of the process of identifying, screening, and including trials. Reports excluded are cited in Appendix 3.

**Figure 4.**
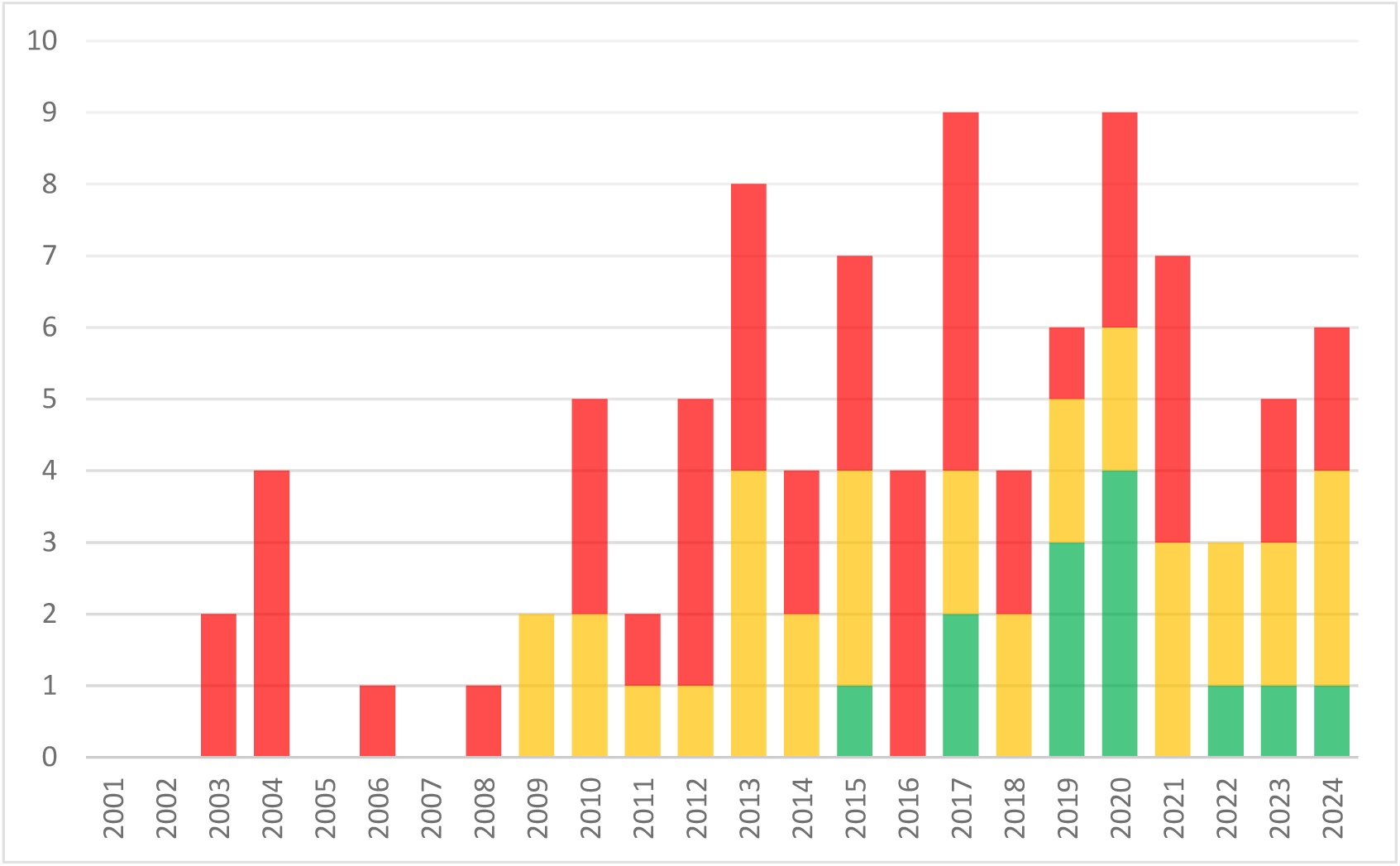
Distribution of included trials over time. X-axis = Publication year, Y-axis = Number of published trials. Green = Prospectively registered, Yellow = Retrospectively registered, Red = Not registered.

Table 1 summarises reporting characteristics of the included trials. For a more detailed overview of each trial, we refer to the raw data extraction sheet and risk of bias assessments available in the data repository available through the registration [dataset] (27).

### Trial design differences

Included trials with prospective registration presented with a higher median of participants, longer time until follow-up (for the extracted primary outcome), and more frequently used multicentre designs than non-prospectively registered trials. A brief description of the intervention and comparator groups, the selected outcome, and risk of bias domain judgements is available for the 94 included trials in Appendix 4.

### Trial reporting differences

As presented in Table 1, in the sample of trials included in this review, prospectively registered trials were more clearly reported than retrospectively registered trials, and both more so than non-registered trials for: definition of a single primary outcome (61.5%/54.5%/31.3%), reporting of sample size calculations (92.3%/84.9%/47.9%), dropout reporting (100%/81.8%/70.8%), adverse events reporting (61.5%/51.5%/45.8%), and adherence to ITT principles (61.5%/30.3%/20.8%).

In the prospectively registered trials: A single primary outcome consistent with a single prospectively registered, unaltered, primary outcome was reported in 4/13 (30.8%) trials (39,62,80,82). The primary outcome was changed (from walking speed to walking distance), during participant enrolment in 1/13 (7.7%) trials (42). Multiple primary outcomes were registered and/or reported in 7/13 (53.9%) trials (54,60,63,71,86,107,108), and 1/13 (7.7%) trials did not report accurately on any of multiple registered outcomes (46). Across all included trials, only three of the prospectively registered trials has a reference to a publicly available statistical analysis plan (42,80,82).

### Risk of Bias in trials

An overview of the risk of bias is available in Table 1, an extended risk of bias table, also presenting experimental and control exercise therapy interventions is available in Appendix 4. The complete raw assessments are available in the data repository [dataset] (27). As presented in the Risk of Bias figure (Figure 5) below, four trial outcomes were judged at overall low risk of bias, 24 at some concern of bias, and 66 at high risk of bias. The prospectively registered trials represented 3/4 (75%) of the trial outcomes judged at low risk of bias, 5/24 (20.8%) of the trial outcomes at some concerns of bias, and 5/66 (7.6%) for the trial outcomes at high risk of bias. Risk of bias in selection of the reported result predominantly arose from a lack of clear definitions of primary outcomes, and for retrospectively registered trials, frequently reporting was not consistent with the registered details.

**Figure 5.**
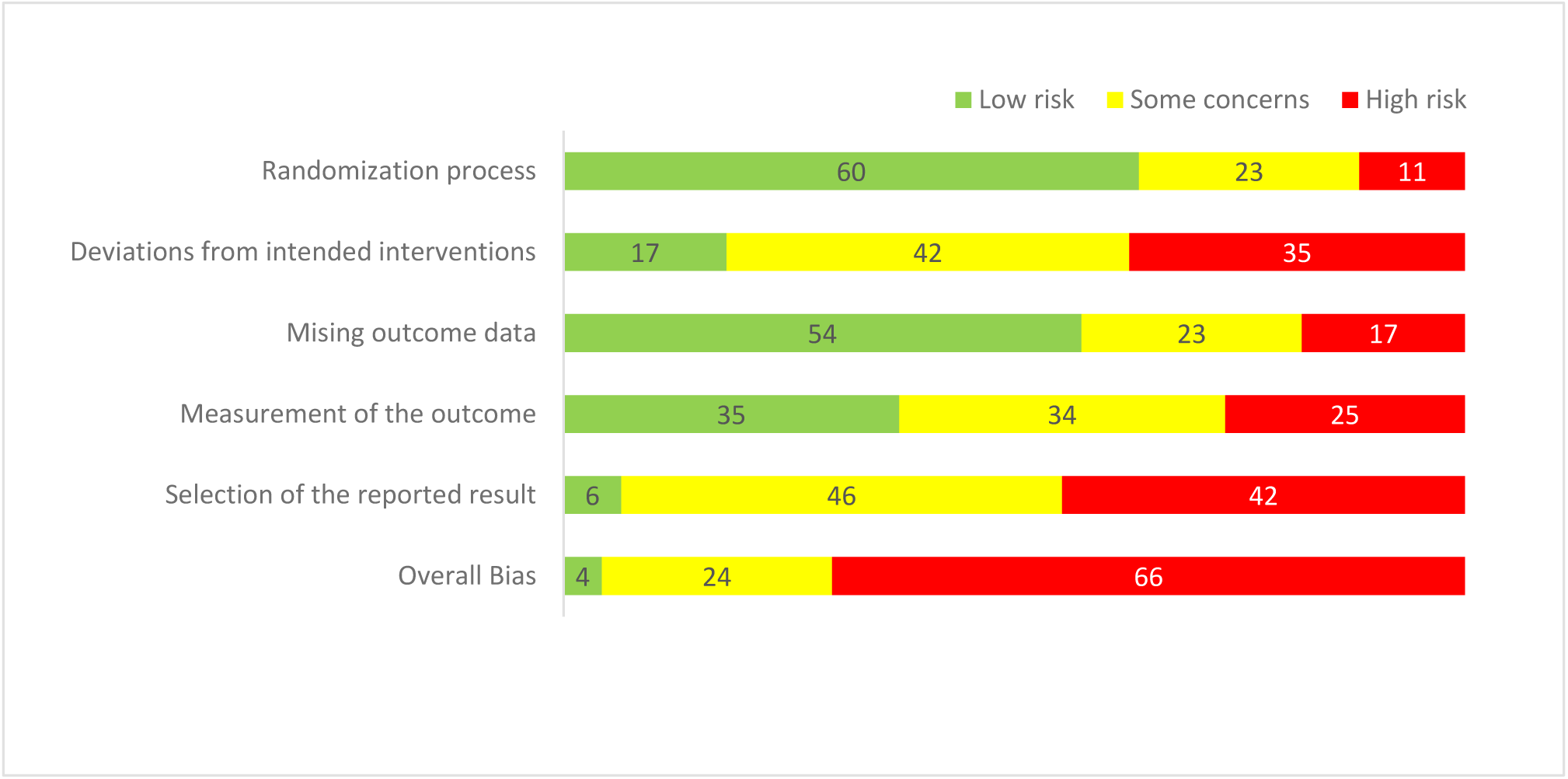
Risk of Bias domains. Summary of risk of bias assessments for the included primary outcome using the Cochrane Risk of Bias 2 (RoB2) tool. n = 94.

### Study effect differences

The combined forest plot (Figure 6) presents the pooled effect size of the primary outcomes from prospective, retrospective, and unregistered trials. The SMD of prospectively registered trials was 0.06 [95% CI: −0.03, 0.16] not favouring either group, whereas the SMD of retrospectively trials was 0.67 [95% CI: 0.22, 1.11] and non-registered trials 0.59 [95% CI: 0.32, 0.86] both favouring the intervention group. Very low I^2^ heterogeneity was present between prospectively registered trials, and high I^2^ heterogeneity for the retrospectively- and unregistered trials. An extended forest plot showing all trials with available data, grouped by registration status and presenting the primary outcome selected, coloured by overall risk of bias, is available in Appendix 5 and funnel plots in Appendix 6.

**Figure 6.**
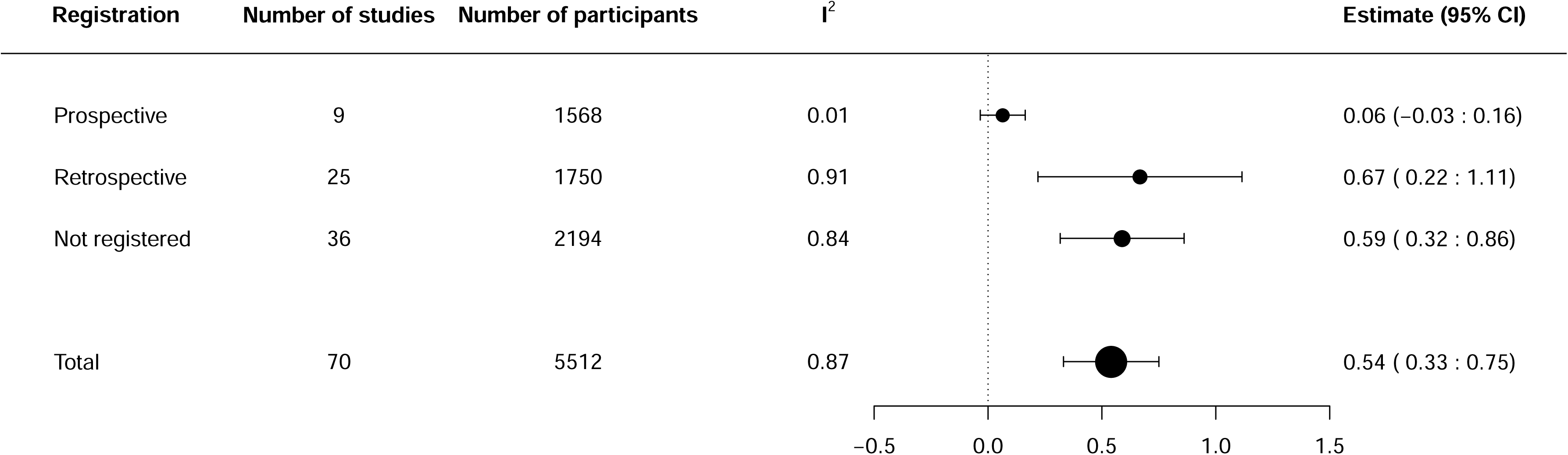
Combined forest plot. Presentation of standardised mean differences of prospective, retrospective, and unregistered RCTs and their pooled SMD. A standardised mean difference favouring the exposure group is represented with values above 0, values below 0 favour the control group.

The meta-regression analysis of the included trials observed the following differences: Retrospectively- and unregistered trials reported a 0.53 and 0.37 larger SMD than the prospectively registered trials, respectively. Comparing primary outcome hierarchy levels, outcomes not prospectively registered as single primary outcome produced a 0.34 to 0.62 larger SMD compared to those prospectively registering it in an online trial registry. Trials defining a single primary outcome (i.e. either in online registration or final report) showed a 0.14 smaller SMDs than trials with no single primary outcome reported. Trials with primary outcomes judged at high risk of bias showed a 0.42 to 0.60 larger SMDs compared to trials with some concerns or low risk of bias. Trials not presenting adequate dropout descriptions showed 0.71 to 0.62 larger SMD compared to those with partial or adequate reporting of dropouts. Compared to trials following ITT principles, those following per protocol reported a 0.4 larger SMD. Those trials where reporting failed to clearly define ITT or PP analysis showed a 1.04 larger SMD and explained the largest proportion of between study variance (Tau^2^). Generally, almost all covariates tested produced large 95% confidence intervals, meaning that large uncertainty is present in the results. Despite the large uncertainty - registration status, ITT adherence, dropout reporting, and outcome definition showed consistent associations with effect size estimates and explained notable between-study variance.

The interaction meta-regression analysis (Appendix 7) showed some estimates with interaction effects, those most pronounced being description of trial dropouts, primary outcome hierarchy level and following ITT principles. However, none of the interactions were statistically significant after correction, and confidence intervals were generally wide, likely due to few observations being available within some levels.

### Sensitivity analysis

Excluding trials not required to be prospectively registered - according to the 1^st^ of July 2005 ICMJE grace period - did not change the outcomes (see Appendix 8). Only minor changes were observed: A slightly larger SMD estimate for non-registered trials, slightly larger confidence intervals, slightly larger Tau^2^ values, and the Eggers test in the funnel plot for non-registered trials changing from statistically significant to showing weak/borderline evidence of asymmetry.

## Discussion

This comprehensive systematic review identified 94 RCTs comparing exercise therapy interventions after TKA, of which 13 were prospectively registered, 33 retrospectively registered, and 48 unregistered. We found that non-prospectively registered trials tended to report larger effect sizes than prospectively registered ones, a similar finding to a 2016 meta-epidemiological study of Cochrane reviews (20). Fewer than half of the included trials clearly defined a single primary outcome. Notably, many trials listed multiple “primary” outcomes without the methodological elements typically required when more than one primary endpoint is intended (e.g., an explicit success criterion such as co-primary vs. at-least-one, prespecified multiplicity control, and sample-size planning aligned with that choice) (126). Our observation is not intended as a critique of legitimate co-primary or dual-primary frameworks per se, but of insufficient pre-specification in trials that simply labelled several outcomes as “primary”. Such practice likely complicates consistent outcome reporting and interpretation and may contribute to inflated or unstable effect estimates when combined with non-prospective registration.

Pinto et al. (2013) in their cohort study of 200 randomly selected physiotherapy trials (from 2009) found that 67/200 (34%) were registered and only 5/200 (2.5%) trials had prospectively and adequately defined primary outcomes (127). In comparison the current review found 46/94 (48%) trials registered and 4/94 (4.3%) of those prospectively and adequately registered. Babu et al. reported that only 29% of trials published in 2012 by physiotherapy specialty journals were registered and suggested that registration practice appeared to be improving (24). As seen in Figure 4, there does not appear to be a year-on-year improvement to the rate that trials are prospectively registered. Rather, Figure 4 indicates that the rate has remained unchanged since 2013.

In their review of RCTs involving musculoskeletal physical therapy published in 15 physiotherapy journals from 2016 to 2020, Riley et al. found that 116/138 (84%) RCT’s were registered and 49/138 (36%) prospectively (25), which is significantly more than in the 14% prospectively registered RCT’s in the current review. Further, Riley et al. described that reporting in the 42 of the 49 (86%) prospectively registered RCT’s was consistent with prospectively registered outcomes. For the remainder of the 138 included RCT’s, post-randomisation bias could not be ruled out (25). In the current review, prospective registration was less prevalent, due to inclusion of older studies (2000-2025 vs. 2016-2020). Considering the differences in effect sizes between prospectively registered and non-prospectively registered trials in the current sample, and that only three studies reported a statistical analysis plan; post-randomisation bias cannot be ruled out in RCTs comparing exercise therapy interventions after total knee replacement either. Only 30.8% of prospectively registered trials reported a single primary outcome consistent with their registration. While this is an example, that timely registration of outcomes enables readers (and peer-reviewers) to assess the coherence between registration and reporting; it also shows that higher methodological quality isn’t automatically assured, just by achieving timely registration. Further, although the ability to register multiple primary outcomes may reflect flexibility in registry systems, our findings suggest this can complicate consistent outcome reporting and interpretation.

A synthesis of meta-epidemiological studies of rehabilitation studies concluded that inadequate or unclear randomisation sequence generation and allocation concealment were likely to produce exaggerated effect estimates for continuous outcomes, and higher risk of reporting bias appeared associated with an overestimation of effects (128). In the current review, most of the extracted outcomes were at low risk of bias in the randomisation process domain; suggesting that the difference in effect between the prospectively, retrospectively and unregistered trials is likely not explained by randomisation and concealment bias. The reporting bias domain presented with the highest percent of outcomes at some concern or high risk of bias in retrospectively and unregistered trials, which suggests that reporting bias may, to some extent, explain why the average effect size in those trials was larger than in prospectively registered trials.

Higher adherence to CONSORT guideline has been associated with lower risk of bias in trials published in rehabilitation journals (129), underscoring the importance of clear and transparent reporting. The current review had no formal assessment of included trials’ adherence to reporting guidelines such as CERT (130), TIDieR (131), or CONSORT (132) – because some trials were published prior to the guidelines.

This review followed a comprehensive, prospectively registered protocol and statistical analysis plan. All steps of study selection and data extraction were done in double, and the study is reported transparently by adhering to the PRISMA checklist and providing full access to all extracted data. The review does have limitations: The SAP was registered after data-extraction but before analysis, but all changes to the review are reported transparently in the amendments section. Due to the method chosen to select the primary outcome, the results in the current meta-analyses should not be interpreted as an indication of the overall effect of exercise therapy after TKA, but rather the effect as measured with those specific outcomes. The findings represent the effect sizes on outcomes selected using the hierarchy and aggregates outcomes representing different constructs (pain, disability, and performance-based outcomes). The use of standardized mean differences is recommended for combining results on different scales but only within the *same* construct (Cochrane handbook 6.5, section 15.5.3). Still, we combined different constructs because we were interested in comparing effect sizes of primary outcomes and whether the effect sizes were influenced by the trial characteristics, not clinical interpretation.

Our review findings show that non-prospectively registered RCT’s of exercise therapy interventions after TKA tend to report larger, and possibly inflated, effect sizes compared to more methodologically rigorous prospectively registered trials. Caution is warranted because inflated effect estimates from inadequately registered trials could influence treatment recommendations, if it is not accounted for in the evidence syntheses informing clinical practice guidelines. We concur with Dechartres et al. and likewise advocate that peer-reviewers and publishers more systematically verify RCT reports against their registration, and employ suitable reporting guidelines to ensure transparent reporting (20). Further, we also encourage that future systematic reviews (of RCT’s) clearly define and report registration characteristics, and conduct sensitivity analyses to evaluate the impact of including secondary outcome data (20), and clearly designate the origin of aggregated outcomes (primary/secondary and prospective/retrospective/unregistered). Ultimately, improving registration practices is a shared responsibility between researchers (including peer-reviewers), funders, and journals —and doing so will meaningfully enhance the transparency, reproducibility, and clinical relevance of future trials.

## Conclusion

This systematic review found that prospective registration and clear definitions of a single primary outcome are rare in trials comparing effects of exercise therapy following TKA. Prospectively registered trials generally presented with larger sample sizes, clearer reporting, lower risk of bias, and smaller effect size estimates with less variance, compared to retrospectively- and unregistered trials. These findings suggest that prospective registration is an important indicator of methodological quality and that greater adherence to prospective registration and transparent reporting is essential to strengthen the evidence base that informs guidelines for clinical practice in exercise therapy following TKA.

### Registration and Protocol

This review was registered with Open Science Framework (OSF.io).

Protocol registration (25^th^ Oct 2019) available at https://doi.org/10.17605/OSF.IO/6WZRE

Protocol document (4^th^ Oct 2019) available at https://osf.io/vmpu6

Statistical Analysis Plan (SAP) (6^th^ Mar 2025) available at https://osf.io/v5w4x

### Amendments since protocol registration

Available through registration on OSF, in original protocol document, and statistical analysis plan document. Since the original protocol was published, the following changes were made:

### Amendment(s) to extracted outcome variables

#### 1^st^ Amendment

Change from three extracted outcomes (pain, disability, performance-based function), using the hierarchy to using the hierarchy to select a single outcome.

Reason: As we always intended to compare the primary outcome in each trial across domains, it became redundant to also collect “secondary primary outcomes” in the other domains. We believed that sticking with a single outcome would make interpretations more focused on the methodological choices made in trials, the conciseness of their reporting, and its impact on the effect estimates.

Timing: This change was made after the search was originally completed, during May 2021, before data-extraction was initiated on June 1^st^, 2021.

#### 2^nd^ Amendment

Two data variables were dropped: publisher impact factor and external funding.

Reason: It turned out to be very problematic and time consuming to find verifiable and accurate impact factor values for the time the trials were published. External funding was omitted as it was considered outside the scope of the current review.

Timing: The variables were dropped shortly after data-extraction was initiated in 2020.

### Amendment(s) to eligibility criteria since registration

#### Amendment

Trials investigating the effect of a non-exercise component, added to one of two groups with the same underlying exercise intervention were excluded from the review.

Reason: When the search was updated in 2024, during full-text assessment, it became clear that a large proportion of trials were investigating the effect of a non-exercise variable (such as drugs, massage, behaviour change, or continuous passive motion), with the same underlying exercise therapy in groups (e.g. comparing effects of 1) exercise therapy and massage against 2) exercise therapy alone). In the protocol research question was formulated with the intent of finding trials comparing different exercise modes.

Timing: This was implemented during title/abstract screening during the update of the search initiated on 12^th^ of August 2024, with the change noted on 15^th^ of November 2024.

### Amendment(s) to analyses since registration

#### 1^st^ Amendment

Grouping at three instead of two levels. Registration status was changed from Prospective- and Non-prospective registration, to Prospectively-, Retrospectively-, and Unregistered.

Reason: During the last search update, it was clear that several trials were not timely registered due to oversight, and that some these retrospectively registered trials were better reported than most non-registered trials. By presenting 3 layers of comparison, we hoped to enable a more nuanced representation of non-prospectively registered trials and clarify differences between trials with retrospective registration and no registration.

Timing: This was implemented, and described in the SAP, which was made available online on 6^th^ of March 2025, before statistical analyses were undertaken.

#### 2^nd^ Amendment

The Chi-square test or Fishers exact test was planned for analyses of differences between categorical variables reporting study characteristics, but none were conducted.

Reason: The descriptive table 1 is intended to be descriptive and provide context, therefore statistical testing for significant differences may be misleading.

Timing: Implemented after the SAP was registered on 6^th^ of March 2025, but before data analysis was initiated.

## Data Availability

All data produced are available online at the study registration at open science framework (OSF)

https://doi.org/10.17605/OSF.IO/6WZRE

## CRediT Contributions

Conception and design – RH, CC, TB, BG, CJ. Trial selection (title/abstract and full text screening) – BG, LH, RB, TB. Collection and assembly of data – BG, TB, CJ, RH, LH, CC, RB. Analysis and interpretation of the data – BG, TB, CJ, TK. Statistical expertise – TK, CJ. Drafting of the article – BG, TB. Critical revision of the article for important intellectual content – TB, CJ, TK, RH, LH, CC, RB. All authors (BG, TB, RB, RH, CC, LH, CJ, TK) have full access to all data collected and approved final decision to submit the manuscript for publication.

## Availability of Data, Code, and Other Materials

The following data and supplementary files are available for use under the licence CC-BY 4.0 International, at osf.io/kwqcb/files/osfstorage [dataset] (27):

Datasets available: Statistical code files (R), dataset file, data key file, and Risk of Bias assessments.

Supplementary file names: Appendix 1 PRISMA 2020 checklist, Appendix 2 Search strings, Appendix 3 Excluded studies, Appendix 4 RoB2 table, Appendix 5 Extended forest plot, Appendix 6 Funnel plots, Appendix 7 Meta-regression interaction, Appendix 8 Sensitivity analyses.

## References

1. Artz N, Elvers KT, Lowe CM, Sackley C, Jepson P, Beswick AD. Effectiveness of physiotherapy exercise following total knee replacement: systematic review and meta-analysis. BMC Musculoskeletal Disorders [Internet]. 2015 Jan;16(1). Available from: http://search.ebscohost.com/login.aspx?direct=true&db=rzh&AN=109723492&site=ehost-live

2. Florez-García M, García-Pérez F, Curbelo R, Pérez-Porta I, Nishishinya B, Rosario Lozano MP, et al. Efficacy and safety of home-based exercises versus individualized supervised outpatient physical therapy programs after total knee arthroplasty: a systematic review and meta-analysis. Knee Surg Sports Traumatol Arthrosc. 2017 Nov;25(11):3340–53.

3. Alrawashdeh W, Eschweiler J, Migliorini F, El Mansy Y, Tingart M, Rath B. Effectiveness of total knee arthroplasty rehabilitation programmes: A systematic review and meta-analysis. J Rehabil Med. 2021 Jun 2;53(6):jrm00200.

4. Sattler LN, Hing WA, Vertullo CJ. What is the evidence to support early supervised exercise therapy after primary total knee replacement? A systematic review and meta-analysis. BMC Musculoskeletal Disorders. 2019 Jan 29;20(1):1–11.

5. Chaudhry YP, Hayes H, Wells Z, Papadelis E, Khanuja HS, Deirmengian C. Not All Patients Need Supervised Physical Therapy After Primary Total Knee Arthroplasty: A Systematic Review and Meta-Analysis. Cureus. 2023 Feb;15(2):e35232.

6. Konnyu KJ, Thoma LM, Cao W, Aaron RK, Panagiotou OA, Bhuma MR, et al. Rehabilitation for Total Knee Arthroplasty: A Systematic Review. Am J Phys Med Rehabil. 2023 Jan 1;102(1):19–33.

7. Koster A, Stevens M, van Keeken H, Westerveld S, Seeber GH. Effectiveness and therapeutic validity of physiotherapeutic exercise starting within one year following total and unicompartmental knee arthroplasty for osteoarthritis: a systematic review. Eur Rev Aging Phys Act. 2023 Mar 29;20(1):8.

8. Skoffer B., Dalgas U., Mechlenburg I. Progressive resistance training before and after total hip and knee arthroplasty: a systematic review. Clin Rehabil. 2015;29(1):14–29.

9. Blom AW, Artz N, Beswick AD, Burston A, Dieppe P, Elvers KT, et al. Physiotherapy exercise after total knee replacement: systematic review, survey of provision and feasibility randomised controlled trial. In: Improving patients’ experience and outcome of total joint replacement: the RESTORE programme [Internet]. NIHR Journals Library; 2016 [cited 2025 Sep 24]. Available from: https://www.ncbi.nlm.nih.gov/books/NBK379640/

10. Recommendations | Joint replacement (primary): hip, knee and shoulder | Guidance | NICE [Internet]. NICE; 2020 [cited 2025 Feb 6]. Available from: https://www.nice.org.uk/guidance/ng157/chapter/Recommendations#postoperative-care

11. Frieden TR. Evidence for Health Decision Making — Beyond Randomized, Controlled Trials. New England Journal of Medicine. 2017 Aug 3;377(5):465–75.

12. Deaton A, Cartwright N. Understanding and misunderstanding randomized controlled trials. Soc Sci Med. 2018 Aug;210:2–21.

13. Andrade C. HARKing, Cherry-Picking, P-Hacking, Fishing Expeditions, and Data Dredging and Mining as Questionable Research Practices. J Clin Psychiatry. 2021 Feb 18;82(1):20f13804.

14. Cook C, Garcia AN. Post-randomization bias. Journal of Manual & Manipulative Therapy. 2020 Mar 14;28(2):69–71.

15. Andrade C. The Primary Outcome Measure and Its Importance in Clinical Trials. J Clin Pshychiatry. 2015 Oct 21;76(10).

16. Bandholm T, Christensen R, Thorborg K, Treweek S, Henriksen M. Preparing for what the reporting checklists will not tell you: the PREPARE Trial guide for planning clinical research to avoid research waste. British Journal of Sports Medicine. 2017 Oct;51(20):1494–501.

17. WMA. World Medical Association. WMA Declaration of Helsinki - Ethical Principles for Medical Research Involving Human Subjects. 2017; Available from: https://www.wma.net/policies-post/wma-declaration-of-helsinki-ethical-principles-for-medical-research-involving-human-subjects/. Accessed November 15, 2017

18. ICMJE | About ICMJE | Clinical Trials Registration [Internet]. [cited 2025 May 15]. Available from: https://www.icmje.org/about-icmje/faqs/clinical-trials-registration/

19. WHO. Trial registration [Internet]. [cited 2025 May 15]. Available from: https://www.who.int/tools/clinical-trials-registry-platform/network/trial-registration

20. Dechartres A, Ravaud P, Atal I, Riveros C, Boutron I. Association between trial registration and treatment effect estimates: a meta-epidemiological study. BMC Med. 2016 Jul 4;14(1):100.

21. Goldacre B, Drysdale H, Dale A, Milosevic I, Slade E, Hartley P, et al. COMPare: a prospective cohort study correcting and monitoring 58 misreported trials in real time. Trials. 2019 Feb 14;20(1):118.

22. Emdin C, Odutayo A, Hsiao A, Shakir M, Hopewell S, Rahimi K, et al. Association of cardiovascular trial registration with positive study findings: Epidemiological Study of Randomized Trials (ESORT). JAMA Intern Med. 2015 Feb;175(2):304–7.

23. Gopal AD, Wallach JD, Aminawung JA, Gonsalves G, Dal-Ré R, Miller JE, et al. Adherence to the International Committee of Medical Journal Editors’ (ICMJE) prospective registration policy and implications for outcome integrity: a cross-sectional analysis of trials published in high-impact specialty society journals. Trials. 2018 Aug 23;19(1):448.

24. Babu AS, Veluswamy SK, Rao PT, Maiya AG. Clinical Trial Registration in Physical Therapy Journals: A Cross-Sectional Study. Physical Therapy. 2014 Jan 1;94(1):83–90.

25. Riley SP, Swanson BT, Shaffer SM, Sawyer SF, Cleland JA. The Unknown Prevalence of Postrandomization Bias in 15 Physical Therapy Journals: A Methods Review. J Orthop Sports Phys Ther. 2021 Nov;51(11):542–50.

26. Page MJ, McKenzie JE, Bossuyt PM, Boutron I, Hoffmann TC, Mulrow CD, et al. The PRISMA 2020 statement: an updated guideline for reporting systematic reviews. BMJ. 2021 Mar 29;372:n71.

27. Grønfeldt B, Husted R, Brødsgaard R, Holst L, Chapple C, Kallemose T, et al. Trial registration and reporting characteristics of trials investigating exercise-based rehabilitation following total knee arthroplasty: A systematic review [Internet]. OSF; 2019 [cited 2025 Jun 26]. Available from: https://osf.io/kwqcb/

28. Higgins J, Li T, Deeks J (Editors). Chapter 6: Choosing effect measures and computing estimates of effect [last updated August 2023]. In: Higgins JPT, Thomas J, Chandler J, Cumpston M, Li T, Page MJ, Welch VA (editors) Cochrane Handbook for Systematic Reviews of Interventions version 65 Cochrane, 2024 [Internet]. [cited 2025 Jun 5]. Available from: Available from cochrane.org/handbook.

29. Sterne JAC, Savović J, Page MJ, Elbers RG, Blencowe NS, Boutron I, et al. RoB 2: a revised tool for assessing risk of bias in randomised trials. BMJ. 2019 Aug 28;l4898.

30. Higgins JPT. Measuring inconsistency in meta-analyses. BMJ. 2003 Sep 6;327(7414):557–60.

31. Cohen J. Statistical Power Analysis for the Behavioral Sciences. 2. United States of America: Lawrence Erlbaum Associates; 1988. 579 p.

32. Akbaba YA, Yeldan I, Guney N, Ozdincler AR. Intensive supervision of rehabilitation programme improves balance and functionality in the short term after bilateral total knee arthroplasty. Knee Surg Sports Traumatol Arthrosc. 2016 Jan;24(1):26–33.

33. Alsayani KYA, Baş Aslan U, Bayrak G, Şavkın R, Büker N, Güngör HR. Comparison of the effectiveness of late-phase clinic-based and home-based progressive resistance training in female patients with total knee arthroplasty. Physiotherapy Theory and Practice. 2024 Aug 2;40(8):1687–98.

34. An J, Son YW, Lee BH. Effect of Combined Kinematic Chain Exercise on Physical Function, Balance Ability, and Gait in Patients with Total Knee Arthroplasty: A Single-Blind Randomized Controlled Trial. IJERPH. 2023 Feb 16;20(4):3524.

35. Bäcker HC, Wu CH, Schulz MRG, Weber-Spickschen TS, Perka C, Hardt S. App-based rehabilitation program after total knee arthroplasty: a randomized controlled trial. Arch Orthop Trauma Surg. 2021 Sep;141(9):1575–82.

36. Bade MJ, Struessel T, Dayton M, Foran J, Kim RH, Miner T, et al. Early High-Intensity Versus Low-Intensity Rehabilitation After Total Knee Arthroplasty: A Randomized Controlled Trial. Arthritis Care & Research. 2017 Sep;69(9):1360–8.

37. Bily W, Franz C, Trimmel L, Loefler S, Cvecka J, Zampieri S, et al. Effects of Leg-Press Training With Moderate Vibration on Muscle Strength, Pain, and Function After Total Knee Arthroplasty: A Randomized Controlled Trial. Archives of Physical Medicine and Rehabilitation. 2016 Jun;97(6):857– 65.

38. Bini S, Mahajan J. Clinical outcomes of remote asynchronous telerehabilitation are equivalent to traditional therapy following total knee arthroplasty: A randomized control study. J Telemed Telecare. 2017 Feb;23(2):239–47.

39. Bohl DD, Li J, Calkins TE, Darrith B, Edmiston TA, Nam D, et al. Physical Therapy on Postoperative Day Zero Following Total Knee Arthroplasty: A Randomized, Controlled Trial of 394 Patients. The Journal of Arthroplasty. 2019 Jul;34(7):S173–S177.e1.

40. Bradbury TL, McConnell MJ, Whitacre D, Naylor BH, Gibson BT, DeCook CA. A Remote Physical Therapy Program Demonstrates Similar Outcomes Compared to In-Person, Supervised Physical Therapy After Same-Day Discharge Total Knee Arthroplasty: A Randomized Clinical Trial. The Journal of Arthroplasty. 2024 Nov;39(11):2725–2730.e4.

41. Bruun-Olsen V, Heiberg KE, Wahl AK, Mengshoel AM. The immediate and long-term effects of a walking-skill program compared to usual physiotherapy care in patients who have undergone total knee arthroplasty (TKA): a randomized controlled trial. Disability and Rehabilitation. 2013 Nov;35(23):2008–15.

42. Buhagiar MA, Naylor JM, Harris IA, Xuan W, Kohler F, Wright R, et al. Effect of Inpatient Rehabilitation vs a Monitored Home-Based Program on Mobility in Patients With Total Knee Arthroplasty: The HIHO Randomized Clinical Trial. JAMA. 2017 Mar 14;317(10):1037.

43. Carozzo S, Vatrano M, Coschignano F, Battaglia R, Calabrò RS, Pignolo L, et al. Efficacy of Visual Feedback Training for Motor Recovery in Post-Operative Subjects with Knee Replacement: A Randomized Controlled Trial. JCM. 2022 Dec 11;11(24):7355.

44. Çetinkaya F, Karakoyun A. The effects of elastic band exercise on the pain, kinesiophobia, functional, and psychological status after total knee arthroplasty: a randomized controlled trial. Clin Rheumatol. 2022 Oct;41(10):3179–88.

45. Chang HL, Hsu MF, Wong TH, Chung YC, Huang HL. Effects of a Hybrid Teaching Program on Lower Limb Muscle Strength, Knee Function, and Depression in Older Adults After Total Knee Replacement: A Randomized Controlled Trial. Research in Gerontological Nursing. 2024 Jan;17(1):31–40.

46. Cheng YY, Liu CC, Lin SY, Lee CH, Chang ST, Wang SP. Comparison of the Therapeutic Effects Between Isokinetic and Isotonic Strength Training in Patients After Total Knee Replacement: A Prospective, Randomized Controlled Trial. Orthopaedic Journal of Sports Medicine. 2022 Jun 1;10(6):23259671221105852.

47. Chow TP, Ng GY. Active, passive and proprioceptive neuromuscular facilitation stretching are comparable in improving the knee flexion range in people with total knee replacement: a randomized controlled trial. Clin Rehabil. 2010 Oct;24(10):911–8.

48. Christiansen CL, Bade MJ, Davidson BS, Dayton MR, Stevens-Lapsley JE. Effects of Weight-Bearing Biofeedback Training on Functional Movement Patterns Following Total Knee Arthroplasty: A Randomized Controlled Trial. Journal of Orthopaedic & Sports Physical Therapy. 2015 Sep;45(9):647– 55.

49. Codine Ph, Dellemme Y, Denis-Laroque F, Herisson Ch. The use of low velocity submaximal eccentric contractions of the hamstring for recovery of full extension after total knee replacement: A randomized controlled study. Isokinetics and Exercise Science. 2004 Aug;12(3):215–8.

50. Crawford DA, Duwelius PJ, Sneller MA, Morris MJ, Hurst JM, Berend KR, et al. 2021 Mark Coventry Award: Use of a smartphone-based care platform after primary partial and total knee arthroplasty: a prospective randomized controlled trial. The Bone & Joint Journal. 2021 Jun 1;103-B(6 Supple A):3–12.

51. DeJong G, Hsieh CJ, Vita MT, Zeymo A, Boucher HR, Thakkar SC. Innovative Devices Did Not Provide Superior Total Knee Arthroplasty Outcomes in Post-Operative Rehabilitation: Results From a Four-Arm Randomized Clinical Trial. The Journal of Arthroplasty. 2020 Aug;35(8):2054–65.

52. Den Hertog A, Gliesche K, Timm J, Mühlbauer B, Zebrowski S. Pathway-controlled fast-track rehabilitation after total knee arthroplasty: a randomized prospective clinical study evaluating the recovery pattern, drug consumption, and length of stay. Arch Orthop Trauma Surg. 2012 Aug;132(8):1153–63.

53. Khanli MM, Akbari M, Amiri A. The Effect of Early Hip-strengthening on Physical Function in Patients With Unilateral Total Knee Arthroplasty. J Arak Uni Med Sci. 2021 Feb 1;23(6):912–25.

54. Do K, Yim J. Effects of Muscle Strengthening around the Hip on Pain, Physical Function, and Gait in Elderly Patients with Total Knee Arthroplasty: A Randomized Controlled Trial. Healthcare. 2020 Nov 17;8(4):489.

55. Doerfler D, Gurney B, Mermier C, Rauh M, Black L, Andrews R. High-Velocity Quadriceps Exercises Compared to Slow-Velocity Quadriceps Exercises Following Total Knee Arthroplasty: A Randomized Clinical Study. Journal of Geriatric Physical Therapy. 2016 Oct;39(4):147–58.

56. Eisermann U, Haase I, Kladny B. Computer-Aided Multimedia Training in Orthopedic Rehabilitation: American Journal of Physical Medicine & Rehabilitation. 2004 Sep;83(9):670–80.

57. Evgeniadis G, Beneka A, Malliou P, Mavromoustakos S, Godolias G. Effects of pre- or postoperative therapeutic exercise on the quality of life, before and after total knee arthroplasty for osteoarthritis. BMR. 2008 Sep 1;21(3):161–9.

58. Eymir M, Erduran M, Ünver B. Active heel-slide exercise therapy facilitates the functional and proprioceptive enhancement following total knee arthroplasty compared to continuous passive motion. Knee Surg Sports Traumatol Arthrosc. 2021 Oct;29(10):3352–60.

59. Fleischman AN, Crizer MP, Tarabichi M, Smith S, Rothman RH, Lonner JH, et al. 2018 John N. Insall Award: Recovery of Knee Flexion With Unsupervised Home Exercise Is Not Inferior to Outpatient Physical Therapy After TKA: A Randomized Trial. Clin Orthop Relat Res. 2019 Jan;477(1):60–9.

60. Fransen M, Nairn L, Bridgett L, Crosbie J, March L, Parker D, et al. Post-Acute Rehabilitation After Total Knee Replacement: A Multicenter Randomized Clinical Trial Comparing Long-Term Outcomes. Arthritis Care & Research. 2017 Feb;69(2):192–200.

61. Fung V, Ho A, Shaffer J, Chung E, Gomez M. Use of Nintendo Wii Fit^TM^ in the rehabilitation of outpatients following total knee replacement: a preliminary randomised controlled trial. Physiotherapy. 2012 Sep;98(3):183–8.

62. Hamilton DF, Beard DJ, Barker KL, Macfarlane GJ, Tuck CE, Stoddart A, et al. Targeting rehabilitation to improve outcomes after total knee arthroplasty in patients at risk of poor outcomes: randomised controlled trial. BMJ. 2020 Oct 13;m3576.

63. Han ASY, Nairn L, Harmer AR, Crosbie J, March L, Parker D, et al. Early Rehabilitation After Total Knee Replacement Surgery: A Multicenter, Noninferiority, Randomized Clinical Trial Comparing a Home Exercise Program With Usual Outpatient Care. Arthritis Care & Research. 2015 Feb;67(2):196–202.

64. Hardt S, Schulz MRG, Pfitzner T, Wassilew G, Horstmann H, Liodakis E, et al. Improved early outcome after TKA through an app-based active muscle training programme—a randomized-controlled trial. Knee Surg Sports Traumatol Arthrosc. 2018 Nov;26(11):3429–37.

65. Harmer AR, Naylor JM, Crosbie J, Russell T. Land-based versus water-based rehabilitation following total knee replacement: A randomized, single-blind trial. Arthritis & Rheumatism. 2009 Feb 15;61(2):184–91.

66. Heikkilä A, Sevander-Kreus N, Häkkinen A, Vuorenmaa M, Salo P, Konsta P, et al. Effect of total knee replacement surgery and postoperative 12 month home exercise program on gait parameters. Gait & Posture. 2017 Mar;53:92–7.

67. Hepperger C, Gföller P, Hoser C, Ulmer H, Fischer F, Schobersberger W, et al. The effects of a 3-month controlled hiking programme on the functional abilities of patients following total knee arthroplasty: a prospective, randomized trial. Knee Surg Sports Traumatol Arthrosc. 2017 Nov;25(11):3387–95.

68. Husby VS, Foss OA, Husby OS, Winther SB. Randomized controlled trial of maximal strength training vs. standard rehabilitation following total knee arthroplasty. Eur J Phys Rehabil Med [Internet]. 2018 Jun [cited 2025 May 15];54(3). Available from: https://www.minervamedica.it/index2.php?show=R33Y2018N03A0371

69. Jacksteit R, Stöckel T, Behrens M, Feldhege F, Bergschmidt P, Bader R, et al. Low-Load Unilateral and Bilateral Resistance Training to Restore Lower Limb Function in the Early Rehabilitation After Total Knee Arthroplasty: A Randomized Active-Controlled Clinical Trial. Front Med. 2021 Jun 22;8:628021.

70. Jakobsen TL, Kehlet H, Husted H, Petersen J, Bandholm T. Early Progressive Strength Training to Enhance Recovery After Fast-Track Total Knee Arthroplasty: A Randomized Controlled Trial. Arthritis Care & Research. 2014 Dec;66(12):1856–66.

71. Janhunen M, Katajapuu N, Paloneva J, Pamilo K, Oksanen A, Keemu H, et al. Effects of a home-based, exergaming intervention on physical function and pain after total knee replacement in older adults: a randomised controlled trial. BMJ Open Sport Exerc Med. 2023 Mar;9(1):e001416.

72. Jiao S, Feng Z, Dai T, Huang J, Liu R, Meng Q. High-Intensity Progressive Rehabilitation Versus Routine Rehabilitation After Total Knee Arthroplasty: A Randomized Controlled Trial. The Journal of Arthroplasty. 2024 Mar;39(3):665–671.e2.

73. Johnson AW, Myrer JW, Hunter I, Feland JB, Hopkins JT, Draper DO, et al. Whole-body vibration strengthening compared to traditional strengthening during physical therapy in individuals with total knee arthroplasty. Physiotherapy Theory and Practice. 2010 Jan;26(4):215–25.

74. Karaman A, Yuksel I, Kinikli GI, Caglar O. Do Pilates-based exercises following total knee arthroplasty improve postural control and quality of life? Physiotherapy Theory and Practice. 2017 Apr 3;33(4):289–95.

75. Kauppila AM, Kyllönen E, Ohtonen P, Hämäläinen M, Mikkonen P, Laine V, et al. Multidisciplinary rehabilitation after primary total knee arthroplasty: a randomized controlled study of its effects on functional capacity and quality of life. Clin Rehabil. 2010 May;24(5):398–411.

76. Kelly MA, Finley M, Lichtman SW, Hyland MR, Edeer AO. Comparative Analysis of High-Velocity Versus Low-Velocity Exercise on Outcomes After Total Knee Arthroplasty: A Randomized Clinical Trial. Journal of Geriatric Physical Therapy. 2016 Oct;39(4):178–89.

77. Kiraç Can E, Tomruk M, Gelecek N. BİLATERAL TOTAL DİZ PROTEZİ SONRASI ERKEN İLERLEYİCİ KAPALI KİNETİK ZİNCİR EGZERSİZLERİNİN STANDART EGZERSİZ PROGRAMINA GÖRE ETKİLERİ – RANDOMİZE KONTROLLÜ ÇALIŞMA. Türk Fizyoterapi ve Rehabilitasyon Dergisi. 2023 Apr 20;34(1):102–14.

78. Ko V, Naylor J, Harris I, Crosbie J, Yeo A, Mittal R. One-to-One Therapy Is Not Superior to Group or Home-Based Therapy After Total Knee Arthroplasty: A Randomized, Superiority Trial. Journal of Bone and Joint Surgery. 2013 Nov 6;95(21):1942–9.

79. Kramer JF, Speechley M, Bourne R, Rorabeck C, Vaz M. Comparison of Clinic- and Home-Based Rehabilitation Programs After Total Knee Arthroplasty. Clinical Orthopaedics & Related Research. 2003 May;410:225–34.

80. Larsen JB, Skou ST, Laursen M, Bruun NH, Arendt-Nielsen L, Madeleine P. Exercise and Pain Neuroscience Education for Patients With Chronic Pain After Total Knee Arthroplasty: A Randomized Clinical Trial. JAMA Netw Open. 2024 May 24;7(5):e2412179.

81. Lee HG, An J, Lee BH. The Effect of Progressive Dynamic Balance Training on Physical Function, The Ability to Balance and Quality of Life Among Elderly Women Who Underwent a Total Knee Arthroplasty: A Double-Blind Randomized Control Trial. IJERPH. 2021 Mar 3;18(5):2513.

82. Lenguerrand E, Artz N, Marques E, Sanderson E, Lewis K, Murray J, et al. Effect of Group-Based Outpatient Physical Therapy on Function After Total Knee Replacement: Results From a Multicenter Randomized Controlled Trial. Arthritis Care & Research. 2020 Jun;72(6):768–77.

83. Lenssen AF, Crijns YH, Waltjé EM, Van Steyn MJ, Geesink RJ, Van Den Brandt PA, et al. Efficiency of immediate postoperative inpatient physical therapy following total knee arthroplasty: an RCT. BMC Musculoskelet Disord. 2006 Dec;7(1):71.

84. Levine M, McElroy K, Stakich V, Cicco J. Comparing Conventional Physical Therapy Rehabilitation With Neuromuscular Electrical Stimulation After TKA. Orthopedics [Internet]. 2013 Mar [cited 2025 May 15];36(3). Available from: https://journals.healio.com/doi/10.3928/01477447-20130222-20

85. Li L, Cheng S, Wang G, Duan G, Zhang Y. Tai chi chuan exercises improve functional outcomes and quality of life in patients with primary total knee arthroplasty due to knee osteoarthritis. Complementary Therapies in Clinical Practice. 2019 May;35:121–5.

86. Liao CD, Chiu YS, Ku JW, Huang SW, Liou TH. Effects of Elastic Resistance Exercise on Postoperative Outcomes Linked to the ICF Core Sets for Osteoarthritis after Total Knee Replacement in Overweight and Obese Older Women with Sarcopenia Risk: A Randomized Controlled Trial. JCM. 2020 Jul 11;9(7):2194.

87. Liao CD, Liou TH, Huang YY, Huang YC. Effects of balance training on functional outcome after total knee replacement in patients with knee osteoarthritis: a randomized controlled trial. Clin Rehabil. 2013 Aug;27(8):697–709.

88. Liebs TR, Herzberg W, Rüther W, Haasters J, Russlies M, Hassenpflug J. Ergometer Cycling After Hip or Knee Replacement Surgery: A Randomized Controlled Trial. The Journal of Bone and Joint Surgery-American Volume. 2010 Apr;92(4):814–22.

89. Liebs TR, Herzberg W, Rüther W, Haasters J, Russlies M, Hassenpflug J. Multicenter Randomized Controlled Trial Comparing Early Versus Late Aquatic Therapy After Total Hip or Knee Arthroplasty. Archives of Physical Medicine and Rehabilitation. 2012 Feb;93(2):192–9.

90. Madsen M, Larsen K, Madsen IK, Søe H, Hansen TB. Late group-based rehabilitation has no advantages compared with supervised home-exercises after total knee arthroplasty. Dan Med J. 2013 Apr;(60(4):A4607).

91. Maeda T, Sasaki E, Kasai T, Igarashi S, Wakai Y, Sasaki T, et al. Therapeutic effect of knee extension exercise with single-joint hybrid assistive limb following total knee arthroplasty: a prospective, randomized controlled trial. Sci Rep. 2024 Feb 16;14(1):3889.

92. Mau-Moeller A, Behrens M, Finze S, Bruhn S, Bader R, Mittelmeier W. The effect of continuous passive motion and sling exercise training on clinical and functional outcomes following total knee arthroplasty: a randomized active-controlled clinical study. Health Qual Life Outcomes. 2014;12(1):68.

93. Moffet H, Collet JP, Shapiro SH, Paradis G, Marquis F, Roy L. Effectiveness of intensive rehabilitation on functional ability and quality of life after first total knee arthroplasty: a single-blind randomized controlled trial. Archives of Physical Medicine and Rehabilitation. 2004 Apr;85(4):546–56.

94. Moffet H, Tousignant M, Nadeau S, Mérette C, Boissy P, Corriveau H, et al. In-Home Telerehabilitation Compared with Face-to-Face Rehabilitation After Total Knee Arthroplasty: A Noninferiority Randomized Controlled Trial. The Journal of Bone and Joint Surgery-American Volume. 2015 Jul;97(14):1129–41.

95. Monticone M, Ferrante S, Rocca B, Salvaderi S, Fiorentini R, Restelli M, et al. Home-Based Functional Exercises Aimed at Managing Kinesiophobia Contribute to Improving Disability and Quality of Life of Patients Undergoing Total Knee Arthroplasty: A Randomized Controlled Trial. Archives of Physical Medicine and Rehabilitation. 2013 Feb;94(2):231–9.

96. Moutzouri M, Gleeson N, Coutts F, Tsepis E, Gliatis J. Early self-managed focal sensorimotor rehabilitative training enhances functional mobility and sensorimotor function in patients following total knee replacement: a controlled clinical trial. Clin Rehabil. 2018 Jul;32(7):888–98.

97. Núñez-Cortés R, López-Bueno L, López-Bueno R, Cuenca-Martínez F, Suso-Martí L, Silvestre A, et al. Acute Effects of In-Hospital Resistance Training on Clinical Outcomes in Patients Undergoing Total Knee Arthroplasty: A Randomized Controlled Trial. Am J Phys Med Rehabil. 2024 May;103(5):401–9.

98. Papotto BA, Mills T. Treatment of Severe Flexion Deficits Following Total Knee Arthroplasty: A Randomized Clinical Trial. Orthopaedic Nursing. 2012 Jan;31(1):29–34.

99. Park D, Kim J, Lee H. Effectiveness of Modified Quadriceps Femoris Muscle Setting Exercise for the Elderly in Early Rehabilitation after Total Knee Arthroplasty. J Phys Ther Sci. 2012;24(1):27–30.

100. Park KJ, Seo TB, Kim YP. Effects of proprioceptive neuromuscular facilitation and both sides up ball exercise on pain level, range of motion, muscle function after total knee arthroplasty. J Exerc Rehabil. 2024 Feb 21;20(1):17–23.

101. Pastore E Silva AL. Estudo comparativo entre dois métodos de reabilitação fisioterapêutica na artroplastia total do joelho: protocolo padrão do IOT vs. protocolo avançado. Fisioter Bras. 2016 Jun 30;16(2):100–6.

102. Piqueras M, Marco E, Coll M, Escalada F, Ballester A, Cinca C, et al. Effectiveness of an interactive virtual telerehabilitation system in patients after total knee arthoplasty: A randomized controlled trial. J Rehabil Med. 2013;45(4):392–6.

103. Rahmann AE, Brauer SG, Nitz JC. A Specific Inpatient Aquatic Physiotherapy Program Improves Strength After Total Hip or Knee Replacement Surgery: A Randomized Controlled Trial. Archives of Physical Medicine and Rehabilitation. 2009 May;90(5):745–55.

104. Rajan R, Pack Y, Jackson H, Gillies C, Asirvatham R. No need for outpatient physiotherapy following total knee arthroplastyA randomized trial of 120 patients. Acta Orthopaedica Scandinavica. 2004 Jan;75(1):71–3.

105. Russell TG, Buttrum P, Wootton R, Jull GA. Low-bandwidth telerehabilitation for patients who have undergone total knee replacement: Preliminary results. J Telemed Telecare. 2003 Dec;9(2_suppl):44–7.

106. Russell TG, Buttrum P, Wootton R, Jull GA. Internet-Based Outpatient Telerehabilitation for Patients Following Total Knee Arthroplasty: A Randomized Controlled Trial. The Journal of Bone and Joint Surgery-American Volume. 2011 Jan;93(2):113–20.

107. Sattler LN, Hing WA, Vertullo CJ. Pedaling-Based Protocol Superior to a 10-Exercise, Non-Pedaling Protocol for Postoperative Rehabilitation After Total Knee Replacement: A Randomized Controlled Trial. The Journal of Bone and Joint Surgery. 2019 Apr 17;101(8):688–95.

108. Schache MB, McClelland JA, Webster KE. Incorporating hip abductor strengthening exercises into a rehabilitation program did not improve outcomes in people following total knee arthroplasty: a randomised trial. Journal of Physiotherapy. 2019 Jul;65(3):136–43.

109. Schulz M, Krohne B, Röder W, Sander K. Randomized, prospective, monocentric study to compare the outcome of continuous passive motion and controlled active motion after total knee arthroplasty. THC. 2018 Jun 29;26(3):499–506.

110. Shabbir M, Umar B, Ehsan S, Munir S, Bunin U, Sarfraz K. Comparison of functional training and strength training in improving knee extension lag after first four weeks of total knee replacement. 2017 [cited 2025 May 15]; Available from: https://www.biomedres.info/abstract/comparison-of-functional-training-and-strength-training-in-improving-knee-extension-lag-after-first-four-weeks-of-total-knee-repla-7757.html

111. Suh MJ, Kim BR, Kim SR, Han EY, Lee SY. Effects of Early Combined Eccentric-Concentric Versus Concentric Resistance Training Following Total Knee Arthroplasty. Ann Rehabil Med. 2017;41(5):816.

112. Tanaka R, Hayashizaki T, Taniguchi R, Kobayashi J, Umehara T. Effect of an intensive functional rehabilitation program on the recovery of activities of daily living after total knee arthroplasty: A multicenter, randomized, controlled trial. Journal of Orthopaedic Science. 2020 Mar;25(2):285–90.

113. Tanaka Y, Oka H, Nakayama S, Ueno T, Matsudaira K, Miura T, et al. Improvement of walking ability during postoperative rehabilitation with the hybrid assistive limb after total knee arthroplasty: A randomized controlled study. SAGE Open Medicine. 2017 Jan 1;5:2050312117712888.

114. Teissier V, Leclercq R, Schiano-Lomoriello S, Nizard R, Portier H. Does eccentric-concentric resistance training improve early functional outcomes compared to concentric resistance training after total knee arthroplasty? Gait & Posture. 2020 Jun;79:145–51.

115. Tousignant M, Moffet H, Boissy P, Corriveau H, Cabana F, Marquis F. A randomized controlled trial of home telerehabilitation for post-knee arthroplasty. J Telemed Telecare. 2011 Jun;17(4):195–8.

116. Trudelle-Jackson E, Hines E, Medley A, Thompson M. Exploration of Habitual Walking Behavior and Home-Based Muscle Power Training in Individuals With Total Knee Arthroplasty. Journal of Physical Activity and Health. 2020 Mar 1;17(3):331–8.

117. Tsukada Y, Matsuse H, Shinozaki N, Takano Y, Nago T, Shiba N. Combined Application of Electrically Stimulated Antagonist Muscle Contraction and Volitional Muscle Contraction Prevents Muscle Strength Weakness and Promotes Physical Function Recovery After Total Knee Arthroplasty: A Randomized Controlled Trial. Kurume Med J. 2018 Dec 31;65(4):145–54.

118. Unver B, Bakirhan S, Karatosun V. Does a weight-training exercise programme given to patients four or more years after total knee arthroplasty improve mobility: A randomized controlled trial. Archives of Gerontology and Geriatrics. 2016 May;64:45–50.

119. Valtonen A, Pöyhönen T, Sipilä S, Heinonen A. Effects of Aquatic Resistance Training on Mobility Limitation and Lower-Limb Impairments After Knee Replacement. Archives of Physical Medicine and Rehabilitation. 2010 Jun;91(6):833–9.

120. Vuorenmaa M, Ylinen J, Piitulainen K, Salo P, Kautiainen H, Pesola M, et al. Efficacy of a 12-month, monitored home exercise programme compared with normal care commencing 2 months after total knee arthroplasty: A randomized controlled trial. J Rehabil Med. 2014;46(2):166–72.

121. Warner S, Ahmad A, Afzal MW, Khan S, Aslam MM, Gillani SA. Comparison of routine physical therapy exercises with and without core stability exercises in total knee replacement patients. Rawal Medical Journal. 2020 Dec 17;45(4):842–842.

122. Xu T, Yang D, Liu K, Gao Q, Lu H, Qiao Y, et al. Efficacy and safety of a self-developed home-based enhanced knee flexion exercise program compared with standard supervised physiotherapy to improve mobility and quality of life after total knee arthroplasty: a randomized control study. J Orthop Surg Res. 2021 Dec;16(1):382.

123. Yang TH, Yeh WL, Chen HY, Chen YF, Ni KC, Lee KH. Compare the Traditional Chinese Medicine Manipulation With Rehabilitation on In-Patients After Total Knee Arthroplasty. The Journal of Arthroplasty. 2013 Jun;28(6):954–9.

124. Yousefian Molla R, Sadeghi H, Kahlaee AH. The Effect of Early Progressive Resistive Exercise Therapy on Balance Control of Patients With Total Knee Arthroplasty: A Randomized Controlled Trial. Topics in Geriatric Rehabilitation. 2017 Oct;33(4):286–94.

125. Zietek P, Zietek J, Szczypior K, Safranow K. Effect of adding one 15-minute-walk on the day of surgery to fast-track rehabilitation after total knee arthroplasty: a randomized, single-blind study. Eur J Phys Rehabil Med. 2015 Jun;51(3):245–52.

126. Großhennig A, Thomas NH, Brannath W, Koch A. How to avoid concerns with the interpretation of two primary endpoints if significant superiority in one is sufficient for formal proof of efficacy. Pharmaceutical Statistics. 2023;22(5):836–45.

127. Pinto RZ, Elkins MR, Moseley AM, Sherrington C, Herbert RD, Maher CG, et al. Many randomized trials of physical therapy interventions are not adequately registered: a survey of 200 published trials. Phys Ther. 2013 Mar;93(3):299–309.

128. Arienti C, Armijo-Olivo S, Ferriero G, Feys P, Hoogeboom T, Kiekens C, et al. The influence of bias in randomized controlled trials on rehabilitation intervention effect estimates: what we have learned from meta-epidemiological studies. Eur J Phys Rehabil Med. 2023 Dec 12;60(1):135–44.

129. Innocenti T, Giagio S, Salvioli S, Feller D, Minnucci S, Brindisino F, et al. Completeness of Reporting Is Suboptimal in Randomized Controlled Trials Published in Rehabilitation Journals, With Trials With Low Risk of Bias Displaying Better Reporting: A Meta-research Study. Archives of Physical Medicine and Rehabilitation. 2022 Sep;103(9):1839–47.

130. Slade SC, Dionne CE, Underwood M, Buchbinder R, Beck B, Bennell K, et al. Consensus on Exercise Reporting Template (CERT): Modified Delphi Study. Phys Ther. 2016 Oct;96(10):1514–24.

131. Hoffmann TC, Glasziou PP, Boutron I, Milne R, Perera R, Moher D, et al. Better reporting of interventions: template for intervention description and replication (TIDieR) checklist and guide. BMJ. 2014 Mar 7;348:g1687.

132. Moher D, Hopewell S, Schulz KF, Montori V, Gøtzsche PC, Devereaux PJ, et al. CONSORT 2010 explanation and elaboration: Updated guidelines for reporting parallel group randomised trials. International Journal of Surgery. 2012;10(1):28–55.

