## Appendix 2 for "Registration and reporting characteristics of trials investigating exercise therapy following total knee arthroplasty: A systematic review"

**Cochrane RCT search filters:**

| Table x The Cochrane Highly Sensitive Search Strategy for identifying randomized trials in MEDLINE, PubMed format | |
|---|---|
| #1 | randomized controlled trial [pt] |
| #2 | controlled clinical trial [pt] |
| #3 | randomized [tiab] |
| #4 | placebo [tiab] |
| #5 | randomly [tiab] |
| #6 | trial [tiab] |
| #7 | groups [tiab] |
| #8 | #1 OR #2 OR #3 OR #4 OR #5 OR #6 OR #7 |

| Table x The Cochrane Highly Sensitive Search Strategy for identifying randomized trials in MEDLINE, Ovid format | |
|---|---|
| #1 | randomized controlled trial.pt. |
| #2 | controlled clinical trial.pt. |
| #3 | randomized.ab. |
| #4 | placebo.ab. |
| #5 | randomly.ab. |
| #6 | trial.ab. |
| #7 | groups.ab. |
| #8 | #1 OR #2 OR #3 OR #4 OR #5 OR #6 OR #7 |

**Pubmed/MEDLINE full search-string:**

("randomized controlled trial"[Publication Type] OR "controlled clinical trial"[Publication Type] OR "randomized"[Title/Abstract] OR "placebo"[Title/Abstract] OR "randomly"[Title/Abstract] OR "trial"[Title/Abstract] OR "groups"[Title/Abstract]) AND ("knee arthroplast*"[Title/Abstract] OR "total knee arthroplast*"[Title/Abstract] OR "knee replacement*"[Title/Abstract] OR "total knee replacement*"[Title/Abstract] OR "arthroplasty, replacement, knee"[MeSH Terms]) AND ("exercise"[MeSH Terms] OR "exercise*"[Title/Abstract] OR "Resistance training"[Title/Abstract] OR "Resistance training"[MeSH Terms] OR "exercise therap*"[Title/Abstract] OR "Exercise therapy"[MeSH Terms] OR "rehabilitation*"[Title/Abstract] OR "rehabilitation"[MeSH Terms] OR "physical therap*"[Title/Abstract] OR "Physical therapy modalities"[MeSH Terms] OR "physiotherap*"[Title/Abstract]).

**EMBase via OVID**

(crossover procedure OR double-blind procedure OR randomized controlled trial OR single-blind procedure).hw. OR (random$ OR factorial$ OR crossover$ OR cross over$ OR placebo$ OR doubl$ blind$ OR singl$ blind$ OR assign$ OR allocate$ OR volunteer$).hw,ab,ti.

(Knee arthroplasty$ OR Total knee arthroplasty$ OR Knee replacement OR Total knee replacement$).ab,ti. OR exp Knee arthroplasty/

(Exercise$ OR Resistance training OR Exercise therap$ OR Rehabilitation$ OR Physical therap$ OR Physiotherap$).ti,ab. OR exp resistance training/ OR exp kinesiotherapy/ OR exp Rehabilitation/ OR exp Physiotherapy/ OR exp exercise/

**CiNAHL RCT filter:**

Cochrane CiNAHL plus filter, animal trials portion omitted (Glanville et al. 2019)

S1 MH randomized controlled trials

S2 MH double-blind studies

S3 MH single-blind studies

S4 MH random assignment

S5 MH pretest-posttest design

S6 MH cluster sample

S7 TI (randomised OR randomized)

S8 AB (random*)

S9 TI (trial)

S10 MH (sample size) AND AB (assigned OR allocated OR control)

S11 MH (placebos)

S12 PT (randomized controlled trial)

S13 AB (control W5 group)

S14 (MH “crossover design”) OR (MH “comparative studies”)

S15 AB (cluster W3 RCT)

**CINAHL via EBSCO**

| | |
|---|---|
| S33 | S6 AND S17 AND S32 |
| S32 | S18 OR S19 OR S20 OR S21 OR S22 OR S23 OR S24 OR S25 OR S26 OR S27 OR S28 OR S29 OR S30 OR S31 |
| S31 | AB (cluster W3 RCT) |
| S30 | (MH “crossover design”) OR (MH “comparative studies”) |
| S29 | AB (control W5 group) |
| S28 | PT randomized controlled trial |
| S27 | (MH "Placebos") |
| S26 | (MH "sample size") AND AB (assigned OR allocated OR control) |
| S25 | TI (trial) |
| S24 | AB (random*) |
| S23 | TI (randomised OR randomized) |

| | |
|---|---|
| S22 | (MH "Cluster Sample+") |
| S21 | (MH "Pretest-Posttest Design+") |
| S20 | (MH "Random Assignment") |
| S19 | (MH "Double-Blind Studies") OR (MH "Single-Blind Studies") |
| S18 | (MH "Randomized Controlled Trials+") |
| S17 | S7 OR S8 OR S9 OR S10 OR S11 OR S12 OR S13 OR S14 OR S15 OR S16 |
| S16 | TI physical therap* OR AB physical therap* OR TI physiotherap* OR AB physiotherap* |
| S15 | (MH "Physical Therapy+") |
| S14 | TI rehabilitation* OR AB rehabilitation* |
| S13 | (MH "Rehabilitation+") |
| S12 | TI exercise therapy* OR AB exercise therapy* |
| S11 | (MH "Therapeutic Exercise+") |
| S10 | TI resistance training* OR AB resistance training* |
| S9 | (MH "resistance training+") |
| S8 | TI exercise* OR AB exercise* |
| S7 | (MH "exercise+") |
| S6 | S1 OR S2 OR S3 OR S4 OR S5 |
| S5 | TI Total knee replacement* OR AB Total knee replacement* |
| S4 | TI Knee replacement* OR AB Knee replacement* |
| S3 | (MH "Arthroplasty, Replacement, Knee+") |
| S2 | TI Total knee arthroplast* OR AB Total knee arthroplast* |
| S1 | TI Knee arthroplast* OR AB Knee arthroplast* |

**Cochrane Central Register of Randomized Controlled Trials (CENTRAL)**

| ID | Search | Hits |
|---|---|---|
| #1 | (Knee arthroplast*):ti,ab,kw | |

#2 (Total knee arthroplast*):ti,ab,kw

#3 (Knee replacement?):ti,ab,kw

#4 (Total knee replacement?):ti,ab,kw

#5 MeSH descriptor: [Arthroplasty, Replacement, Knee] explode all trees

#6 #1 OR #2 OR #3 OR #4 OR #5

#7 MeSH descriptor: [Exercise] explode all trees

#8 (exercise? OR exercise*):ti,ab,kw

#9 (resistance training):ti,ab,kw

#10 MeSH descriptor: [Resistance Training] explode all trees

#11 (exercise therap* OR exercise therap?):ti,ab,kw

#12 MeSH descriptor: [Exercise Therapy] explode all trees

#13 (rehabilitation* OR rehabilitation?):ti,ab,kw

#14 MeSH descriptor: [Rehabilitation] explode all trees

#15 (physical therap* OR physical therap?):ti,ab,kw

#16 MeSH descriptor: [Physical Therapy Modalities] explode all trees

#17 (physiotherap* OR physiotherap?):ti,ab,kw

#18 #7 OR #8 OR #9 OR #10 OR #11 OR #12 OR #13 OR #14 OR #15 OR #16 OR #17

#19 #6 AND #18
