## Appendix 3 for "Registration and reporting characteristics of trials investigating exercise therapy following total knee arthroplasty: A systematic review"

| Exclusion reason | No. | Reference |
|---|---|---|
| Same exercise therapy | 127 | (1–127) |
| Conference abstracts, commentaries, supplementary documents, and protocols | 74 | (128–201) |
| Nonrandomised trial design | 35 | (202–236) |
| Population already included | 35 | (237–271) |
| Mixed orthopaedic population | 29 | (272–300) |
| Pilot / Feasibility study | 22 | (301–322) |
| Published before 2000 | 20 | (323–342) |
| Population with UKA | 3 | (343–345) |
| Retracted | 1 | (346) |

*Appendix 3 - Reasons and citations to reports excluded during full text screening*

**References**


1. Alaca N, Atalay A, Güven Z. Comparison of the long-term effectiveness of progressive neuromuscular facilitation and continuous passive motion therapies after total knee arthroplasty. J Phys Ther Sci. 2015;27(11):3377–80.

2. Alghadir A, Iqbal ZA, Anwer S. Comparison of the effect of pre- and post-operative physical therapy versus post-operative physical therapy alone on pain and recovery of function after total knee arthroplasty. J Phys Ther Sci. 2016;28(10):2754–8.

3. Alkire M, Swank M. Use of inpatient continuous passive motion versus no CPM in computer-assisted total knee arthroplasty. 2010;29(1):36-40.

4. Ask S, Lindmark B, Johansson A. [Assessment of continuous passive motion (CPM) in rehabilitation after total knee arthroplasty]. Nordisk Fysioterapi. 2003;7(2):29–39.

5. Avramidis K, Karachalios T, Popotonasios K, Sacorafas D, Papathanasiades A, Malizos K. Does electric stimulation of the vastus medialis muscle influence rehabilitation after total knee replacement? 2011;34(3):175.

6. Avramidis K, Strike P, Taylor P, Swain I. Effectiveness of electric stimulation of the vastus medialis muscle in the rehabilitation of patients after total knee arthroplasty. 2003;84(12):1850-1853.

7. Baloch N, Zubairi AJ, Rashid RH, Hashmi PM, Lakdawala RH. Effect of continuous passive motion on knee flexion range of motion after total knee arthroplasty. J Pak Med Assoc. 2015;65(11 Suppl 3):S32-34.

8. Beaupré L, Davies D, Jones C, Cinats J. Exercise combined with continuous passive motion or slider board therapy compared with exercise only: a randomized controlled trial of patients following total knee arthroplasty. 2001;81(4):1029-1037.

9. Bhatia S., Karvannan H., Prem V. The effect of bio psychosocial model of rehabilitation on pain and quality of life after total knee replacement: A randomized controlled trial. J Arthrosc Jt Surg. 2020;7(4):177–83.

10. Błaszczak E, Franek A, Taradaj J, Widuchowski J, Klimczak J. Assessment of the efficacy and safety of low frequency, low intensity magnetic fields in patients after knee endoprosthesis plasty. Part 2: a clinical study. Bioelectromagnetics. 2009;30(2):152–8.

11. Briones-Cantero M, Fernández-de-Las-Peñas C, Lluch-Girbés E, Osuna-Pérez MC, Navarro-Santana MJ, Plaza-Manzano G, et al. Effects of Adding Motor Imagery to Early Physical Therapy in Patients with Knee Osteoarthritis who Had Received Total Knee Arthroplasty: A Randomized Clinical Trial. Pain Med. 2020;21(12):3548–55.

12. Bruun-Olsen V, Heiberg K, Mengshoel A. Continuous passive motion as an adjunct to active exercises in early rehabilitation following total knee arthroplasty - a randomized controlled trial. 2009;31(4):277-283.

13. Casaña J, Calatayud J, Ezzatvar Y, Vinstrup J, Benítez J, Andersen L. Preoperative high-intensity strength training improves postural control after TKA: randomized-controlled trial. 2019;27(4):1057-1066.

14. Chen B, Zimmerman J, Soulen L, DeLisa J. Continuous passive motion after total knee arthroplasty: a prospective study. 2000;79(5):421-426.

15. Chen G, Gu R, Xu D. The application of electroacupuncture to postoperative rehabilitation of total knee replacement. 2012;32(4):309-312.

16. Chen H, Du SS, Fan LJ, Jiang J, Liang XY, Zhen P, et al. Effects of combination therapy on the movement function of lower extremity after total knee arthroplasty in the elderly. 2017;21(31):4939-4944.

17. Chen M, Li P, Lin F. Influence of structured telephone follow-up on patient compliance with rehabilitation after total knee arthroplasty. 2016;10:257-264.

18. Chen Q, Huang S, Chen X, Feng L, Zhu X. Clinical efficacy of multi-pattern detumescence after total knee arthroplasty treated with acupoint massage and mild moxibustion. 2016;36(5):471-475.

19. Cheng HJ, Duh YJ, Wu MH, Pan CC, Chen KH, Lee CH. Continuous passive motion: effect on post-total knee arthroplasty elders at an orthopedic ward in central Taiwan. Journal of Nursing & Healthcare Research. 2012;8(2):166–166.

20. Christiansen CL, Kline PW, Anderson CB, Melanson EL, Sullivan WJ, Richardson VL, et al. Optimizing Total Knee Arthroplasty Rehabilitation with Telehealth Physical Activity Behavior Change Intervention: A Randomized Clinical Trial. Phys Ther. 2024;pzae088.

21. Christiansen M, Thoma L, Master H, Voinier D, Schmitt L, Ziegler M, et al. The feasibility and preliminary outcomes of a physical therapist-administered physical activity intervention after total knee replacement. 2019; Available from: https://www.cochranelibrary.com/central/doi/10.1002/central/CN-01937514/full

22. Ciani O, Pascarelli NA, Giannitti C, Galeazzi M, Meregaglia M, Fattore G, et al. Mud-Bath Therapy in Addition to Usual Care in Bilateral Knee Osteoarthritis: An Economic Evaluation Alongside a Randomized Controlled Trial. Arthritis Care & Research. 2017;69(7):966–72.

23. Cien A., Riggle P.K., Brazier B.G., Popovich J.M., Crawford S., Cochran J.M. Examining the use of the pressure modulated knee rehabilitation machine (PMKR) with traditional physical therapy versus traditional physical therapy alone following total knee arthroplasty: A randomized control study. Curr Orthop Pract. 2021;32(1):15–22.

24. Coleman G, White DK, Thoma LM, Mathews D, Christiansen MB, Schmitt LA, et al. Does a physical therapist-administered physical activity intervention reduce sedentary time after total knee replacement: An exploratory study? Musculoskeletal Care. 2021;19(1):142–5.

25. Davies D, Johnston D, Beaupre L, Lier D. Effect of adjunctive range-of-motion therapy after primary total knee arthroplasty on the use of health services after hospital discharge. 2003;46(1):30-36.

26. Debbi E, Bernfeld B, Herman A, Salai M, Laufer Y, Wolf A. A Biomechanical Foot-Worn Device Improves Total Knee Arthroplasty Outcomes. 2019;34(1):47-55.

27. Demircioglu DT, Paker N, Erbil E, Bugdayci D, Emre TY. The effect of neuromuscular electrical stimulation on functional status and quality of life after knee arthroplasty: a randomized controlled study. J Phys Ther Sci. 2015;27(8):2501–6.

28. Denis M, Moffet H, Caron F, Ouellet D, Paquet J, Nolet L. Effectiveness of continuous passive motion and conventional physical therapy after total knee arthroplasty: a randomized clinical trial. 2006;86(2):174-185.

29. Donec V, Kriščiūnas A. The effectiveness of Kinesio Taping® after total knee replacement in early postoperative rehabilitation period. A randomized controlled trial. 2014;50(4):363-371.

30. Duong V, Robbins SR, Dennis S, Venkatesha V, Ferreira ML, Hunter DJ. Combined Digital Interventions for Pain Reduction in Patients Undergoing Knee Replacement: A Randomized Clinical Trial. JAMA Netw Open. 2023;6(9):e2333172.

31. Ersözlü S, Sahin O, Ozgür A, Tuncay I. The effects of two different continuous passive motion protocols on knee range of motion after total knee arthroplasty: a prospective analysis. 2009;43(5):412-418.

32. Eymir M, Unver B, Karatosun V. Relaxation exercise therapy improves pain, muscle strength, and kinesiophobia following total knee arthroplasty in the short term: a randomized controlled trial. Knee Surg Sports Traumatol Arthrosc. 2022;30(8):2776–85.

33. Fitz W, Shukla P, Li L, Scott R. Early regain of function and proprioceptive improvement following knee arthroplasty. 2018;6(6):523-531.

34. Fortuno Godes J, Martin Baranera M, Kadar García E, Redondo Parra I, Gallardo Perez B. Decrease in pain and weight loss during physiotherapy treatment in patients operated on for knee prosthesis. Fisioterapia. 2010;32(1):11–6.

35. Gianola S, Stucovitz E, Castellini G, Mascali M, Vanni F, Tramacere I, et al. Effects of early virtual reality-based rehabilitation in patients with total knee arthroplasty: A randomized controlled trial. Medicine (Baltimore). 2020;99(7):e19136.

36. Giaquinto S, Ciotola E, Dall'Armi V, Margutti F. Hydrotherapy after total knee arthroplasty. A follow-up study. 2010;51(1):59-63.

37. Gür O, Başar S. The effect of virtual reality on pain, kinesiophobia and function in total knee arthroplasty patients: A randomized controlled trial. Knee. 2023;45:187–97.

38. Hardwick M, Pulido P, Adelson W. Nursing intervention using healing touch in bilateral total knee arthroplasty. 2012;31(1):5-11.

39. Hasubhai PZ, D. BD, A. TR. Effectiveness of Conventional Physiotherapy along with Continuous Passive Motion after Total Knee Arthroplasty. Indian Journal of Physiotherapy & Occupational Therapy. 2017;11(4):195–200.

40. Herbold J, Bonistall K, Blackburn M, Agolli J, Gaston S, Gross C, et al. Randomized controlled trial of the effectiveness of continuous passive motion after total knee replacement. 2014;95(7):1240-1245.

41. Huang D, Peng Y, Su P, Ye W, Liang A. The effect of continuous passive motion after total knee arthroplasty on joint function. 2003;7(11):1661-1662.

42. Huang P, He J, Zhang Y. The mobile application of patient management in education and follow-up for patients following total knee arthroplasty. 2017;97(20):1592-1595.

43. Jacofsky D, Kocisky S, Dixon D, Jacofsky M. Secure tracks device improves functional recovery and pain after total knee arthroplasty: a prospective, randomized, pilot study. 2010;20:357-361.

44. Jain S, Wasnik S, Mittal A, Hegde C. Outcome of subvastus approach in elderly nonobese patients undergoing bilateral simultaneous total knee arthroplasty: A randomized controlled study. Indian J Orthop. 2013;47(1):45–9.

45. Jarvis S, Johnson-Wo A, Onstot B, Bhowmik-Stoker M, Shrader M, Jacofsky M, et al. Differences between standard and minimally invasive parapatellar surgical approaches for total knee arthroplasty in the tasks of sitting and standing. 2013;26(4):249-256.

46. Jenkins C, Barker K, Pandit H, Dodd C, Murray D. After partial knee replacement, patients can kneel, but they need to be taught to do so: a single-blind randomized controlled trial. 2008;88(9):1012-1021.

47. Jin C, Feng Y, Ni Y, Shan Z. Virtual reality intervention in postoperative rehabilitation after total knee arthroplasty: a prospective and randomized controlled clinical trial. 2018;11(6):6119-6124.

48. Joshi R, White P, Murray-Weir M, Alexiades M, Sculco T, Ranawat A. Prospective Randomized Trial of the Efficacy of Continuous Passive Motion Post Total Knee Arthroplasty: experience of the Hospital for Special Surgery. 2015;30(12):2364-2369.

49. Ju CJ, Zhou X, Dong CC, Lin LQ, Liu HN, Hou Y. [Clinical observation of warm moxibustion therapy to improve quadriceps weakness after total knee arthroplasty]. Zhongguo Zhen Jiu. 2019;39(3):276–9.

50. Kang K, Geng Q, Xu H, Zheng X, Dong J, Li T, et al. Clinical study of a new wearable device for rehabilitation after total knee arthroplasty. 2018;98(15):1162-1165.

51. Karaduz E.G., Demirbas R.E., Yagcioglu A., Hantal S.B. Sensorimotor versus core stabilization home exercise programs following total knee arthroplasty: a randomized controlled trial. Adv Rehabil. 2024;38(1):20–34.

52. Kim SM, Kim SR, Lee YK, Kim BR, Han EY. The effect of mechanical massage on early outcome after total knee arthroplasty: a pilot study. J Phys Ther Sci. 2015;27(11):3413–6.

53. Labraca N, Castro-Sánchez A, Matarán-Peñarrocha G, Arroyo-Morales M, Sánchez-Joya Mdel M, Moreno-Lorenzo C. Benefits of starting rehabilitation within 24 hours of primary total knee arthroplasty: randomized clinical trial. 2011;25(6):557-566.

54. Larsen K, Sørensen O, Hansen T, Thomsen P, Søballe K. Accelerated perioperative care and rehabilitation intervention for hip and knee replacement is effective: a randomized clinical trial involving 87 patients with 3 months of follow-up. 2008;79(2):149-159.

55. Laskin R, Maruyama Y, Villaneuva M, Bourne R. Deep-dish congruent tibial component use in total knee arthroplasty: a randomized prospective study. 2000;(380):36-44.

56. Lee JY, Kim JH, Lee BH. Effect of Dynamic Balance Exercises Based on Visual Feedback on Physical Function, Balance Ability, and Depression in Women after Bilateral Total Knee Arthroplasty: A Randomized Controlled Trial. Int J Environ Res Public Health. 2020;17(9).

57. Lenssen A, Bie R, Bulstra S, Steyn Mja. Continuous passive motion (CPM) in rehabilitation following total knee arthroplasty: a randomised controlled trial. 2003;8:123-129.

58. Lenssen T, van Steyn M, Crijns Y, Waltjé E, Roox G, Geesink R, et al. Effectiveness of prolonged use of continuous passive motion (CPM), as an adjunct to physiotherapy, after total knee arthroplasty. 2008;9:60.

59. Leonard H. Live Music Therapy During Rehabilitation After Total Knee Arthroplasty: a Randomized Controlled Trial. 2019;56(1):61-89.

60. Li L, Wang Z, Yin MH, Li Q, Qi ZM. Effect of early gait training on the functional rehabilitation after total knee arthroplasty. 2017;21(27):4288-4293.

61. Li Z, Li B, Wang G, Wang K, Chen J, Liang Y, et al. Impact of enhanced recovery nursing combined with limb training on knee joint function and neurological function after total knee arthroplasty in patients with knee osteoarthritis. Am J Transl Res. 2021;13(6):6864–72.

62. Lin Y, Hu X, Cao Y, Wang X, Tong Y, Yao F, et al. The Role of 6-Minute Walk Test Guided by Impedance Cardiography in the Rehabilitation Following Knee Arthroplasty: A Randomized Controlled Trial. Front Cardiovasc Med. 2021;8:736208.

63. Liu M, Jia Y, Shi N. Application value of an integrated treatment model of orthopedic rehabilitation in patients undergoing total knee arthroplasty. 2018;11(9):9455-9461.

64. Liu P, Li L, Zhang Y, Li M, Kane K, Wang Y, et al. A comparison of two rehabilitation protocols after simultaneous bilateral total knee arthroplasty: a controlled, randomized study. 2009;37(3):746-756.

65. Liu W, Wu YL, Cong RJ, Fu PL, Li XH, Wu HS. Controlled active motion and continuous passive motion are beneficial to function rehabilitation after total knee arthroplasty. 2011;15(35):6509-6513.

66. Liu YJ, Xu HP, Zhao H. Self-management approaches following total knee arthroplasty. 2011;15(17):3057-3061.

67. Losina E, Collins J, Deshpande B, Smith S, Michl G, Usiskin I, et al. Financial Incentives and Health Coaching to Improve Physical Activity Following Total Knee Replacement: a Randomized Controlled Trial. 2018;70(5):732-740.

68. Luo SM, Liao YM, Chen L, Huang Y, Zhou L, Tang J. Effect of continuous passive motion on blood coagulation condition in patients with total knee joint replacement. 2012;16(44):8182-8185.

69. Machi A, Sztain J, Kormylo N, Madison S, Abramson W, Monahan A, et al. Discharge-readiness following tricompartment knee arthroplasty: adductor canal versus femoral continuous nerve blocks; a dual-center, randomized trial. 2015;40(5) (no pagination). Available from: https://www.cochranelibrary.com/central/doi/10.1002/central/CN-01171997/full

70. Marzetti E, Rabini A, Piccinini G, Piazzini DB, Vulpiani M Donec V, Kriscinas A. The effectiveness of Kinesio Taping(R) after total knee replacement in early postoperative rehabilitation period. A randomized controlled trial. European Journal of Physical & Rehabilitation Medicine. 2014;50(4):363–71.

71. Mat Eil Ismail M, Sharifudin M, Shokri A, Ab Rahman S. Preoperative physiotherapy and short-term functional outcomes of primary total knee arthroplasty. 2016;57(3):138-143.

72. McHugh G. The role of perhabilitation on the outcome of total knee arthroplasty: A randomized control trial. [Internet]. University College Dublin (Ireland); 2011. Available from: http://search.ebscohost.com/login.aspx?direct=true&db=rzh&AN=109865629&site=ehost-live

73. Mitchell C, Walker J, Walters S, Morgan A, Binns T, Mathers N. Costs and effectiveness of pre- and post-operative home physiotherapy for total knee replacement: randomized controlled trial. 2005;11(3):283-292.

74. Moretti B, Notarnicola A, Moretti L, Setti S, De Terlizzi F, Pesce V, et al. I-ONE therapy in patients undergoing total knee arthroplasty: a prospective, randomized and controlled study. 2012;13:88.

75. Moukarzel M, Di Rienzo F, Lahoud JC, Hoyek F, Collet C, Guillot A, et al. The therapeutic role of motor imagery during the acute phase after total knee arthroplasty: a pilot study. 2019;41(8):926-933.

76. Nigam A, Taylor D, Valeyeva Z. Non-invasive interactive neurostimulation (InterX™) reduces acute pain in patients following total knee replacement surgery: a randomised, controlled trial. 2011;6:45.

77. Nuevo M, Rodríguez-Rodríguez D, Jauregui R, Fabrellas N, Zabalegui A, Conti M, et al. Telerehabilitation following fast-track total knee arthroplasty is effective and safe: a randomized controlled trial with the ReHub® platform. Disabil Rehabil. 2024;46(12):2629–39.

78. Oh HT, Hwangbo G. The effects of proprioception exercise with and without visual feedback on the pain and balance in patients after total knee arthroplasty. J Phys Ther Sci. 2018;30(1):124–6.

79. Paravlic AH, Maffulli N, Kovač S, Pisot R. Home-based motor imagery intervention improves functional performance following total knee arthroplasty in the short term: a randomized controlled trial. J Orthop Surg Res. 2020;15(1):451.

80. Paravlic AH, Pisot R, Marusic U. Specific and general adaptations following motor imagery practice focused on muscle strength in total knee arthroplasty rehabilitation: A randomized controlled trial. PLoS One. 2019;14(8):e0221089.

81. Park SD, Song HS, Kim JY. The effect of action observation training on knee joint function and gait ability in total knee replacement patients. J Exerc Rehabil. 2014;10(3):168–71.

82. Park SA, Jeong Y. The Effect of a Multidimensional Home Rehabilitation Program for Post-Total Knee Arthroplasty Elderly Patients. Orthop Nurs. 2023;42(1):22–32.

83. Pereira LC, Jolles BM. The effect of end-of-range grade A+ knee mobilisation following acute primary total knee arthroplasty: A randomised controlled trial. International Journal of Therapy & Rehabilitation. 2015;22(12):583–91.

84. Petersen T, Hautopp H, Duus B, Juhl C. No effect of Acupuncture as adjunctive therapy for patients with total knee replacement: a randomized controlled trial. 2018;19(6):1280-1289.

85. Petterson S, Mizner R, Stevens J, Raisis L, Bodenstab A, Newcomb W, et al. Improved function from progressive strengthening interventions after total knee arthroplasty: a randomized clinical trial with an imbedded prospective cohort. 2009;61(2):174-183.

86. Pichonnaz C, Bassin J, Lécureux E, Christe G, Currat D, Aminian K, et al. Effect of Manual Lymphatic Drainage After Total Knee Arthroplasty: a Randomized Controlled Trial. 2016;97(5):674-682.

87. Pournajaf S, Goffredo M, Pellicciari L, Piscitelli D, Criscuolo S, Le Pera D, et al. Effect of balance training using virtual reality-based serious games in individuals with total knee replacement: A randomized controlled trial. Ann Phys Rehabil Med. 2022;65(6):101609.

88. Prvu Bettger J, Green CL, Holmes DN, Chokshi A, Mather RC 3rd, Hoch BT, et al. Effects of Virtual Exercise Rehabilitation In-Home Therapy Compared with Traditional Care After Total Knee Arthroplasty: VERITAS, a Randomized Controlled Trial. J Bone Joint Surg Am. 2020;102(2):101–9.

89. Pua YH, Yeo SJ, Clark RA, Tan BY, Haines T, Bettger JP, et al. Cost and outcomes of Hospital-based Usual cAre versus Tele-monitor self-directed Rehabilitation (HUATR) in patients with total knee arthroplasty: A randomized, controlled, non-inferiority trial. Osteoarthritis Cartilage. 2024;32(5):601–11.

90. Rakel B, Zimmerman M, Geasland K, Embree J, Clark C, Noiseux N, et al. Transcutaneous electrical nerve stimulation for the control of pain during rehabilitation after total knee arthroplasty: a randomized, blinded, placebo-controlled trial. 2014;155(12):2599-2611.

91. Rockstroh G, Schleicher W, Krummenauer F. Effectiveness of microcurrent therapy as a constituent of post-hospital rehabilitative treatment in patients after total knee alloarthroplasty - a randomized clinical trial. 2010;49(3):173-179.

92. Ródenas-Martínez S, Santos-Andrés JF, Abril-Boren C, Usabiaga-Bernal T, Abouh-Lais S, Aguilar-Naranjo JJ. Effectiveness of a pre-surgery rehabilitation program in total knee arthroplasty. Rehabilitacion. 2008;42(1):4–12.

93. Roig-Casasús S, Blasco JM, López-Bueno L, Blasco-Igual MC. Balance Training With a Dynamometric Platform Following Total Knee Replacement: A Randomized Controlled Trial. Journal of Geriatric Physical Therapy. 2018;41(4):204–9.

94. Russo L, Benedetti M, Mariani E, Roberti di Sarsina T, Zaffagnini S. The Videoinsight® Method: improving early results following total knee arthroplasty. 2017;25(9):2967-2971.

95. Sahin A, Agar A, Erturk C. The effect of telerehabilitation on early outcomes in patients undergoing primary total knee replacement: A prospective randomized study. Journal of Surgery & Medicine (JOSAM). 2022;6(2):139–43.

96. Sahin E, Akalin E, Bircan C, Karaoglan O, Tatari H, Alper S, et al. The effects of continuous passive motion on outcome in total knee arthroplasty. 2006;17(2):85-90.

97. Shanb AS, Youssef E. Effects of adding biofeedback training to active exercises after total knee arthroplasty. 2014;17(1). Available from: https://www.cochranelibrary.com/central/doi/10.1002/central/CN-01041112/full

98. Shim GY, Kim EH, Lee SJ, Chang CB, Lee YS, Lee JI, et al. Postoperative rehabilitation using a digital healthcare system in patients with total knee arthroplasty: a randomized controlled trial. Arch Orthop Trauma Surg. 2023;143(10):6361–70.

99. Sindhupakorn B., Numpaisal P.-O., Thienpratharn S., Jomkoh D. A home visit program versus a non-home visit program in total knee replacement patients: a randomized controlled trial. J Orthop Surg Res. 2019;14(1):405.

100. Sklempe Kokic I, Vuksanic M, Kokic T, Peric I, Duvnjak I. Effects of Electromyographic Biofeedback on Functional Recovery of Patients Two Months after Total Knee Arthroplasty: A Randomized Controlled Trial. J Clin Med. 2022;11(11).

101. Skoffer B, Maribo T, Mechlenburg I, Hansen P, Søballe K, Dalgas U. Efficacy of Preoperative Progressive Resistance Training on Postoperative Outcomes in Patients Undergoing Total Knee Arthroplasty. 2016;68(9):1239-1251.

102. Skou S, Roos E, Simonsen O, Laursen M, Rathleff M, Arendt-Nielsen L, et al. The effects of total knee replacement and non-surgical treatment on pain sensitization and clinical pain. 2016;20(10):1612-1621.

103. Skov Husted R, Wilquin L, Linding Jakobsen T, Holsgaard-Larsen A, Bandholm T. RAPID KNEE-EXTENSIONS TO INCREASE QUADRICEPS MUSCLE ACTIVITY IN PATIENTS WITH TOTAL KNEE ARTHROPLASTY: A RANDOMIZED CROSS-OVER STUDY. International Journal of Sports Physical Therapy. 2017;12(1):105–16.

104. Smith W, Zucker-Levin A, Mihalko W, Williams M, Loftin M, Gurney J. A Randomized Study of Exercise and Fitness Trackers in Obese Patients After Total Knee Arthroplasty. 2019;50(1):35-45.

105. Song LX, Yang L, Li Y, Lei FQ, Qin Y, Wang LH, et al. Influence of health education based on the transtheoretical model on kinesiophobia levels and rehabilitation outcomes in elderly patients undergoing total knee arthroplasty. Heliyon. 2024;10(12):e32445.

106. Spiegl C, Strutzenberger G, Schwameder H. EMG-Biofeedbacktraining bei Patienten mit Knieendoprothese. Zeitschrift fur Physiotherapeuten. 2018;70(3):87–94.

107. Stevens-Lapsley J, Balter J, Wolfe P, Eckhoff D, Kohrt W. Early neuromuscular electrical stimulation to improve quadriceps muscle strength after total knee arthroplasty: a randomized controlled trial. 2012;92(2):210-226.

108. Sueta D, Kaikita K, Okamoto N, Yamabe S, Ishii M, Arima Y, et al. Edoxaban Enhances Thromboprophylaxis by Physiotherapy After Total Knee Arthroplasty - The Randomized Controlled ESCORT-TKA Trial. 2018;82(2):524-531.

109. Szöts K, Konradsen H, Solgaard S, Østergaard B. Telephone Follow-Up by Nurse After Total Knee Arthroplasty: results of a Randomized Clinical Trial. 2016;35(6):411-420.

110. Tang J, Yang T, Xiong XJ, Chen L, Cheng ZY, Zhan FB, et al. Air wave pressure therapy in prevention of deep vein thrombosis of the lower extremity after total knee arthroplasty. 2013;17(52):8981-8986.

111. Timmers T, Janssen L, van der Weegen W, Das D, Marijnissen WJ, Hannink G, et al. The Effect of an App for Day-to-Day Postoperative Care Education on Patients With Total Knee Replacement: Randomized Controlled Trial. JMIR Mhealth Uhealth. 2019;7(10):e15323.

112. Torpil B, Kaya Ö. The Effectiveness of Client-Centered Intervention With Telerehabilitation Method After Total Knee Arthroplasty. OTJR (Thorofare N J). 2022;42(1):40–9.

113. Tripuraneni KR, Foran JRH, Munson NR, Racca NE, Carothers JT. A Smartwatch Paired With A Mobile Application Provides Postoperative Self-Directed Rehabilitation Without Compromising Total Knee Arthroplasty Outcomes: A Randomized Controlled Trial. J Arthroplasty. 2021;36(12):3888–93.

114. Trzeciak T., Richter M., Ruszkowski K. [Effectiveness of continuous passive motion after total knee replacement]. Chirurgia narzadow ruchu i ortopedia polska. 2011;76(6):345–9.

115. Tsang R, Tsang P, Ko C, Kong B, Lee W, Yip H. Effects of acupuncture and sham acupuncture in addition to physiotherapy in patients undergoing bilateral total knee arthroplasty--a randomized controlled trial. 2007;21(8):719-728.

116. Tugay N, Saricaoglu F, Satilmis T, Alpar U, Akarcali I, Citaker S, et al. Effects on the independence level in functional activities in the early postoperative period in patients with total knee arthroplasty. 2006;11(3):175-179.

117. Vaegter H, Handberg G, Emmeluth C, Graven-Nielsen T. Preoperative Hypoalgesia After Cold Pressor Test and Aerobic Exercise is Associated With Pain Relief 6 Months After Total Knee Replacement. 2017;33(6):475-484.

118. Valdes Vilches M., Fernandez Ferreras T., Serra Tarragon N., Bujedo Pertejo A., San Segundo Mozo R., Molins Roca J. Feedback and neuromuscular electrical stimulation during an early phase of a rehabilitation programe after total knee arthroplasty. Trauma (Spain). 2010;21(3):163–8.

119. Villafañe J, Isgrò M, Borsatti M, Berjano P, Pirali C, Negrini S. Effects of action observation treatment in recovery after total knee replacement: a prospective clinical trial. 2017;31(3):361-368.

120. Wang S, Xia J, Wei Y, Wu J, Huang G. Effect of the knee position during wound closure after total knee arthroplasty on early knee function recovery. 2014;9:79.

121. Witvrouw E, Bellemans J, Victor J. Manipulation under anaesthesia versus low stretch device in poor range of motion after TKA. 2013;21(12):2751-2758.

122. Wozniak-Czekierda W, Wozniak K, Hadamus A, Bialoszewski D. Use of Kinesiology Taping in Rehabilitation after Knee Arthroplasty: a Randomised Clinical Study. 2017;19(5):461-468.

123. Wylde V., Bertram W., Sanderson E., Noble S., Howells N., Peters T.J., et al. The STAR care pathway for patients with pain at 3 months after total knee replacement: a multicentre, pragmatic, randomised, controlled trial. Lancet Rheumat. 2022;4(3):e188–97.

124. Yim S, Min K, Lee Y, Kim H, Kang H. Efficacy of Physiotherapist after Total Knee Arthroplasty. 2009;21(4):258-264.

125. Yoshida Y, Ikuno K, Shomoto K. Comparison of the Effect of Sensory-Level and Conventional Motor-Level Neuromuscular Electrical Stimulations on Quadriceps Strength After Total Knee Arthroplasty: a Prospective Randomized Single-Blind Trial. 2017;(no pagination). Available from: https://www.cochranelibrary.com/central/doi/10.1002/central/CN-01394314/full

126. Zhao R, Cheng L, Zheng Q, Lv Y, Wang YM, Ni M, et al. A Smartphone Application-Based Remote Rehabilitation System for Post-Total Knee Arthroplasty Rehabilitation: A Randomized Controlled Trial. J Arthroplasty. 2024;39(3):575-581.e8.

127. Zhou R., Wu T., Huang L., Wang H., Lu S. Effectiveness of Inertial Measurement Unit Sensor-Based Feedback Assistance in Telerehabilitation of Patients with Diabetes after Total Knee Arthroplasty: A Randomized Controlled Trial. Telemed Rep. 2024;5(1):141–51.

128. Christiansen M, Thoma L, Master H, Mathews D, Schmitt L, Ziegler M, et al. 1-year outcomes from a novel physical therapist-administered physical activity intervention after total knee replacement: a pilot study. 2018;70:439-440.

129. Skou S, Roos E, Laursen M, Arendt-Nielsen L, Rasmussen S, Simonsen O, et al. 2-year cost-effectiveness of total knee replacement: results from the first randomized trial on total knee replacement in addition to non-surgical treatment. 2019;27:S297-.

130. Debbi E, Bernfeld B, Soudry M, Salai M, Laufer Y, Herman A, et al. A biomechanical therapy program for patients after total knee arthroplasty - A randomized controlled trial (preliminary results). 2014;22:S82-.

131. Voinier D., Master H., Thoma L.M., Brunette M., Jakiela J.T., Copson J., et al. A PHYSICAL THERAPIST-ADMINISTERED PHYSICAL ACTIVITY INTERVENTION FOR ADULTS AFTER KNEE REPLACEMENT: RESULTS OF A RANDOMIZED CONTROLLED TRIAL. Osteoarthritis Cartilage. 2022;30(Supplement 1):S54–5.

132. Rosal M, Ayers D, Li W, Oatis C, Borg A, Zheng H, et al. A randomized clinical trial of a peri-operative behavioral intervention to improve physical activity adherence and functional outcomes following total knee replacement. 2011;12:226.

133. Larsen J.B., Skou S.T., Laursen M., Bruun N.H., Arendt-Nielsen L., Madeleine P. A RANDOMIZED TRIAL OF NEUROMUSCULAR EXERCISE AND PAIN NEUROSCIENCE EDUCATION IN PATIENTS WITH CHRONIC PAIN AFTER TOTAL KNEE ARTHROPLASTY. Osteoarthritis Cartilage. 2024;32(Supplement 1):S518–9.

134. Skou ST, Roos EM, Laursen MB. A Randomized, Controlled Trial of Total Knee Replacement. N Engl J Med. 2016;374(7):692.

135. Teuscher DD, Lieberman JR. A Randomized, Controlled Trial of Total Knee Replacement. N Engl J Med. 2016;374(7):691–2.

136. Browne J.A. After Unilateral Total Knee Arthroplasty, Unsupervised Home Exercise Programs Were Noninferior to Outpatient Physiotherapy Services for Increasing Passive Flexion. J Bone Jt Surg Am Vol. 2019;101(22):2063.

137. Matsuse H, Nago T, Shinozaki N, Hashida R, Takano Y, Shiba N. Combined application of electrical stimulation and volitional contraction prevents muscle weakness. 2017;98(10):e45-.

138. Barker K., Room J., Knight R., Dutton S., Toye F., Leal J., et al. Community-based rehabilitation after knee arthroplasty: A randomised controlled trial with economic evaluations (CORKA trial): ISRCTN: 13517704. Physiotherapy. 2021;113(Supplement 1):e1–2.

139. Thiengwittayaporn S, Kangkano N, Krishnamra N, Charoenphandhu N. Comparison between diary-actuated rehabilitation program and conventional physical therapy on mobility and function following total knee arthroplasty. 2017;28:S194-S195.

140. Yuksel E, Unver B, Karatosun V. Comparison between kinesiotaping and cold therapy on muscle strength functional performance outcomes after total knee arthroplasty: preliminary results of a randomized controlled trial. 2016;75:1307-.

141. Guney Deniz H, Kinikli G, Onal S, Sevinc C, Caglar O, Yuksel I. Comparison of kinesio tape application and manual lymphatic drainage on lower extremity oedema and functions after total knee arthroplasty. 2018;77:1791-.

142. Hadley C., McGrath M., Prodoehl J.P., Cohen S.B., Emper W.D., Hammoud S., et al. Comparison of traditional physical therapy to internet-based physical therapy after knee arthroscopy: A prospective randomized controlled trial comparing patient outcomes and satisfaction. Orthop J Sports Med. 2019;7(7 Supplement 5).

143. Stevens-Lapsley J.E., Balter J.E., Kohrt W.M., Eckhoff D.G. Early neuromuscular electrical stimulation improves strength and functional performance after total knee arthroplasty. Arthritis and Rheumatism. 2009;60(SUPPL. 10):1940.

144. Eccentric exercise early after tka surgery: does negative-work work? Journal of Orthopaedic & Sports Physical Therapy. 2011;41(1):A20-1.

145. Alobaidi A, Mahmood A. Effect of continuous passive motion (CPM) exercises on post total knee arthroplasty (TKA) rehabilitation. 2018;Conference: 18th World Conference on Osteoporosis, Degenerative Disease and Musculoskeletal Disorders, WCO-IOF-ESCEO 2018. Poland. 29(1 Supplement 1):S162-S163.

146. Smith WA. Effect of incorporating the Fitbit fitness tracking technology into a prescribed exercise intervention program to improve long-term function in obese individuals one year following total knee arthroplasty. [Internet]. University of Mississippi; 2014. Available from: http://search.ebscohost.com/login.aspx?direct=true&db=rzh&AN=109776140&site=ehost-live

147. Kim JH, Noh JW, Kim MY, Lee JU, Yang SM, Kim J. Effect of orthopedic therapy on patients with total knee replacement for health science research. 2018;10(4):S41-.

148. Nishizaki K, Ikegami H, Tanaka Y, Saegusa H, Fukasawa T, Imai R, et al. Effect of supplementation with a combination of b-hydroxy b-methylbutyrate, L-arginine, and L-glutamine on quadriceps muscle strength in patients with osteoarthritis following total knee arthroplasty. 2014;33:S125-S126.

149. Matei D, Bighea A, Patru S, Traistaru R, Popescu R. Effectiveness of rehabilitation program on lower limb functional status after knee arthroplasty. 2014;25:S277-S278.

150. Juhl C.B., Roth S., Schierbeck R., Nielsen L.N., Nordlien A.-D., Hansen N.F., et al. Effectiveness of technology assisted exercise compared to usual care in total knee arthroplasty. Osteoarthritis and Cartilage. 2016;24(SUPPL. 1):S473.

151. Lee Y, Kim B. Effects of an early eccentrically based rehabilitation after total knee arth-roplasty. 2016;27(1 SUPPL. 1):S88.

152. Carozzo S., Coschignano F., Battaglia R., Giungato A., Grillo F., Pignolo L., et al. Effects of instrumental therapy compared to convection therapy in subjects undergoing knee replacement. Gait Posture. 2022;97(Supplement 2):22–3.

153. Muto T, Kanemura N, Takayanagi K, Ogawa R, Tanikawa H, Okuma K. Effects of multi-joint kinetics-chain exercise versus conventional exercise for patients with TKA: a randomized controlled trial. a 3-months research. 2015;101:eS1061-.

154. Lin SJ, Chen SH, Chai HM, Jan MH, Jiang CC. Effects of proprioceptive neuromuscular facilitation stretching technique on knee joint function in people following unilateral total knee arthroplasty. 2011;97:eS208-eS209.

155. Guney H, Vardar Yagli N, Caglar O, Yuksel I. Effects of relaxation techniques in total knee arthroplasty patients during their hospital stay. 2014;73. Available from: https://www.cochranelibrary.com/central/doi/10.1002/central/CN-01065730/full

156. Nishizaki K, Ikegami H, Tanaka Y, Imai R, Matsumura H. Effects of supplementation with a combination of β-hydroxy-β-methyl butyrate, L-arginine, and L-glutamine on postoperative recovery of quadriceps muscle strength after total knee arthroplasty. 2015;24(3):412-420.

157. van Eeden FM, van Halewijn KF, Swart NM, Festen DA, Bily W, Franz C, et al. Efficacy and Safety of Leg-Press Training With Moderate Vibration After Total Knee Arthroplasty Remains Unclear...Bily W, Franz C, Trimmel L, Loefler S, Cvecka J, Zampieri S, Kasche W, Sarabon N, Zenz P, Kern H. Effects of Leg-Press Training With Moderate Vibration on Muscle Strength, Pain, and Function After Total Knee Arthroplasty: A Randomized Controlled Trial. Arch Phys Med Rehabil. 2016 Jun;97(6):857-65. Archives of Physical Medicine & Rehabilitation. 2016;97(11):2018–9.

158. Ganz SB, Ranawat CS. Efficacy of formal knee flexion exercises on the achievement of functional milestones following total knee arthroplasty. Topics in Geriatric Rehabilitation. 2004;20(4):311–311.

159. Khaptagaev TB, Koneva ES, Strukov RN, Konev SM, Illarionov VE, Zhumanova EN, et al. [Efficiency of balance training with stabilizing platform in early postoperative rehabilitation of patients after arthroplasty]. Vopr Kurortol Fizioter Lech Fiz Kult. 2022;99(4. Vyp. 2):17–21.

160. Moffet H, Tousignant M, Nadeau S, Merette C, Boissy P, Corriveau H, et al. Evaluating the quality of an on-going clinical trial on the effectiveness of telerehabilitation service after knee arthroplasty: a one-year summary. 2011;97:eS821-eS822.

161. DeVivo K., Yang C.-H., Pellegrini C. Exercise Identity at the Start of Outpatient Physical Therapy Prospectively Predicts Physical Activity at 12-weeks after Total Knee Replacement. Arthritis Rheum. 2022;74(Supplement 9):1673–4.

162. Aslan S.N., Gevrek C., Demirel M., Atilla B., Kinikli G.I. HPR COMPARISON OF THE EFFECTIVENESS OF TELEREHABILITATION IN INDIVIDUALS WITH OSTEOARTHRITIS FOLLOWING TOTAL KNEE ARTHROPLASTY: RANDOMIZED CONTROLLED TRIAL. Ann Rheum Dis. 2024;83(Supplement 1):1194–5.

163. Weber-Spickschen S., Hardt S., Horstmann H., Krettek C. Improved early recovery after TKA through an app-based, feedback-controlled active muscle training-A prospective randomized trial. Orthopaedic Journal of Sports Medicine. 2018;6(4 Supplement 2).

164. Poolman R.W. In knee OA, adding total knee replacement to nonsurgical treatment improved pain and function at 12 mo. Annals of Internal Medicine. 2016;164(2):JC8.

165. Ko V, Naylor J, Harris I, Yeo A, Crosbie J. Is centre-based rehabilitation superior to home-based rehabilitation after knee replacement? A single-blind, randomised controlled trial. 2011;63(12):4043-.

166. Kadi M, Hepguler S, Dede E, Ozturk C, Aydogdu S, Aktuglu K, et al. Is electrotherapy effective in the management of pain, range of motion, quality of life, edema following total knee arthroplasty surgery? Randomized controlled trial. 2016;75:841-.

167. Partial body weight support and conventional physical therapy versus conventional physical therapy alone in the rehabilitation of patients following total knee arthroplasty: preliminary results. Journal of Orthopaedic & Sports Physical Therapy. 2011;41(1):A20–A20.

168. Negus J, Cawthorne D, Chen J, Scholes C, Parker D, March L. Patient outcomes using Wii-enhanced rehabilitation after total knee replacement - the TKR-POWER study. 2015;40:47-53.

169. Kierkegaard S, Jørgensen P, Søballe K, Mechlenburg I. Pelvic movements are restored to reference values during stair climbing but not during stepping one year after uni-compartmental knee arthroplasty-a secondary analysis of a randomized controlled trial. 2015;23:A111-A112.

170. Losina E., Yang H.Y., Stanley E.E., Katz J.N., Collins J.E. Physical activity as a novel outcome of total knee replacement: comparing self-report and objective PA assessments. Osteoarthritis and Cartilage. 2019;27(Supplement 1):S218–9.

171. Ryan T, Ohlson B, Adams R. Post operative total knee replacement rehabilitation: a new method using music video. 2009;91-B(SUPP_I):31-3d.

172. Christiansen MB, Thoma LM, Master H, Mathews D, Schmitt LA, White DK. Preliminary findings of a novel physical therapist administered physical activity intervention after total knee replacement. 2018;Conference: 2018 Osteoarthritis Research Society International, OARSI World Congress. United Kingdom. 26(Supplement 1):S334.

173. Macrinici G, Jiotis S, Dharmavaram S, Martini M, Almachnouk M, Chandler FJ, et al. Prospective, double-blind, randomized clinical trial to evaluate adductor canal nerve block versus femoral nerve block: early postoperative period functional outcomes after total knee arthroplasty. 2016;41(5). Available from: https://www.cochranelibrary.com/central/doi/10.1002/central/CN-01333671/full

174. Losina E, Collins J, Wright J, Daigle M, Donnell-Fink L, Strnad D, et al. Randomized controlled trial of postoperative care navigation in total knee arthroplasty patients: does one size fit all? 2014;66:S1249-S1250.

175. Piva S, Schneider M, Moore-Patterson C, Catelani M, Gil A, Klatt B, et al. Randomized trial on exercise at late-stage after total knee replacement. 2018;70:2185-2186.

176. Stevens-Lapsley JE, Balter JE, Wolfe P, Eckhoff DG, Schwartz RS, Schenkman M, et al. Relationship Between Intensity of Quadriceps Muscle Neuromuscular Electrical Stimulation and Strength Recovery After Total Knee Arthroplasty...[corrected] [published erratum appears in PHYS THER 2012; 92(10):1362]. Physical Therapy. 2012;92(9):1187–96.

177. Marcu I, Patru S, Matei D, Bighea A. Role of physical exercise in patients with knee arthroplasty for osteoarthritis. 2017;28:S330-.

178. Hamilton D, Beard D, Barker K, MacFarlane G, Murray G, Simpson H. Targeting physiotherapy to patients at risk of poor outcomes following total knee arthroplasty: the TRIO randomised controlled trial. 2019;105:e160-e161.

179. Hamilton D., Beard D., Barker K., MacFarlane G., Stoddart A., Murray G., et al. Targeting Rehabilitation to Improve Outcomes following total knee arthroplasty (TRIO): a randomised controlled trial of physiotherapy interventions. JBMR Plus. 2021;5(Supplement 2):13–4.

180. Andersen H. Technological assisted rehabilitation following total knee joint replacement. a randomised controlled non-inferiority trial. 2018;77:1823-1824.

181. Losina E, Collins J, Daigle M, Donnell-Fink L, Prokopetz J, Strnad D, et al. The AViKA (Adding Value in Knee Arthroplasty) postoperative care navigation trial: rationale and design features. 2013;14:290.

182. Goytizolo E, Ranawat A, Lin Y, Mayman D, Alexiades M, Soeters R, et al. The combination of adductor canal block and periarticular injection with an accelerated rehabilitation protocol: a novel technique for patients undergoing total knee replacement (ACB PAI). 2017;42(6). Available from: https://www.cochranelibrary.com/central/doi/10.1002/central/CN-01441750/full

183. Vural Gokay B, Karaca S, Unlusoy O, Gokay N. The comparison of analgesic effects of epidural analgesia, continuous femoral block and intravenous patient controlled analgesia. 2013;30:128.

184. Xiaolong Y, Jianzhong W. The effect of acupuncture during post-acute phase of rehabilitation after total knee arthroplasty. 2014;57:e193.

185. Karaman A., Yuksel I., Kinikli G.I., Atilla B. The effect of core stabilization training on functional performance, balance and quality of life in patients with total knee arthroplasty. Osteoarthritis and Cartilage. 2016;24(SUPPL. 1):S468.

186. Walnum C.F., Nielsen M.N., Husted K., Bagger J., Andersen J.R. The effect of maltodextrin after training in the early postoperative stage after total knee or hip arthroplasty a blinded, randomized, placebo controlled intervention study. Clinical Nutrition. 2015;34(SUPPL. 1):S35.

187. Alnahdi A, Zeni J, Snyder-Mackler L. The effect of progressive strengthening programs on function and gait mechanics after unilateral total knee arthroplasty: a randomized clinical trial. 2012;20:S104-S105.

188. Fell N. The effectiveness of mental practice as a complement to traditional therapy in rehabilitation outcomes of patients status-post total knee arthroplasty. 2001; Available from: https://www.cochranelibrary.com/central/doi/10.1002/central/CN-00676359/full

189. Eymir M., Unver B., Karatosun V. The effectiveness of relaxation exercises on pain, functional level and muscle strength in patients with total knee arthroplasty: A preliminary results. Annals of the Rheumatic Diseases. 2018;77(Supplement 2):1795.

190. Duong V., Dennis S., Harris A., Robbins S.R., Venkatesha V., Ferreira M., et al. The Effects Of A Disruptive Digital Technology Intervention Following Total Knee Replacement: Results From The Pathway Randomised Controlled Trial. Osteoarthritis Cartilage. 2023;31(Supplement 1):S30–1.

191. Pereira L, Jolles B. The effects of end-of-range grade a+ mobilisation following acute primary TKA. 2015;101:eS1193-eS1194.

192. Matsuse H. The effects of knee bending exercise combined with electrically stimulated contractions on serum levels of bone turnover markers following total knee arthroplasty: A randomized controlled trial. PM R. 2020;12(SUPPL 1):S22.

193. Darabseh M., Rawashdeh M., Darwish F. The effects of pedometer-based intervention on patients after total knee replacement surgeries. Physiotherapy. 2021;113(Supplement 1):e35–6.

194. Jayabalan P, Almeida GJ, Wan Huang, Sowa GA, Piva SR. THE INVESTIGATION OF CANDIDATE BIOMARKERS TO ASSESS THE EFFICACY OF A NOVEL REHABILITATION REGIMEN FOLLOWING TOTAL KNEE ARTHROPLASTY: A PILOT STUDY. American Journal of Physical Medicine & Rehabilitation. 2014;a72-3.

195. Fung V, Ho A, Shaffer J, Gomez M. The utilization of nintendo wii fit in the rehabilitation of outpatients following total knee replacements: preliminary results of a randomized controlled trial. 2010;91(10):e37-.

196. Fung V, Shaffer J, Chung E, Ho A, Gomez M. The utilization of nintendo wii fittm in the rehabilitation of outpatients following total knee replacements-a randomized controlled trial. 2011;97:eS419-.

197. Fusakul Y, Thiengwittayaporn S, Anukoolkarn K, Aranyavalai T, Phonpichit C, Saensri P, et al. Therapeutic effects of low-level laser therapy in postoperative total knee arthroplasty surgery: a double-blinded randomized, placebo-controlled trial. 2014;73. Available from: https://www.cochranelibrary.com/central/doi/10.1002/central/CN-01065813/full

198. Cawthorne D, March L, Parker D, Coolican M, Negus J. TKR-power-patient outcomes using wii enhanced rehabilitation after a total knee replacement. 2015;101:eS204-eS205.

199. Skou S, Roos E, Simonsen O, Laursen M, Rathleff M, Arendt-Nielsen L, et al. Total knee replacement followed by a non-surgical treatment program reduce localized and spreading pain sensitization-a pre-defined ancillary analysis from a randomized controlled trial. 2016;24:S182-S183.

200. Clatworthy M. Total knee replacement plus nonsurgical treatment was better than nonsurgical treatment alone for knee osteoarthritis. Journal of Bone and Joint Surgery - American Volume. 2016;98(10):873.

201. Skou S, Roos E, Laursen M, Rathleff M, Arendt-Nielsen L, Rasmussen S, et al. Two year outcome from two parallel randomized trials on total knee replacement and non-surgical treatment of knee osteoarthritis. 2017;25:S35-S36.

202. Chen L, Chen C, Lin S, Chien S, Su J, Huang C, et al. Aggressive continuous passive motion exercise does not improve knee range of motion after total knee arthroplasty. 2013;22(3-4):389-394.

203. Pötzelsberger B, Lindinger SJ, Stöggl T, Buchecker M, Müller E. Alpine Skiing With total knee Arthro Plasty ( ASWAP): effects on gait asymmetries. Scandinavian Journal of Medicine & Science in Sports. 2015;25:49–59.

204. Kristensen M, Pötzelsberger B, Scheiber P, Bergdahl A, Hansen CN, Andersen JL, et al. Alpine Skiing With total knee Arthro Plasty ( ASWAP): metabolism, inflammation, and skeletal muscle fiber characteristics. Scandinavian Journal of Medicine & Science in Sports. 2015;25:40–8.

205. Pötzelsberger B, Stöggl T, Scheiber P, Lindinger SJ, Seifert J, Fink C, et al. Alpine Skiing With total knee Arthro Plasty ( ASWAP): symmetric loading during skiing. Scandinavian Journal of Medicine & Science in Sports. 2015;25:60–6.

206. Hofstaedter T, Fink C, Dorn U, Pötzelsberger B, Hepperger C, Gordon K, et al. Alpine Skiing With total knee ArthroPlasty ( ASWAP): clinical and radiographic outcomes. Scandinavian Journal of Medicine & Science in Sports. 2015;25:10–5.

207. Kösters A, Pötzelsberger B, Dela F, Dorn U, Hofstaedter T, Fink C, et al. Alpine Skiing With total knee ArthroPlasty ( ASWAP): study design and intervention. Scandinavian Journal of Medicine & Science in Sports. 2015;25:3–9.

208. Sano Y, Iwata A, Wanaka H, Matsui M, Yamamoto S, Koyanagi J, et al. An easy and safe training method for trunk function improves mobility in total knee arthroplasty patients: a quasi-randomized controlled trial. 2018;13(10):e0204884.

209. Yi X, Lee JH, Yu X, Yi G, Lee HS. Assessing the Efficacy of the Early Rehabilitation Pathway in Combination with Morita Therapy after Hip and Knee Arthroplasty. J Healthc Eng. 2022;2022:4285197.

210. Kim T, Park K, Yoon S, Kim S, Chang C, Seong S. Clinical value of regular passive ROM exercise by a physical therapist after total knee arthroplasty. 2009;17(10):1152-1158.

211. Bedekar NPASASKsP. Comparative study of conventional therapy and additional yogasanas for knee rehabilitation after total knee arthroplasty. 2012;5(2):118-122.

212. Leach W, Reid J, Murphy F. Continuous passive motion following total knee replacement: a prospective randomized trial with follow-up to 1 year. 2006;14(10):922-926.

213. Fukaya T, Mutsuzaki H, Yoshikawa K, Koseki K, Iwai K. Effect of Training With the Hybrid Assistive Limb on Gait Cycle Kinematics After Total Knee Arthroplasty. Geriatr Orthop Surg Rehabil. 2021;12:21514593211049075.

214. Haas R, O'Brien L, Bowles KA, Haines T. Effectiveness of a weekend physiotherapy service on short-term outcomes following hip and knee joint replacement surgery: a quasi-experimental study. 2018;32(11):1493-1508.

215. Nakamura M, Kise C, Hasegawa S, Misaki S. Effectiveness of early high-intensity balance training for early home life independence after total knee arthroplasty: a pseudo-randomized controlled trial. Physical therapy research. 2020;23(1):79–86.

216. Hadamus A, Błażkiewicz M, Wydra KT, Kowalska AJ, Łukowicz M, Białoszewski D, et al. Effectiveness of Early Rehabilitation with Exergaming in Virtual Reality on Gait in Patients after Total Knee Replacement. J Clin Med. 2022;11(17).

217. Karandikar GS, Sabnis SM, Patwari SV, Bedekar NS, Rairikar SA, Shyam AK, et al. Effects of Closed Chain Exercises as Early Intervention on Knee Joint Proprioception after Total Knee Arthroplasty. Indian Journal of Physiotherapy & Occupational Therapy. 2014;8(1):28–31.

218. Kubota M, Kokubo Y, Miyazaki T, Matsuo H, Naruse H, Shouji K, et al. Effects of knee extension exercise starting within 4 h after total knee arthroplasty. Eur J Orthop Surg Traumatol. 2022;32(5):803–9.

219. Alonso-Rodríguez AM, Sánchez-Herrero H, Nunes-Hernández S, Criado-Fernández B, González-López S, Solís-Muñoz M. [Efficacy of hydrotherapy versus gym treatment in primary total knee prosthesis due to osteoarthritis: a randomized controlled trial]. An Sist Sanit Navar. 2021 Aug 20;44(2):225–41.

220. Maempel JF, Walmsley PJ. Enhanced recovery programmes can reduce length of stay after total knee replacement without sacrificing functional outcome at one year. Ann R Coll Surg Engl. 2015;97(8):563–7.

221. Yoshioka T, Kubota S, Sugaya H, Arai N, Hyodo K, Kanamori A, et al. Feasibility and efficacy of knee extension training using a single-joint hybrid assistive limb, versus conventional rehabilitation during the early postoperative period after total knee arthroplasty. J Rural Med. 2021;16(1):22–8.

222. Jaczewska-Bogacka J, Stolarczyk A. Improvement in Gait Pattern After Knee Arthroplasty Followed by Proprioceptive Neuromuscular Facilitation Physiotherapy. Adv Exp Med Biol. 2018;1096:1–9.

223. Matla J, Ogrodzka K, Bac A, Gądek A, Sorysz T. Ocena stanu funkcjonalnego pacjentów po zabiegu alloplastyki całkowitej stawu kolanowego. Advances in Rehabilitation. 2017;31(2):17–27.

224. Tayrose G, Newman D, Slover J, Jaffe F, Hunter T, Bosco J. Rapid mobilization decreases length-of-stay in joint replacement patients. Bull Hosp Jt Dis (2013). 2013;71(3):222–6.

225. Summers S.H., Gnecco T., Slotkin E.M., Law T.Y., Nunley R.M. Significant Cost Savings and Improved Early Clinical Outcomes in Medicare Patients Utilizing a Clinician-Controlled Telerehabilitation System Following Total Knee Arthroplasty. J Arthroplasty. 2024;39(8 Supplement 1):S137–42.

226. Wilk-Frańczuk M, Zemła J, Sliwiński Z. The application of biofeedback exercises in patients following arthroplasty of the knee with the use of total endoprosthesis. Med Sci Monit. 2010;16(9):CR423-426.

227. Pagnotta G, Rich E, Eckardt P, Lavin P, Burriesci R. The Effect of a Rapid Rehabilitation Program on Patients Undergoing Unilateral Total Knee Arthroplasty. Orthopaedic Nursing. 2017;36(2):112–23.

228. Yousefian Molla R, Sadeghi H, Kahlaee A. The Effect of Early Progressive Resistive Exercise Therapy on Balance Control of Patients with Total Knee Arthroplasty. 2017;33(4):286-294.

229. Candiri B, Talu B, Guner E, Ozen M. The effect of graded motor imagery training on pain, functional performance, motor imagery skills, and kinesiophobia after total knee arthroplasty: randomized controlled trial. Korean J Pain. 2023;36(3):369–81.

230. Thonga T, Stasi S, Papathanasiou G. The Effect of Intensive Close-Kinetic-Chain Exercises on Functionality and Balance Confidence After Total Knee Arthroplasty. Cureus. 2021;13(10):e18965.

231. Radulovic TN, Lazovic M, Jandric S, Bucma T, Cvjetkovic DD, Manojlovic S. The Effects of Continued Rehabilitation After Primary Knee Replacement. Med Arch. 2016;70(2):131–4.

232. Sanzo P, Niccoli S, Droll K, Puskas D, Cullinan C, Lees SJ. The effects of exercise and active assisted cycle ergometry in post-operative total knee arthroplasty patients - a randomized controlled trial. J Exp Orthop. 2021;8(1):41.

233. Müller M, Toussaint R, Kohlmann T. Total hip and knee arthroplasty : results of outpatient orthopedic rehabilitation. 2015;44(3):203-211.

234. Ference D, Ference RJ, Rempher E, Freeman DC. Total knee arthroplasty patients using the in-home X10 machine fully recovered. No additional therapy required. J Orthop. 2021;27:79–83.

235. Yoshikawa K, Mutsuzaki H, Sano A, Koseki K, Fukaya T, Mizukami M, et al. Training with Hybrid Assistive Limb for walking function after total knee arthroplasty. 2018;13(1):163-.

236. Hsu W.-H., Hsu W.-B., Shen W.-J., Lin Z.-R., Chang S.-H., Hsu R.W.-W. Twenty-four-week hospital-based progressive resistance training on functional recovery in female patients post total knee arthroplasty. Knee. 2019;26(3):729–36.

237. Beaupre L, Jones C. A randomized, controlled clinical trial comparing analgesic use, length of stay and health services utilization in patients receiving continuous passive motion, slider board therapy or physical therapy alone following total knee arthroplasty. 2001;83 Suppl 1:16.

238. Marmon AR, Snyder-Mackler L. ACTIVATION DEFICITS DO NOT LIMIT QUADRICEPS STRENGTH TRAINING GAINS IN PATIENTS AFTER TOTAL KNEE ARTHROPLASTY. International Journal of Sports Physical Therapy. 2014;9(3):329–37.

239. Kristensen M.S., Jorgensen P.B., Bogh S.B., Kierkegaard S., Mechlenburg I., Dalgas U. Acute and chronic effects of early progressive resistance training on knee pain and knee joint effusion after unicompartmental knee arthroplasty. Acta orthopaedica Belgica. 2018;84(3):262–8.

240. Peiris C, Taylor N, Shields N. Additional Saturday allied health services increase habitual physical activity among patients receiving inpatient rehabilitation for lower limb orthopedic conditions: a randomized controlled trial. 2012;93(8):1365-1370.

241. Würth S, Finkenzeller T, Pötzelsberger B, Müller E, Amesberger G. Alpine Skiing With total knee Arthro Plasty ( ASWAP): physical activity, knee function, pain, exertion, and well-being. Scandinavian Journal of Medicine & Science in Sports. 2015;25:74–81.

242. Kösters A, Rieder F, Wiesinger H, Dorn U, Hofstaedter T, Fink C, et al. Alpine Skiing With total knee ArthroPlasty (ASWAP): effect on tendon properties. 2015;25 Suppl 2:67-73.

243. Pötzelsberger B, Stöggl T, Lindinger SJ, Dirnberger J, Stadlmann M, Buchecker M, et al. Alpine Skiing With total knee ArthroPlasty (ASWAP): effects on strength and cardiorespiratory fitness. Scandinavian Journal of Medicine & Science in Sports. 2015;25:16–25.

244. Narici M, Conte M, Salvioli S, Franceschi C, Selby A, Dela F, et al. Alpine Skiing With total knee ArthroPlasty (ASWAP): impact on molecular and architectural features of musculo-skeletal ageing. Scandinavian Journal of Medicine & Science in Sports. 2015;25:33–9.

245. Boissy P, Tousignant M, Moffet H, Nadeau S, Brière S, Mérette C, et al. Conditions of Use, Reliability, and Quality of Audio/Video-Mediated Communications During In-Home Rehabilitation Teletreatment for Postknee Arthroplasty. 2016;22(8):637-649.

246. Larsen K, Hansen T, Thomsen P, Christiansen T, Søballe K. Cost-effectiveness of accelerated perioperative care and rehabilitation after total hip and knee arthroplasty. 2009;91(4):761-772.

247. Piva SR, Farrokhi S, Almeida G, Fitzgerald G K, Levison TJ, DiGioia AM. Dose-Associated Changes in Gait Parameters in Response to Exercise Programs after Total Knee Arthroplasty: Secondary Analysis of Two Randomized Studies. Int J Phys Med Rehabil. 2015;3(6):3–7.

248. Moutzouri M, Coutts F, Gliatis J, Billis E, Tsepis E, Gleeson N. Early initiation of home-based sensori-motor training improves muscle strength, activation and size in patients after knee replacement: a secondary analysis of a controlled clinical trial. BMC Musculoskeletal Disorders. 2019;20(1):1–10.

249. Kauppila A, Sintonen H, Aronen P, Ohtonen P, Kyllönen E, Arokoski J. Economic evaluation of multidisciplinary rehabilitation after primary total knee arthroplasty based on a randomized controlled trial. 2011;63(3):335-341.

250. Almeida G, Moore-Patterson C, Smith C, Jayabalan P, Piva S. Effect of changes in physical activity on cartilage degradation in knee osteoarthritis. 2018;70:444-445.

251. Hsieh CJ, DeJong G, Vita M, Zeymo A, Desale S. Effect of Outpatient Rehabilitation on Functional Mobility After Single Total Knee Arthroplasty: A Randomized Clinical Trial. JAMA Netw Open. 2020;3(9):e2016571.

252. Liao CD, Tsauo JY, Chiu YS, Ku JW, Huang SW, Liou TH. Effects of Elastic Resistance Exercise After Total Knee Replacement on Muscle Mass and Physical Function in Elderly Women With Osteoarthritis: A Randomized Controlled Trial. Am J Phys Med Rehabil. 2020;99(5):381–9.

253. Jiao S, Feng Z, Huang J, Dai T, Liu R, Meng Q. Enhanced recovery after surgery combined with quantitative rehabilitation training in early rehabilitation after total knee replacement: a randomized controlled trial. Eur J Phys Rehabil Med. 2024;60(1):74–83.

254. Liao C, Lin L, Huang Y, Huang S, Chou L, Liou T. Functional outcomes of outpatient balance training following total knee replacement in patients with knee osteoarthritis: a randomized controlled trial. 2015;29(9):855-867.

255. Naylor J, Ko V. Heart rate response and factors affecting exercise performance during home- or class-based rehabilitation for knee replacement recipients: lessons for clinical practice. 2012;18(2):449-458.

256. Pesce V, Notarnicola A, Setti S, Moretti L, Vicenti G, Moretti B. I-ONE therapy in patients undergoing total knee arthroplasty: results at 12 months. 2012;13:S44-.

257. Knoop J, Steultjens M, Roorda L, Lems W, van der Esch M, Thorstensson C, et al. Improvement in upper leg muscle strength underlies beneficial effects of exercise therapy in knee osteoarthritis: secondary analysis from a randomised controlled trial. 2015;101(2):171-177.

258. Langkilde A, Jakobsen T, Bandholm T, Eugen-Olsen J, Blauenfeldt T, Petersen J, et al. Inflammation and post-operative recovery in patients undergoing total knee arthroplasty-secondary analysis of a randomized controlled trial. 2017;25(8):1265-1273.

259. Naylor J, Crosbie J, Ko V. Is there a role for rehabilitation streaming following total knee arthroplasty? Preliminary insights from a randomized controlled trial. 2015;47(3):235-241.

260. Cheng YY, Chen CH, Wang SP. Isokinetic training of lower extremity during the early stage promote functional restoration in elder patients with disability after Total knee replacement (TKR) - a randomized control trial. BMC Geriatr. 2024;24(1):173.

261. Valtonen A, Pöyhönen T, Sipilä S, Heinonen A. Maintenance of aquatic training-induced benefits in mobility and lower extremity muscles among persons with unilateral knee replacement. 2011;97:eS1275-eS1276.

262. Arendt-Nielsen L, Simonsen O, Laursen M, Roos E, Rathleff M, Rasmussen S, et al. Pain and sensitization after total knee replacement or nonsurgical treatment in patients with knee osteoarthritis: identifying potential predictors of outcome at 12 months. 2018;22(6):1088-1102.

263. Moffet H, Tousignant M, Nadeau S, Mérette C, Boissy P, Corriveau H, et al. Patient Satisfaction with In-Home Telerehabilitation After Total Knee Arthroplasty: results from a Randomized Controlled Trial. 2017;23(2):80-87.

264. Tousignant M, Moffet H, Boissy P, Corriveau H, Cabana F, Marquis E. Patients and physiotherapists satisfaction of in-home telerehabilittion for post-knee arthroplasty. 2011;97:eS1246-eS1247.

265. Bade M., Struessel T., Paxton R., Winters J., Baym C., Stevens-Lapsley J. Performance on a Clinical Quadriceps Activation Battery Is Related to a Laboratory Measure of Activation and Recovery After Total Knee Arthroplasty. Archives of Physical Medicine and Rehabilitation. 2018;99(1):99–106.

266. Bade M, Kittelson J, Kohrt W, Stevens-Lapsley J. Predicting functional performance and range of motion outcomes after total knee arthroplasty. 2014;93(7):579-585.

267. Crosbie J, Naylor J, Harmer A, Russell T. Predictors of functional ambulation and patient perception following total knee replacement and short-term rehabilitation. 2010;32(13):1088-1098.

268. Liebs T, Herzberg W, Rüther W, Russlies M, Hassenpflug J. Quality-Adjusted Life Years Gained by Hip and Knee Replacement Surgery and Its Aftercare. 2016;97(5):691-700.

269. Stevens-Lapsley J, Balter J, Wolfe P, Eckhoff D, Schwartz R, Schenkman M, et al. Relationship between intensity of quadriceps muscle neuromuscular electrical stimulation and strength recovery after total knee arthroplasty. 2012;92(9):1187-1196.

270. Fleeton G, Harmer A, Nairn L, Crosbie J, March L, Crawford R, et al. Self-Reported Knee Instability Before and After Total Knee Replacement Surgery. 2016;68(4):463-471.

271. Skou ST, Roos EM, Laursen MB, Rathleff MS, Arendt-Nielsen L, Rasmussen S, et al. Total knee replacement and non-surgical treatment of knee osteoarthritis: 2-year outcome from two parallel randomized controlled trials. Osteoarthritis & Cartilage. 2018;26(5):N.PAG-N.PAG.

272. Lysack C, Dama M, Neufeld S, Andreassi E. A compliance and satisfaction with home exercise: a comparison of computer-assisted video instruction and routine rehabilitation practice. 2005;34(2):76-82.

273. Campbell K, Louie P, Bohl D, Edmiston T, Mikhail C, Li J, et al. A Novel, Automated Text-Messaging System Is Effective in Patients Undergoing Total Joint Arthroplasty. 2019;101(2):145-151.

274. Bellelli G, Buccino G, Bernardini B, Padovani A, Trabucchi M. Action Observation Treatment Improves Recovery of Postsurgical Orthopedic Patients: Evidence for a Top-Down Effect? Archives of Physical Medicine & Rehabilitation. 2010;91(10):1489–94.

275. Harper C, Dong Y, Thornhill T, Wright J, Ready J, Brick G, et al. Can therapy dogs improve pain and satisfaction after total joint arthroplasty? A randomized controlled trial. 2015;473(1):372-379.

276. Ma Y, Fan Z, Gao W, Yu Z, Ren M, Ma Q, et al. Cognitive therapeutic exercise in early proprioception recovery after knee osteoarthritis surgery. Front Rehabil Sci. 2022;3:915010.

277. Weaver F, Hughes S, Almagor O, Wixson R, Manheim L, Fulton B, et al. Comparison of two home care protocols for total joint replacement. 2003;51(4):523-528.

278. Mockford B, Thompson N, Humphreys P, Beverland D. Does a standard outpatient physiotherapy regime improve the range of knee motion after primary total knee arthroplasty? 2008;23(8):1110-1114.

279. Dong F, Li M, Liu J. Effect of department of orthopedics rehabilitation integrated mode on knee joint pain, function and quality of life in total knee arthroplasty. Biomedical research (india). 2017;28(21):9534-9537.

280. Mehta SJ, Hume E, Troxel AB, Reitz C, Norton L, Lacko H, et al. Effect of Remote Monitoring on Discharge to Home, Return to Activity, and Rehospitalization After Hip and Knee Arthroplasty: A Randomized Clinical Trial. JAMA Netw Open. 2020;3(12):e2028328.

281. Jogi P, Overend T, Spaulding S, Zecevic A, Kramer J. Effectiveness of balance exercises in the acute post-operative phase following total hip and knee arthroplasty: a randomized clinical trial. 2015;3. Available from: https://www.cochranelibrary.com/central/doi/10.1002/central/CN-01069766/full

282. Piva SR, Schneider MJ, Moore CG, Catelani MB, Gil AB, Klatt BA, et al. Effectiveness of Later-Stage Exercise Programs vs Usual Medical Care on Physical Function and Activity After Total Knee Replacement: A Randomized Clinical Trial. JAMA Network Open. 2019;2(2):e190018–e190018.

283. Li DY, Zhang WW, Tang XW. Effects of modified versus conventional training methods for high-flexion knee replacement. 2010;14(30):5543-5546.

284. Pohl T, Brauner T, Wearing S, Stamer K, Horstmann T. Effects of sensorimotor training volume on recovery of sensorimotor function in patients following lower limb arthroplasty. 2015;16(1). Available from: https://www.cochranelibrary.com/central/doi/10.1002/central/CN-01090172/full

285. Jogi P, Zecevic A, Overend T, Spaulding S, Kramer J. Force-plate analyses of balance following a balance exercise program during acute post-operative phase in individuals with total hip and knee arthroplasty: a randomized clinical trial. 2016;4. Available from: https://www.cochranelibrary.com/central/doi/10.1002/central/CN-01335559/full

286. Hoorntje A, Waterval-Witjes S, Koenraadt KLM, Kuijer PPFM, Blankevoort L, Kerkhoffs GMMJ, et al. Goal Attainment Scaling Rehabilitation Improves Satisfaction with Work Activities for Younger Working Patients After Knee Arthroplasty: Results from the Randomized Controlled ACTION Trial. J Bone Joint Surg Am. 2020;102(16):1445–53.

287. Aprile I, Rizzo R, Romanini E, De Santis F, Marsan S, Rinaldi G, et al. Group rehabilitation versus individual rehabilitation following knee and hip replacement: a pilot study with randomized, single-blind, cross-over design. 2011;47(4):551-559.

288. Barker KL, Room J, Knight R, Dutton S, Toye F, Leal J, et al. Home-based rehabilitation programme compared with traditional physiotherapy for patients at risk of poor outcome after knee arthroplasty: the CORKA randomised controlled trial. BMJ Open. 2021;11(8):e052598.

289. Mazurek J, Cieślik B, Wrzeciono A, Gajda R, Szczepańska-Gieracha J. Immersive Virtual Reality Therapy Is Supportive for Orthopedic Rehabilitation among the Elderly: A Randomized Controlled Trial. J Clin Med. 2023;12(24).

290. Brandes M, Wirsik N, Niehoff H, Heimsoth J, Möhring B. Impact of a tailored activity counselling intervention during inpatient rehabilitation after knee and hip arthroplasty - an explorative RCT. 2018;19(1):209.

291. Bethge M, Bartel S, Streibelt M, Lassahn C, Thren K. Improved outcome quality following total knee and hip arthroplasty in an integrated care setting: results of a controlled study. 2011;50(2):86-93.

292. Cui X.-Q., Wang H.-Z., Li W. Influence of time for continuous passive activities on range of motion following total knee arthroplasty. Journal of Clinical Rehabilitative Tissue Engineering Research. 2009;13(22):4237–40.

293. Iwakiri K, Ohta Y, Shibata Y, Minoda Y, Kobayashi A, Nakamura H. Initiating range of motion exercises within 24 hours following total knee arthroplasty affects the reduction of postoperative pain: A randomized controlled trial. Asia Pac J Sports Med Arthrosc Rehabil Technol. 2020;21:11–6.

294. Mahomed N, Davis A, Hawker G, Badley E, Davey J, Syed K, et al. Inpatient compared with home-based rehabilitation following primary unilateral total hip or knee replacement: a randomized controlled trial. 2008;90(8):1673-1680.

295. Barker KL, Room J, Knight R, Dutton SJ, Toye F, Leal J, et al. Outpatient physiotherapy versus home-based rehabilitation for patients at risk of poor outcomes after knee arthroplasty: CORKA RCT. Health Technol Assess. 2020;24(65):1–116.

296. Smith TO, Parsons S, Ooms A, Dutton S, Fordham B, Garrett A, et al. Randomised controlled trial of a behaviour change physiotherapy intervention to increase physical activity following hip and knee replacement: the PEP-TALK trial. BMJ Open. 2022;12(5):e061373.

297. Ficklscherer A, Stapf J, Meissner K, Niethammer T, Lahner M, Wagenhauser M, et al. Testing the feasibility and safety of the Nintendo Wii gaming console in orthopedic rehabilitation: a pilot randomized controlled study. 2016;12(6):1273-1278.

298. Wang Q, Hunter S, Lee RLT, Chan SWC. The effectiveness of a mobile application-based programme for rehabilitation after total hip or knee arthroplasty: A randomised controlled trial. Int J Nurs Stud. 2023;140:104455.

299. Hensman-Crook A. The effectiveness of physiotherapy intervention with home exercise programme versus patient directed home exercise programme following total knee replacement. 2011;41(S1):39.

300. Eichler S, Salzwedel A, Rabe S, Mueller S, Mayer F, Wochatz M, et al. The Effectiveness of Telerehabilitation as a Supplement to Rehabilitation in Patients After Total Knee or Hip Replacement: Randomized Controlled Trial. JMIR Rehabil Assist Technol. 2019;6(2):e14236.

301. Piva S, Gil A, Almeida G, DiGioia A, Levison T, Fitzgerald G. A balance exercise program appears to improve function for patients with total knee arthroplasty: a randomized clinical trial. 2010;90(6):880-894.

302. Paxton R, Forster J, Miller M, Gerron K, Stevens-Lapsley J, Christiansen C. A Feasibility Study for Improved Physical Activity After Total Knee Arthroplasty. 2018;26(1):7-13.

303. Li J, Wu T, Xu Z, Gu X. A pilot study of post-total knee replacement gait rehabilitation using lower limbs robot-assisted training system. 2014;24(2):203-208.

304. Frost H, Lamb S, Robertson S. A randomized controlled trial of exercise to improve mobility and function after elective knee arthroplasty. Feasibility, results and methodological difficulties. 2002;16(2):200-209.

305. McAvoy R. Aquatic and land based therapy vs. land therapy on the outcome of total knee arthroplasty: a pilot randomized clinical trial. Journal of Aquatic Physical Therapy. 2009;17(1):8–15.

306. Pellegrini C, Chang R, Dunlop D, Conroy D, Lee J, Van Horn L, et al. Comparison of a Patient-Centered Weight Loss Program starting before versus after knee replacement: a pilot study. 2018;12(5):472-478.

307. Artz N, Dixon S, Wylde V, Marques E, Beswick A, Lenguerrand E, et al. Comparison of group-based outpatient physiotherapy with usual care after total knee replacement: a feasibility study for a randomized controlled trial. 2017;31(4):487-499.

308. Minns Lowe C, Barker K, Holder R, Sackley C. Comparison of postdischarge physiotherapy versus usual care following primary total knee arthroplasty for osteoarthritis: an exploratory pilot randomized clinical trial. 2012;26(7):629-641.

309. Sindhu B, Sharma M, Biraynia RK. Comparison of Supervised Rehabilitation vs. Home Based Unsupervised Rehabilitation Programs after Total Knee Arthroplasty: A Pilot Study. Indian Journal of Physiotherapy & Occupational Therapy. 2013;7(3):50–3.

310. Piva S.R., Catelani M.B., Almeida G.J. Comprehensive behavioral intervention compared to standard of care exercise program after total knee arthroplasty: A pilot randomized trial. Arthritis and Rheumatism. 2013;65(SUPPL. 10):S356.

311. Piva S, Almeida G, Gil A, DiGioia A, Helsel D, Sowa G. Effect of Comprehensive Behavioral and Exercise Intervention on Physical Function and Activity Participation After Total Knee Replacement: a Pilot Randomized Study. 2017;69(12):1855-1862.

312. Stocker B, Babendererde C, Rohner-Spengler M, Müller UW, Meichtry A, Luomajoki H. Effective therapy to reduce edema after total knee arthroplasty Multi-layer compression therapy or standard therapy with cool pack - a randomized controlled pilot trial. Pflege. 2018;31(1):19–29.

313. Wylde V, Artz N, Dixon S, Marques E, Lenguerrand E, Blom A, et al. Effectiveness and cost-effectiveness of a group-based outpatient physiotherapy intervention following knee replacement for osteoarthritis: feasibility study for a randomised controlled trial. 2014;22:S433-.

314. Piva SR, Almeida G, Gil AB, Teixeira PE, Fitzgerald GK. Effectiveness and feasibility of a balance training program post total knee arthroplasty: pilot randomized trial. Journal of Orthopaedic & Sports Physical Therapy. 2009;39(1):A29-30.

315. Walls RJ, McHugh G, O'Gorman DJ, Moyna NM, O'Byrne JM. Effects of preoperative neuromuscular electrical stimulation on quadriceps strength and functional recovery in total knee arthroplasty. A pilot study. BMC Musculoskelet Disord. 2010;11:119.

316. Goto K, Morishita ,Takashi, Kamada ,Satoshi, Saita ,Kazuya, Fukuda ,Hiroyuki, Shiota ,Etsuji, et al. Feasibility of rehabilitation using the single-joint hybrid assistive limb to facilitate early recovery following total knee arthroplasty: A pilot study. Assistive Technology. 2017 Oct 2;29(4):197–201.

317. Kotani N, Morishita T, Saita K, Kamada S, Maeyama A, Abe H, et al. Feasibility of supplemental robot-assisted knee flexion exercise following total knee arthroplasty. J Back Musculoskelet Rehabil. 2020;33(3):413–21.

318. Harikesavan K., Chakravarty R.D., Maiya A.G., Hegde S.P., Shivanna S.Y. Hip abductor strengthening improves physical function following total knee replacement: One-year follow-up of a randomized pilot study. Open Rheumatology Journal. 2017;11((Maiya) Department of Physiotherapy, School of Allied Health Sciences, Manipal University, Manipal, India):30–42.

319. Byrne Notte B, Fazzini C, Mooney RA. Reiki's effect on patients with total knee arthroplasty: A pilot study. Nursing. 2016;46(2):17–23.

320. Cai L, Liu Y, Wei Z, Liang H, Liu Y, Cui M. Robot-assisted rehabilitation training improves knee function and daily activity ability in older adults following total knee arthroplasty. Res Nurs Health. 2023;46(2):203–9.

321. SUN MI KIM, SANG-RIM KIM, YONG KI LEE, BO RYUN KIM, EUN YOUNG HAN. The effect of mechanical massage on early outcome after total knee arthroplasty: a pilot study. Journal of Physical Therapy Science. 2015;27(11):3413–6.

322. Bugbee W, Pulido P, Goldberg T, D'Lima D. Use of an Anti-Gravity Treadmill for Early Postoperative Rehabilitation After Total Knee Replacement: a Pilot Study to Determine Safety and Feasibility. 2016;45(4):E167-73.

323. McInnes J, Larson M, Daltroy L, Brown T, Fossel A, Eaton H, et al. A controlled evaluation of continuous passive motion in patients undergoing total knee arthroplasty. 1992;268(11):1423-1428.

324. Siong N, Jin Y. An alternative early knee flexion regimen of continuous passive motion for total knee arthroplasty. 1999;2(2):53-63.

325. Ng TS, Yeo SJ. An alternative early knee flexion regimen of continuous passive motion for total knee arthroplasty. Physiotherapy Singapore. 1999;2(2):53–63.

326. Dowsey M, Kilgour M, Santamaria N, Choong P. Clinical pathways in hip and knee arthroplasty: a prospective randomised controlled study. 1999;170(2):59-62.

327. May LA, Busse W, Zayac D, Whitridge MR. Comparison of continuous passive motion (CPM) machines and lower limb mobility boards (LLiMB) in the rehabilitation of patients with total knee arthroplasty. Canadian Journal of Rehabilitation. 1999;12(4):257–63.

328. Harms M, Engstrom B. Continuous passive motion as an adjunct to treatment in the physiotherapy management of the total knee arthroplasty patient. 1991;77(4):301-307.

329. Montgomery F, Eliasson M. Continuous passive motion compared to active physical therapy after knee arthroplasty: similar hospitalization times in a randomized study of 68 patients. 1996;67(1):7-9.

330. Ritter M, Gandolf V, Holston K. Continuous passive motion versus physical therapy in total knee arthroplasty. 1989;(244):239-243.

331. Munin M, Rudy T, Glynn N, Crossett L, Rubash H. Early inpatient rehabilitation after elective hip and knee arthroplasty. 1998;279(11):847-852.

332. Singelyn F, Deyaert M, Joris D, Pendeville E, Gouverneur J. Effects of intravenous patient-controlled analgesia with morphine, continuous epidural analgesia, and continuous three-in-one block on postoperative pain and knee rehabilitation after unilateral total knee arthroplasty. 1998;87(1):88-92.

333. Hecht P, Bachmann S, Booth R, Rothman R. Effects of thermal therapy on rehabilitation after total knee arthroplasty. A prospective randomized study. 1983;(178):198-201.

334. Haug J, Wood LT. Efficacy of neuromuscular stimulation of the quadriceps femoris during continuous passive motion following total knee arthroplasty. Arch Phys Med Rehabil. 1988;69(6):423–4.

335. Gotlin R, Hershkowitz S, Juris P, Gonzalez E, Scott W, Insall J. Electrical stimulation effect on extensor lag and length of hospital stay after total knee arthroplasty. 1994;75(9):957-959.

336. Worland R, Arredondo J, Angles F, Lopez-Jimenez F, Jessup D. Home continuous passive motion machine versus professional physical therapy following total knee replacement. 1998;13(7):784-787.

337. Nielsen P, Rechnagel K, Nielsen S. No effect of continuous passive motion after arthroplasty of the knee. 1988;59(5):580-581.

338. Shepperd S, Harwood D, Jenkinson C, Gray A, Vessey M, Morgan P. Randomised controlled trial comparing hospital at home care with inpatient hospital care. I: three month follow up of health outcomes. 1998;316(7147):1786-1791.

339. Shepperd S, Harwood D, Gray A, Vessey M, Morgan P. Randomised controlled trial comparing hospital at home care with inpatient hospital care. II: cost minimisation analysis. 1998;316(7147):1791-1796.

340. Munin M, Glynn N, Rudy T, Crossett L, Rubash H. Randomized controlled trial of early inpatient rehabilitation after elective hip and knee arthroplasty. 1996;77:928.

341. Kumar P, McPherson E, Dorr L, Wan Z, Baldwin K. Rehabilitation after total knee arthroplasty: a comparison of 2 rehabilitation techniques. 1996;(331):93-101.

342. Chiarello C, Gundersen L, O'Halloran T. The effect of continuous passive motion duration and increment on range of motion in total knee arthroplasty patients. 1997;25(2):119-127.

343. Tong D., Zhang J., Liang X.-Y. Application of sensory and motor training in AIDET communication mode in patients after knee arthroplasty. World J Clin Cases. 2024;12(25):5720–8.

344. Fillingham Y, Darrith B, Lonner J, Culvern C, Crizer M, Della Valle C. Formal Physical Therapy May Not Be Necessary After Unicompartmental Knee Arthroplasty: a Randomized Clinical Trial. 2018;33(7S):S93-S99.e3.

345. Jørgensen P, Bogh S, Kierkegaard S, Sørensen H, Odgaard A, Søballe K, et al. The efficacy of early initiated, supervised, progressive resistance training compared to unsupervised, home-based exercise after unicompartmental knee arthroplasty: a single-blinded randomized controlled trial. 2017;31(1):61-70.

346. Lin H, Xu A, Wu H, Xu H, Lu Y, Yang H. Effect of Proprioception and Balance Training Combined with Continuous Nursing on BBS Score and HSS Score of Patients Undergoing Total Knee Arthroplasty. Comput Math Methods Med. 2022;2022:7074525.
