## Appendix 4 for "Registration and reporting characteristics of trials investigating exercise therapy following total knee arthroplasty: A systematic review"

| | | | |
|---|---|---|---|
| + | Low risk | D1 | Randomisation process |
| | | D2 | Deviations from the intended interventions |
| ! | Some concerns | D3 | Missing outcome data |
| | | D4 | Measurement of the outcome |
| - | High risk | D5 | Selection of the reported result |

| **Study ID** | **Experimental** | **Comparator** | **Outcome** | **D1** | **D2** | **D3** | **D4** | **D5** | **Overall** |
|---|---|---|---|---|---|---|---|---|---|
| Akbaba 2014 | The exercise programme with intensive supervision of physical therapist (2 days/1 week) was applied in Group 1. | The exercise programme with less intensive supervision of physical therapist (1 day/2 weeks) was applied in Group 2. | WOMAC function | ! | - | - | + | ! | - |
| Alsayani 2024 | Clinic based (PRT) 8w supervised 2x week with physio cable machines | Home based (PRT) same freq, duration, elastic band exercise | VAS at rest from 3m to 5m | ! | ! | ! | ! | ! | ! |
| An 2023 | CCE - individual Combined Kinetic Chain Exercise Program 30min for 5 days weekly 4 weeks | OKCE - individual open kinetic chain exercise - matched duration | WOMAC physical function from pre-surg to 6w | - | ! | + | ! | - | - |
| Bade 2017 | HI intervention was a high intensity, progression-based, rehabilitation program | LI intervention was a time-based rehabilitation program, Key differences from the HI program were an initial focus on isometric and ROM exercise for the first 4 weeks, a slower transition to weight-bearing exercises, less progression in difficulty of weight-bearing exercises, no resistance beyond body weight or elastic bands, and restricted activity outside of activities of daily living for the first 4 weeks, gradually building to 30 minutes | stair climb test | + | + | + | + | + | + |
| Bäcker 2021 | standard postoperative intervention - both grp same basic exercise CPM and NMES. In addition, app-based knee trainer for 6weeks 3-5 times daily | same standard postoperative intervention - basic exercise CPM and NMES as other group | lower extremity circumference pre-op to 6.74d | ! | ! | - | - | - | - |

| Study ID | Experimental | Comparator | Outcome | D1 | D2 | D3 | D4 | D5 | Overall |
|---|---|---|---|---|---|---|---|---|---|
| Bily 2016 | leg press vibration strength exercise 2s/w for 6w 3-hour total exercise time | physical therapy, manual therapy, individualized exercises 2s/wk for 6 weeks 6 hours total time | isometric knee-extensor strength | + | - | ! | ! | - | - |
| Bini 2016 | Self-administered exercise via internet-based tablet (home exercise) | 1 on 1 physical therapy exercise (self-administered) | VAS | ! | - | ! | - | - | - |
| Bohl 2019 | initiate physical therapy on day 0 | initiate physical therapy on day 1 | Length of Stay | + | ! | + | + | ! | ! |
| Bradbury 2024 | RPT - Same preop exer - after surgery app-based remote physical therapy (RPT) sessions at home twice daily for 6 weeks. Followed by knee maintenance program available for 3 months after surgery. | OPT - Same preop exercise - treatment by a licensed physical therapist of their choice 3 times per week for 6 weeks after surgery. Same maintenance program after available for 6 weeks after intervention. | knee ROM baseline to 6w | + | ! | + | - | ! | - |
| Bruun-Olsen 2013 | group based walking skill exercise 12 sessions | 12 session usual care physical therapy primarily ROM and resistance-based exercise, performed sitting | 6mwt | + | + | + | + | - | - |
| Buhagiar 2017 | 10 day inpatient physical therapy period followed by home exercises | received home exercises only | 6mwt | + | + | + | + | + | + |
| Can 2023 | CKCE home based closed kinetic chain progressive strength exercise for 6 weeks | Control - home based standard progressive loaded(weights) exercise for 6 weeks | VAS10 baseline 1w to 7w | - | ! | + | ! | ! | - |
| Carozzo 2022 | VFB - virtual feedback balance training - 20 mins 2xdaily 5d/week 6weeks + 40 mins conventional therapy | traditional 1h 2x/day 5d/week 6weeks - cryo, iso contractions, moderate prog resistance training, active/passive mobil, gait and balance training | Barthel index baseline to followup | + | ! | + | + | ! | ! |
| Cetinkaya and Karakoyun 2022 | same homebased exercise as control, plus elastic band exercises 4 times/daily for 4 weeks, plus attention phone calls | self-administered home based knee flexion and extension exercises for 4 weeks | VAS10 from 2w to 6w | + | - | + | - | - | - |

| Study ID | Experimental | Comparator | Outcome | D1 | D2 | D3 | D4 | D5 | Overall |
|---|---|---|---|---|---|---|---|---|---|
| Chang 2023 | EG - 30 minute daily home exercise 16w - unclear hybrid teaching and daily 2 sets of 5-10 rep exercises lasting 7 minutes + at least 15 minutes - abstract states one intervention hybrid teaching has multiple exercise program description - description is unclear | CG - routine care home-based exercise 16w- twice daily 8-10 reps exercises after discharge - unclear what exercise intensity etc. | KOOS preoperatively to 16w (end of intervention) | + | - | - | ! | - | - |
| Cheng 2022 | isokinetic maximal - outpatient 3 weekly for 4 weeks plus other standard exercises | isotonic 50% torque - outpatient 3 weekly for 4 weeks plus other standard exercises | TUG baseline to 4w | - | - | - | + | - | - |
| Chow 2010 | grp 1 1 hour unspecified exercise plus active stretching 2week intervention | grp 2 1 hour unspecified exercise plus passive stretching 2 week intervention | active ROM after 1st session | + | ! | ! | ! | ! | ! |
| Christiansen 2015 | weight-bearing biofeedback training | dose-matched standard-of-care rehab | weight-bearing ratio during 5time sit to stand test | + | ! | + | + | + | ! |
| Codine 2004 | same as control with hamstring exercise added | knee mobilisation (manually and with continuous passive motion), isometric strengthening for all the muscle groups of the knee and leg, proprioceptive enhancement and walking exercises | knee extension ROM | - | - | ! | - | - | - |
| Crawford 2021 | app based homebased self-management with exercise programme 3 sessions per day 6d/week for 6weeks | usual care - standardised 3 times pr weeks for 4 weeks | readmission rate at 90d | + | - | ! | ! | ! | - |
| DeJong 2020 | 2) body weight adjustable anti-gravity treadmill 12w 20 mins out of 40 min sessions | 1) stationary recumbent bike 12w 20 mins out of 40 min sessions | AM-PAC mobility score from baseline to end of intervention (2-3m) | + | ! | - | - | - | - |
| den Hertog 2012 | Fast track rehabilitation, 2h daily 6 days planned, started day of surgery, followed by similar post-discharge treatment | standard care rehabilitation,1h daily unaware of time-to-discharge, followed by similar post-discharge treatment | American Knee Society Score | + | ! | + | + | ! | ! |

| **Study ID** | **Experimental** | **Comparator** | **Outcome** | **D1** | **D2** | **D3** | **D4** | **D5** | **Overall** |
|---|---|---|---|---|---|---|---|---|---|
| Do and Yim 2020 | Hip strength and range of motion home-based with elastic band. 3 sets of 20. 4 weeks with 3 weekly sessions followed by 8 weeks home exercise | The control group (Group C) was assigned an active range of motion exercises. For 12 weeks three times pr week | knee flexion ROM baseline (week 0) to end of intervention (week 12) | + | ! | ! | + | - | - |
| Doerfler 2015 | high velocity exercise 2/w 8 weeks 60-80min duration (1s contractile phase 3s eccentric) | slow velocity exercise 2/w 8 weeks 60-80min duration (3s contractile, 3s eccentric phase) | 6mwt | + | - | ! | + | - | - |
| Dujin Park 2012 | modified quadriceps setting in addition to physical rehabilitation | all the same as in intervention group with conventional position of the quadriceps exercise | quadriceps muscle strength | - | - | + | ! | ! | - |
| Eisermann 2004 | computer aided training unsupervised 30m 3-5d a week | supervised group exercise 30 mins 3-5d a week | acceptance of computer-aided training | ! | ! | ! | - | - | - |
| Evgeniadis 2008 | POP group performed a supervised eight-week home exercise program to strengthen lower extremities | control group followed the standard preoperative evaluation, TKA, an inpatient rehabilitation program and follow-up controls | SF36 mental health | ! | - | ! | ! | ! | - |
| Eymir 2021 | AHSE Active heel slide exercise 60 mins pr day 1d from surgery + standard PT until discharge at 5,65d + home exercise after | CPM 1d + standard PT from surgery to discharge at 5,65d + home exercises after | active knee flexion 1-2d pre-surgery to discharge 5,65d | + | ! | ! | - | - | - |
| Fleischman 2019 | web based home exercise PT 6w | outpatient PT, 4-8 weeks usual care | ROM at 1m | + | + | + | + | ! | ! |
| Fransen 2017 | post-acute (6w after) group based exercise, home-based exercise until then | usual acute care varying contents | WOMAC pain at 12m | + | + | + | ! | - | - |
| Fung 2012 | 15min Wii board exercise added to 60mins physical therapy | 15min exercise added to 60mins of physical therapy (same exercises) | Length of Stay | ! | ! | + | + | ! | ! |

| Study ID | Experimental | Comparator | Outcome | D1 | D2 | D3 | D4 | D5 | Overall |
|---|---|---|---|---|---|---|---|---|---|
| Hamilton 2020 | Intensive outpatient physiotherapy. Participants will visit hospital for physiotherapy sessions once a week for 6 weeks in addition to completing prescribed exercises twice a week at home on their own. | Home physiotherapy exercises. Patients will be asked to complete prescribed exercises at home on their own 3 times a week for 6 weeks. | OKS from 6w after surgery to 52week follow-up | + | + | + | + | ! | ! |
| Han 2015 | HEP home exercise program (primarily mobility and own bodyweight exercises) | usual care treatment (clinic-based outpatient physiotherapy) | WOMAC pain at 6w | + | + | + | ! | - | - |
| Hardt 2018 | standard physiotherapy with 5 mins exercise added with a strength device | standard physical therapy | AROM | + | + | + | - | - | - |
| Harmer 2009 | Waterbased exercise plus homebased exercise | land-based exercise plus home based | 6mwt at 8w | + | + | + | + | ! | ! |
| Heikkilä 2017 | home based progressive exercise, with adherence/follow-up phone calls | home based exercise with no follow-up | normal gait velocity at 12m | ! | ! | + | + | - | - |
| Hepperger 2016 | hiking group based exercise | ADL individual home exercise | SCT 3 months (ascent) | + | ! | + | - | ! | - |
| Husby 2018 | standard fast-track and maximal strength training | standard fast-track and standard-home exercise | 1RM operated leg, leg press at 10w | + | - | ! | ! | - | - |
| Jacksteit 2021 | continuous active motion bilateral alternating (CAMbi) - low-load resistance training of the operated leg three times daily for ~30min and 30 min once daily for bilateral leg for 9d + standard in-hospital pt. | continuous passive motion (CPM) unilateral operated leg (active control group; standard-of-care therapy) 30min 3x pr day for 9d + standard in-hospital pt - same physio as intervention group (30mins). | Active knee flexion 1d preoperatively to 9d after surgery | + | ! | ! | + | - | - |
| Jakobsen 2014 | PST progressive strength training 15 minutes | CON without progressive strength training 15 minutes | 6mwt at 8w | + | + | + | + | ! | ! |

| Study ID | Experimental | Comparator | Outcome | D1 | D2 | D3 | D4 | D5 | Overall |
|---|---|---|---|---|---|---|---|---|---|
| Janhunen 2023 | IG homebased progressive exergaming - for 16 weeks. Progressive increase in available games, duration, reps, sets and intensity. | CG - 11-12 home exercises with progressive frequency, reps and sets | OKS from before surgery to 4 months after | + | ! | ! | ! | ! | ! |
| Jiao 2023 | high-intensity progressive rehabilitation training group - preop + postop for 5 days postop | RRT group (routine rehabilitation training group) postop only. The main differences from intervention: (1) No progressive training and no standardized instruction; (2) The training frequency is low; and (3) postoperative period only. | HSS score from preop to 2w (closest to intervention completion) | + | ! | - | + | ! | - |
| Johnson 2010 | Whole body vibration during squat, lunge, calfraise exercises, progressive load increase | traditional progressive strength training | operated MVIC pre-post | - | - | ! | ! | - | - |
| Karaman 2017 | pilates 6 weeks with monitoring | unclear, mix of patellar mobilisation, stretching, isotonic strength exercise, resistance exercises home exercise after discharge monitored once pr. week | berg balance test | - | - | ! | - | ! | - |
| Kauppila 2010 | standard physiotherapy added with a 10day multidisciplinary intervention at 2-4 months after surgery | standard physiotherapy intervention, conventional care, same as other group | WOMAC function at 12months | ! | ! | + | ! | ! | ! |
| Kelly 2015 | High velocity exercise 2/w 6 weeks, progressive exercises same as other group | Low velocity exercise 2/w 6 weeks, progressive exercises same as other group | 6mwt at mean 59d | + | ! | + | + | ! | ! |
| Khanli 2021 | same as control (routine, TENS, infrared) plus hip strength exercises. Routine: TENS and infrared treatment, and knee strength and ROM exercises (for total of 10 sessions, 3 sessions week) | Routine exercises, TENS and infrared treatment plus knee extensor and flexor strength and ROM exercises (10 sessions, 3 sessions pr week) | VAS10 pain from baseline (2d post) to follow-up (10 sessions later, maybe 4th week?) | + | ! | - | - | - | - |

| **Study ID** | **Experimental** | **Comparator** | **Outcome** | **D1** | **D2** | **D3** | **D4** | **D5** | **Overall** |
|---|---|---|---|---|---|---|---|---|---|
| Ko 2013 | one on one therapy including manual/cryo therapy and specific exercises. 2w usual care at hospital and same home exercise program as other groups | group based therapy 50min circuit exercise. 2w usual care at hospital and same home exercise program as other groups | OKS at 10w | + | + | + | ! | - | - |
| Kramer 2003 | 5-15 minute phone calls to check compliance once a month. Both groups had 3 times daily home exercise for first 12 weeks, then once daily after. | Clinic based group 1hr 2 times weekly outpatient PT. Both groups had 3 times daily home exercise for first 12 weeks, then once daily after | knee society clinical rating scale at 12 weeks | ! | ! | + | + | ! | ! |
| Larsen 2024 | 12-week neuromuscular exercise program (2 times pr week, 24 sessions of 1 hour grp exercise) and 2 PNE sessions (education) | PNE only, 2 hours of group education | KOOS4 from baseline to 12m | + | + | + | + | + | + |
| Lee 2021 | inpatient (presumably) dynamic balance training program (PDBT) with physical therapy for 30 minutes per day, five times per week for six weeks | presumably inpatient. General physical therapy (same as other group without balance exercise) | WOMAC pain baseline to 6weeks | ! | - | + | ! | - | - |
| Lenguerrand 2019 | Usual care and 1 weekly group exercise session for 6 weeks. Individualized home exercise plan after | Usual care. Individualized home exercise plan after | LEFS score at 12 months | ! | + | + | ! | + | ! |
| Lenssen 2006 | twice daily PT - 40 minutes | once daily PT - 20 minutes | Passive flexion ROM at 4d | + | + | + | + | ! | ! |
| Levine 2013 | NMES 14d preoperatively and 60d after surgery with ROM exercises | ROM exercises and progressive strengthening exercise for 60d | quadriceps strength get up and go test pre op to 6m post op | + | - | ! | - | - | - |
| Li 2019 | 2w hospital exercise, then 12 weeks tai chi chuan 5 times weekly of 45m (TG in table 1 and 2) | 2w hospital exercise, then 12 weeks of traditional physical exercise (unloaded basic mobility exercises) 5 days pr week 45 mins. (CG in table 1 and 2) | WOMAC function | + | - | - | ! | ! | - |

| Study ID | Experimental | Comparator | Outcome | D1 | D2 | D3 | D4 | D5 | Overall |
|---|---|---|---|---|---|---|---|---|---|
| Laio 2013 | conventional functional training and additional balance training programme 90 mins unclear how often | conventional Functional exercises 60min duration unclear how often | Forward functional reach test FHR from pre to post | + | ! | - | + | - | - |
| Liao 2020 | EG underwent two 60-min progressive heavy strength (65-80% 1RM) training sessions each week, for 12 wks. | The conservative physical therapy had 2 sessions per week. At each 60-min exercise session, active and passive range of motion exercise, stretching exercise, and cycling/treadmill. | body composition (appendicular lean mass index) from 2w pre-surgery (t0) to 4m after surgery (t2) | + | ! | - | + | - | - |
| Liebs 2010 | standard postop. Physiotherapy (ROM, strength, ADL, balance) plus ergometer cycling 3 times weekly for 3 weeks | standard postop. Physiotherapy (ROM, strength, ADL, balance) | WOMAC function from baseline to 3 months | + | ! | ! | ! | ! | ! |
| Liebs 2012 | early initiation d6 of aquatic therapy 30m 3x/w until w5, both groups normal therapy | delayed initiation d14 of aquatic therapy, 30m 3x/w until w5, both grps normal therapy | WOMAC function from before surgery to 3m | + | + | ! | ! | - | - |
| Madsen 2013 | group based strength and endurance exercise 2x/w for 6w, education and group discussion with home exercise. Starting 4 to 8 w after surgery | home based exercise same as other grp, and 1-2 visits with local physiotherapist. Starting 8-12 weeks after surgery | OKS baseline to 6m | ! | - | - | ! | ! | - |
| Maeda 2024 | intervention group performed knee extension exercises with robot suit HAL single joint 50 times per day and normal physical therapy exercise. The intervention was performed 5 times per week, and a total of 10 times for 10d. 2w total | control group standard physical therapy exercise - knee extension exercises 50 times per day. The intervention was performed 5 times per week, and a total of 10 times. 2w total | VAS100 pre-surgery to d10 | + | ! | + | - | - | - |
| Mau-Moeller 2014 | all grps normal physical therapy during and 3w after discharge. ST - sling exercises 30m each day (elevated knee flexion with mechanical support) | all grps normal physical therapy during and 3w after discharge. CPM 30m each day | passive knee flexion range of motion at discharge (post test) | + | + | + | ! | ! | ! |

| Study ID | Experimental | Comparator | Outcome | D1 | D2 | D3 | D4 | D5 | Overall |
|---|---|---|---|---|---|---|---|---|---|
| Moffet 2004 | 12 supervised rehabilitation sessions with individualized home exercises performed on the days without supervised treatments. All subjects attended the 12 supervised rehabilitation sessions (duration, 60 –90min) over a period of 6 to 8 weeks started 2months after surgery. Both groups performed home exercise program started directly after TKA | usual care with home training exercises - unclear what usual care entails | 6mwt from baseline (2m) to 6m POST2 | + | ! | + | + | ! | ! |
| Moffet 2015 | Telerehabilitation group. Patient education, physical therapy assessment before and after and 30min exercise for 16 sessions, 2m. Same as other grp | face to face home visit. Patient education, physical therapy assessment before and after and 30min exercise for 16 sessions, 2m. Same as other grp | WOMAC from discharge to 4m, per protocol analysis | + | - | + | ! | ! | - |
| Molla 2017 | Extra early resistive exercises. The patients in both groups received 3 rehabilitation sessions a week for an hour per session and for 6 weeks (all patients were discharged from hospital within 3-4 days from the date of surgery). | The patients in both groups received 3 rehabilitation sessions a week for an hour per session and for 6 weeks (all patients were discharged from hospital within 3-4 days from the date of surgery). | berg balance test | - | ! | + | + | - | - |
| Monticone 2013 | patient education in fear-avoidance, home-exercise book about managing kinesiophobia plus 60m home exercise 2x week and monthly phone calls from rehab team to enhance adherence | general advice to stay active and recover usual ADL on basis of exercises learned during hospitalization | KOOS-ADL from t0 to t1 | + | - | + | ! | - | - |

| Study ID | Experimental | Comparator | Outcome | D1 | D2 | D3 | D4 | D5 | Overall |
|---|---|---|---|---|---|---|---|---|---|
| Moutzouri 2018 | Intervention group: Participants undergo the early initialised, self-managed, enhanced sensori-motor programme as well as the standard rehabilitation. This includes participants being prescribed to perform the same number of exercises daily for 35-45 minutes which includes walking (progressively for 10-20 minutes) for six weeks. This is followed by a further six weeks of training (three times weekly for 45 minutes). Participants received support during the programme including encouragement to continue with the self-managed exercises | Control group: Participants receive the usual level of care which consists of 15 home-based exercises, focusing on improving knee motion and muscle strengthening. | TUG from baseline (presurg) to 14w | + | - | + | - | ! | - |
| Núñez-Cortés 2024 | (1) intervention group (elastic resistance strengthening for 3 days) 4 exercises to failure RPE controlled | (2) control group (conventional inpatient physical therapy exercise) 7 exercises by reps no resistance for 3 days | strength by number of reps in each exercise - not reported | + | ! | + | - | ! | - |
| Papotto 2012 | high intensity stretching home therapy. Patients were asked to use the device multiple sessions per day. Each session was broken into 5- to 10-min increments of stretching followed by a 5- to 10-min recovery interval. Patients were asked to repeat the stretch-recovery cycles until 20–30 min had passed, and were asked to repeat treatment sessions until they had achieved 60 min of end-range stretching per day. | Low intensity stretch home therapy. Patients were asked to use the StaticPro Knee device in three 30-min increments per day and were asked to routinely increase the force applied to the joint every 5 min during each of the 30-min treatment sessions. | ROM (passive knee flexion) | ! | - | + | - | - | - |
| Park 2024 | CPM + BOSU for 2 weeks probably 3 times per week (6 total sessions) | CPM + stretching (Proprioceptive neuromuscular facilitation) for 2 weeks either twice pr day or 3 times per week (either 20 sessions or 6 sessions) | VAS10 pre to post | ! | - | - | - | - | - |
| Pastore 2015 | 2m intensive rehabilitation exercise program | 3m standard rehabilitation exercise program | Borg CR10 | + | - | - | - | - | - |

| **Study ID** | **Experimental** | **Comparator** | **Outcome** | **D1** | **D2** | **D3** | **D4** | **D5** | **Overall** |
|---|---|---|---|---|---|---|---|---|---|
| Piqueras 2013 | 1h interactive virtual software remote rehabilitation for 10 days | 1hr standard TKA rehab for 10 days | active knee flexion ROM | + | - | - | + | - | - |
| Rahmann 2009 | 10days daily aquatic physiotherapy and ward treatment | 10 days daily physiotherapy exercise with an additional ward treatment (2 total). Ward treatment contains bodyweight ADL exercises | hip abduction strength d14 | ! | - | - | + | - | - |
| Rajan 2004 | inpatient physiotherapy and 4-6 times outpatient physiotherapy. Both grps given home exercise regime to follow after discharge. | inpatient physiotherapy only. Both grps given home exercise regime to follow after discharge. | Range of motion at 3 months | ! | - | + | ! | ! | - |
| Russell 2003 | telerehabilitation 45 min session once pr week for 6 weeks | face-to-face 45 min session once pr week for 6 weeks | TUG | - | - | + | + | - | - |
| Russell 2011 | telerehabilitation 45 min session once pr week for 6 weeks. Addition to twice daily home exercise | face-to-face 45 min session once pr week for 6 weeks (control) in outpatient setting. Addition to twice daily home exercise | WOMAC baseline to 6w | + | ! | + | ! | ! | ! |
| Sattler 2019 | pedaling based exercise 20 twice daily until discharge, continued after, until 2w post-op review | multi exercise - 10 exercise program 20 mins twice daily until discharge, continued until two-week post op review. | 6mwt from baseline to end of intervention (t2) | + | ! | + | + | ! | ! |
| Schache 2019 | experimental - add hip exercises. 12 days of inpatient physiotherapy followed by 6 weeks of outpatient physiotherapy, which aimed to improve knee range of movement, strength and mobility. The experimental group completed a standard rehabilitation protocol with the addition of hip abductor strengthening. | control - standard of care. 12 days of inpatient physiotherapy followed by 6 weeks of outpatient physiotherapy, which aimed to improve knee range of movement, strength and mobility. The control group completed the same standard rehabilitation protocol, with the addition of 15 minutes of general functional exercises | KOOS from baseline to 6w follow-up | + | + | + | + | + | + |
| Schulz 2018 | controlled active motion (CAM) | continuous passive motion (CPM) | KOOS symptoms subscale from pre surgery to hospital discharge (end of intervention) | + | ! | + | ! | ! | ! |

| Study ID | Experimental | Comparator | Outcome | D1 | D2 | D3 | D4 | D5 | Overall |
|---|---|---|---|---|---|---|---|---|---|
| Shabbir 2017 | treatment group Functional activities included squats, lunges and steps (step up, step down). Functional activities were demonstrated to the treatment group by trained physical therapists. Patients were observed while performing those functional activities. They were provided with illustrated pamphlet of exercises as a home exercise plan. | control group home based muscle strengthening program and functional exercise once per day 5 days a week. For 4 weeks | improvement in knee extension lag baseline to 4 weeks | ! | ! | + | + | - | - |
| Suh 2017 | ECC-CON - eccentric and concentric exercise. Both groups performed concentric strength exercise using air resistance machines. ecc con did excentric recumbent cycle ergometer exercise for 30 minutes per session. All rehabilitation programs were performed five times per week for a 2-week period under the supervision of physical therapists. | CON - concentric only exercise. Both groups performed concentric exercise using air resistance machines. con group did concentric cycle ergometer for 30 minutes per session. All rehabilitation programs were performed five times per week for a 2-week period under the supervision of physical therapists. | change in quadriceps muscle strength (peak torque) between pre-surgery and at 4 weeks post-surgery | ! | ! | + | - | ! | - |
| Tanaka 2017 | HAL hybrid assisted limb exercise 20 mins and 20 mins conventional exercise. 5 weekly sessions for 2 weeks, 10 sessions total | Conventional group with 40 mins conventional gait rehab exercise. 5 weekly sessions for 2 weeks, 10 sessions total | 10m walking speed 1-2 weeks | + | - | ! | + | ! | - |
| Tanaka 2019 | IFR intensive functional rehabilitation, in addition to standard care for 2weeks | standard care until 2 weeks | Functional Independence Measure (motor subscale) | + | - | ! | ! | ! | - |
| Teissier 2020 | ECC/CON. Standard rehab and 3 sessions per week which were replaced by exclusively eccentric work. | CON group. Standard rehab with all its concentric exercise sessions consecutively, and subjects trained alternately for knee extensors and flexors. For each exercise, subjects did 3–5 sets with 5–10 repetitions at a weight equivalent to 60–80 % of their individual's estimated 1 RM and a 3–5 min rest was given between the sets | TUG pre to post intervention | - | ! | + | - | - | - |

| Study ID | Experimental | Comparator | Outcome | D1 | D2 | D3 | D4 | D5 | Overall |
|---|---|---|---|---|---|---|---|---|---|
| Tousignant 2011 | telerehabilitation two sessions per week for eight weeks (16 sessions) | usual care home visit/outpatient for about 2 months | ROM flexion knee 1w to 2m | + | - | ! | ! | ! | - |
| Trudelle-Jackson 2020 | High velocity elastic band exercise 8 weeks 3-4 times pr week home exercise, plus step monitoring goal | Step monitoring goal only (attention control - somewhat) | quadriceps muscle torque normalized to bodyweight pre to post | ! | - | - | ! | - | - |
| Tsukada 2020 | HTS. Electrical antagonist stimulation resistance training 19 mins 3x pr week added to 40 mins standard of care 5 times pr. week for 12 weeks | 40 mins standard of care - same as intervention group 5 times pr week for 12 weeks | Knee extension strength operated side from before surgery to 6 weeks | ! | - | + | ! | - | - |
| Unver 2016 | daily, 10 weighted home exercise program 8 weeks | daily, 10 unloaded home exercise (matched) for 8 weeks | isometric quadriceps strength after 8 weeks | + | ! | + | - | ! | - |
| Valtonen 2010 | aquatic exercise ca 50 mins twice weekly for 12 weeks | no intervention | habitual walking speed | + | ! | + | - | ! | - |
| Vourenmaa 2014 | guidance in home exercise (3 sessions/week) at 2m post-op, 3m post-op and 6m post-op with progression of home exercise program. | guidance in home exercise (3 sessions/week) at 2m post-op. Usual care | WOMAC pain | + | - | + | ! | ! | - |
| Warner 2020 | Experimental group was given the core stability exercises, along with routine physical therapy exercise. 6 days pr week for 6 weeks | Control group received routine exercises (same as intervention grp): ankle pumps, straight leg raise in supine, short arc quad, hip abduction in side lying, squats and walk. 6 days pr week for 6 weeks | LEFS baseline to 6w from surgery | ! | - | - | - | ! | - |
| Yang 2013 | 1 of 3 EXP I - rehabilitation exercise and traditional chinese manupulation, also CPM and oral NSAID | 3 of 3 EXP II - traditional chinese manupulation, also CPM and oral NSAID | VAS baseline d2 to discharge d4-5 | - | ! | ! | ! | ! | - |

| Study ID | Experimental | Comparator | Outcome | D1 | D2 | D3 | D4 | D5 | Overall |
|---|---|---|---|---|---|---|---|---|---|
| Zietek 2015 | intensive protocol - 15 min walk highweight rolling walker twice on postopday 1 3 hrs between walks, all other intervention same btw grps | standard protocol - 15 min walk highweight rolling walker once on postopday 1, all other intervention same btw grps | VAS while walking from baseline d0 to postop day 2 | + | ! | + | + | ! | ! |
| Xu 2021 | KFEH - homebased enhanced knee flexion programme with low stool - 20-40 mins, 8-15 reps, 5 days pr week first 8 weeks, 2-3 days pr week after for subsequent 10 months | SPT - standardised physical therapy - after discharge 24 sessions of a physiotherapy and rehabilitation program 2 days/week for the first 7 weeks, followed by 1 day/month for the subsequent 10 months of the year. | VAS10 from pre-surgery to 12m | ! | ! | + | - | - | - |
