## Supplementary figures and images for "Registration and reporting characteristics of trials investigating exercise therapy following total knee arthroplasty: A systematic review"

### Appendix 5

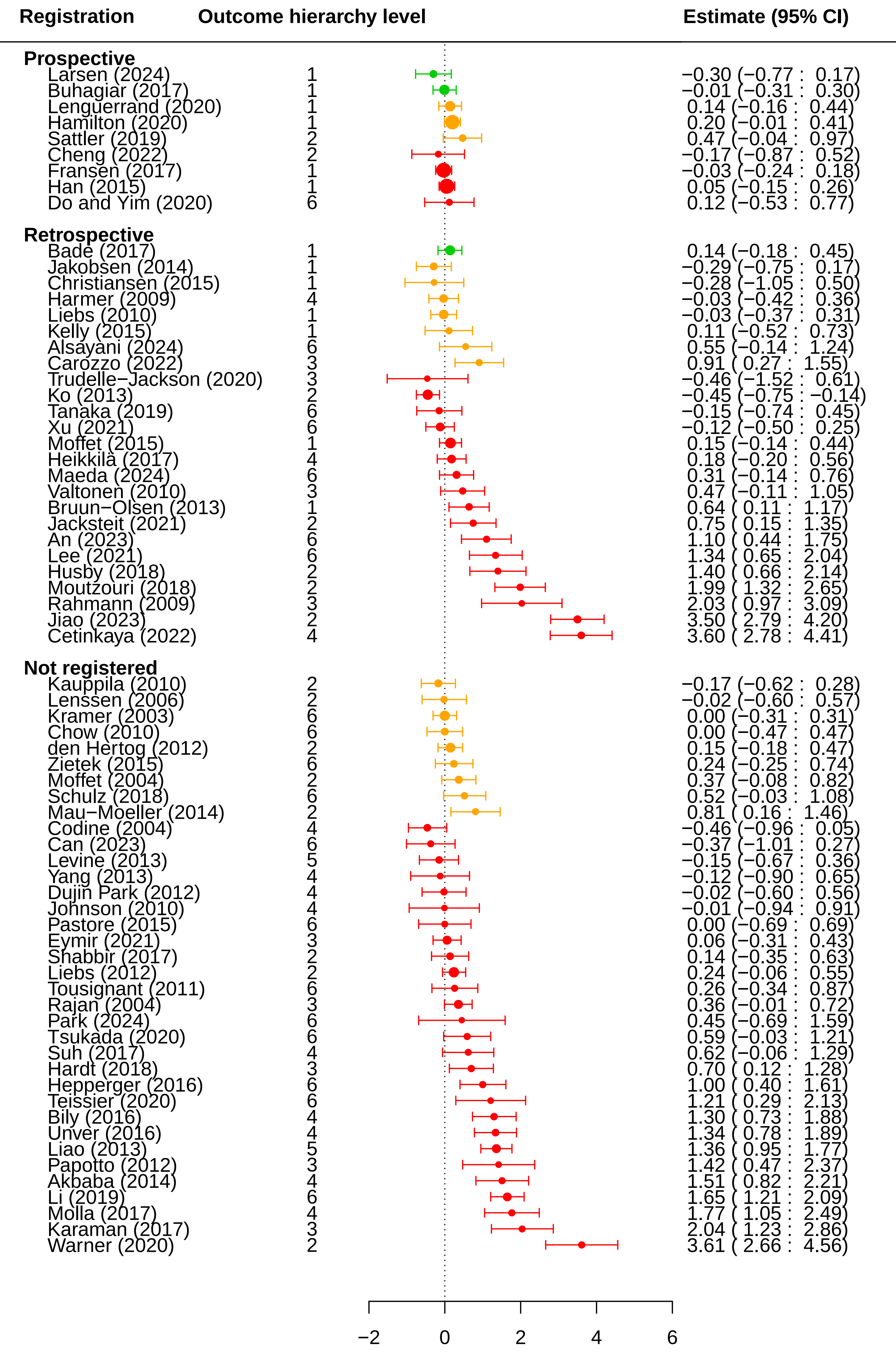

### Appendix 6

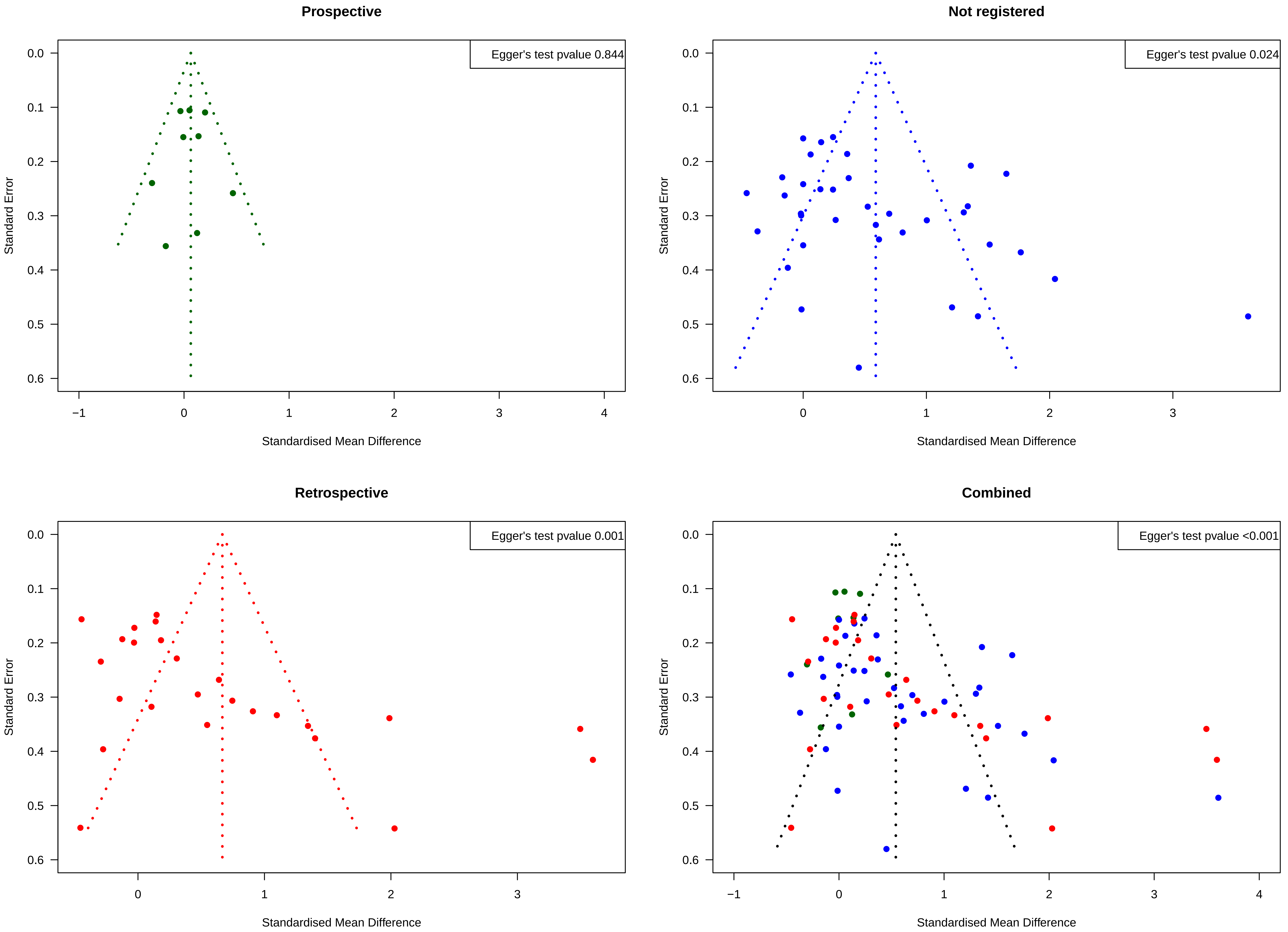
