## Appendix 7 for "Registration and reporting characteristics of trials investigating exercise therapy following total knee arthroplasty: A systematic review"

| Exposure | Comparison | Interaction | 95% CI | P-value | Bonferroni-corrected P-value | Tau² | Tau² impact (% reduction of between study variance) | Studies excluded (missing variables) |
|---|---|---|---|---|---|---|---|---|
| Participants | Pr. 10 extra participants in study | | | | | | | |
| | Retrosp. vs. Prosp. | 0.063 | [-0.01, 0.14] | 0.094 | 1 | 0.448 | 29.5% | (1) |
| | Nonreg. vs. Prosp. | 0.015 | [-0.06, 0.09] | 0.677 | 1 | | | |
| Multicentre study | Yes compared to No | | | | | | | |
| | Retrosp. vs. Prosp. | 0.597 | [-0.62, 1.82] | 0.333 | 1 | 0.452 | 28.9% | (1) |
| | Nonreg. vs. Prosp. | 0.183 | [-1.19, 1.56] | 0.791 | 1 | | | |
| Reported a single primary outcome (not hierarchy) | Yes compared to No | | | | | | | |
| | Retrosp. vs. Prosp. | 0.064 | [-1.16, 1.29] | 0.918 | 1 | 0.489 | 23.1% | (1) |
| | Nonreg. vs. Prosp. | 0.056 | [-1.17, 1.28] | 0.927 | 1 | | | |
| Study arms | 3 or more compared to 2 | | | | | | | |
| | Retrosp. vs. Prosp. | 0.412 | [-1.17, 1.99] | 0.603 | 1 | 0.485 | 23.7% | (1) |
| | Nonreg. vs. Prosp. | 0.299 | [-1.25, 1.85] | 0.701 | 1 | | | |
| Risk of Bias | Low compared to High | | | | | | | |
| | Retrosp. vs. Prosp. | 0.421 | [-1.53, 2.38] | 0.668 | 1 | 0.466 | 26.7% | (1) |
| | Nonreg. vs. Prosp. | N/A | N/A | N/A | N/A | | | |
| | Some concerns compared to High | | | | | | | |
| | Retrosp. vs. Prosp. | 0.628 | [-0.73, 1.99] | 0.359 | 1 | | | |
| | Nonreg. vs. Prosp. | 0.349 | [-0.95, 1.65] | 0.593 | 1 | | | |
| Primary outcome hierarchy level | 2 compared to 1 | | | | | | | |
| | Retrosp. vs. Prosp. | -1.178 | [-2.70, 0.34] | 0.126 | 1 | 0.475 | 25.3% | (1) |
| | Nonreg. vs. Prosp. | -0.089 | [-2.15, 1.97] | 0.931 | 1 | | | |
| | 3 compared to 1 | | | | | | | |
| | Retrosp. vs. Prosp. | N/A | N/A | N/A | N/A | | | |
| | Nonreg. vs. Prosp. | 0.118 | [-1.99, 2.22] | 0.911 | 1 | | | |
| | 4 and 5 compared to 1 | | | | | | | |
| | Retrosp. vs. Prosp. | N/A | N/A | N/A | N/A | | | |
| | Nonreg. vs. Prosp. | 0.613 | [-1.45, 2.68] | 0.554 | 1 | | | |
| | 6 compared to 1 | | | | | | | |
| | Retrosp. vs. Prosp. | -0.119 | [-1.99, 1.75] | 0.899 | 1 | | | |
| | Nonreg. vs. Prosp. | N/A | N/A | N/A | N/A | | | |
| Sample size calculation reported | Yes compared to No | | | | | | | |
| | Retrosp. vs. Prosp. | -0.528 | [-2.26, 1.21] | 0.546 | 1 | 0.480 | 24.5% | (1) |
| | Nonreg. vs. Prosp. | -0.067 | [-1.68, 1.55] | 0.934 | 1 | | | |

| Exposure | Comparison | Interaction | 95% CI | P-value | Bonferroni-corrected P-value | Tau² | Tau² impact (% reduction of between study variance) | Studies excluded (missing variables) |
|---|---|---|---|---|---|---|---|---|
| Dropouts description | Yes compared to No | | | | | | | |
| | Retrosp. vs. Prosp. | N/A | N/A | N/A | N/A | 0.405 | 36.3% | (1) |
| | Nonreg. vs. Prosp. | -1.630 | [-2.86, -0.40] | 0.010 | 0,157 | | | |
| | Partial compared to No | | | | | | | |
| | Retrosp. vs. Prosp. | N/A | N/A | N/A | N/A | | | |
| | Nonreg. vs. Prosp. | -2.200 | [-4.23, -0.18] | 0.033 | 0,497 | | | |
| Adverse events reporting | Yes compared to No | | | | | | | |
| | Retrosp. vs. Prosp. | 0.466 | [-0.75, 1.68] | 0.448 | 1 | 0.437 | 31.3% | (1) |
| | Nonreg. vs. Prosp. | -0.598 | [-1.76, 0.56] | 0.305 | 1 | | | |
| | Partial compared to No | | | | | | | |
| | Retrosp. vs. Prosp. | N/A | N/A | N/A | N/A | | | |
| | Nonreg. vs. Prosp. | -0.085 | [-1.84, 2.01] | 0.930 | 1 | | | |
| Following intent to treat analysis principles | Per protocol compared to ITT | | | | | | | |
| | Retrosp. vs. Prosp. | -0.361 | [-1.52, 0.80] | 0.534 | 1 | 0.363 | 42.9% | (1) |
| | Nonreg. vs. Prosp. | -0.381 | [-1.50, 0.74] | 0.498 | 1 | | | |
| | Unclear compared to ITT | | | | | | | |
| | Retrosp. vs. Prosp. | N/A | N/A | N/A | N/A | | | |
| | Nonreg. vs. Prosp. | 0.998 | [-0.15, 2.15] | 0.089 | 1 | | | |
| Time since surgery (intervention start) | Initiated 100 days later | | | | | | | |
| | Retrosp. vs. Prosp. | 0.139 | [-0.24, 0.52] | 0.469 | 1 | 0.485 | 23.7% | (1–5) |
| | Nonreg. vs. Prosp. | -0.073 | [-0.25, 0.10] | 0.413 | 1 | | | |
| Intervention duration (follow-up – initiated) | 100 days longer | | | | | | | |
| | Retrosp. vs. Prosp. | 0.272 | [-0.22, 0.76] | 0.268 | 1 | 0.479 | 24.6% | (1–5) |
| | Nonreg. vs. Prosp. | 0.038 | [-0.57, 0.64] | 0.900 | 1 | | | |
| Primary outcome domain | Disability compared to Composite | | | | | | | |
| | Retrosp. vs. Prosp. | 0.301 | [-1.89, 2.49] | 0.784 | 1 | 0.515 | 19.0% | (1) |
| | Nonreg. vs. Prosp. | -0.279 | [-2.45, 1.90] | 0.799 | 1 | | | |
| | Pain compared to Composite | | | | | | | |
| | Retrosp. vs. Prosp. | -0.200 | [-2.03, 1.63] | 0.827 | 1 | | | |
| | Nonreg. vs. Prosp. | 0.154 | [-1.82, 2.12] | 0.876 | 1 | | | |
| | Performance Based Function compared to Composite | | | | | | | |
| | Retrosp. vs. Prosp. | 0.353 | [-1.24, 1.95] | 0.659 | 1 | | | |
| | Nonreg. vs. Prosp. | -0.285 | [-2.02, 1.45] | 0.744 | 1 | | | |

| Exposure | Comparison | Interaction | 95% CI | P-value | Bonferroni-corrected P-value | Tau² | Tau² impact (% reduction of between study variance) | Studies excluded (missing variables) |
|---|---|---|---|---|---|---|---|---|
| Baseline measurement timing | Before surgery compared to after | | | | | | | |
| | Retrosp. vs. Prosp. | 0.033 | [-1.27, 1.34] | 0.959 | 1 | 0.511 | 19.6% | (1,6–8) |
| | Nonreg. vs. Prosp. | 0.051 | [-1.18, 1.28] | 0.934 | 1 | | | |
| Time to follow-up | 100 days longer | | | | | | | |
| | Retrosp. vs. Prosp. | 0.196 | [-0.06, 0.45] | 0.135 | 1 | 0.457 | 28.1% | (1,3) |
| | Nonreg. vs. Prosp. | -0.060 | [-0.21, 0.09] | 0.416 | 1 | | | |

*Meta-regression interaction analysis. Raw $Tau^2 = 0.6258$.*

### References for studies excluded from analysis (missing data):


1. Warner S, Ahmad A, Afzal MW, Khan S, Aslam MM, Gillani SA. Comparison of routine physical therapy exercises with and without core stability exercises in total knee replacement patients. Rawal Medical Journal. 2020 Dec 17;45(4):842–842.

2. Akbaba YA, Yeldan I, Guney N, Ozdincler AR. Intensive supervision of rehabilitation programme improves balance and functionality in the short term after bilateral total knee arthroplasty. Knee Surg Sports Traumatol Arthrosc. 2016 Jan;24(1):26–33.

3. Chow TP, Ng GY. Active, passive and proprioceptive neuromuscular facilitation stretching are comparable in improving the knee flexion range in people with total knee replacement: a randomized controlled trial. Clin Rehabil. 2010 Oct;24(10):911–8.

4. Codine Ph, Dellemme Y, Denis-Laroque F, Herisson Ch. The use of low velocity submaximal eccentric contractions of the hamstring for recovery of full extension after total knee replacement: A randomized controlled study. Isokinetics and Exercise Science. 2004 Aug;12(3):215–8.

5. Rajan R, Pack Y, Jackson H, Gillies C, Asirvatham R. No need for outpatient physiotherapy following total knee arthroplastyA randomized trial of 120 patients. Acta Orthopaedica Scandinavica. 2004 Jan;75(1):71–3.

6. Han ASY, Nairn L, Harmer AR, Crosbie J, March L, Parker D, et al. Early Rehabilitation After Total Knee Replacement Surgery: A Multicenter, Noninferiority, Randomized Clinical Trial Comparing a Home Exercise Program With Usual Outpatient Care. Arthritis Care & Research. 2015 Feb;67(2):196–202.

7. Lee HG, An J, Lee BH. The Effect of Progressive Dynamic Balance Training on Physical Function, The Ability to Balance and Quality of Life Among Elderly Women Who Underwent a Total Knee Arthroplasty: A Double-Blind Randomized Control Trial. IJERPH. 2021 Mar 3;18(5):2513.

8. Liebs TR, Herzberg W, Rüther W, Haasters J, Russlies M, Hassenpflug J. Ergometer Cycling After Hip or Knee Replacement Surgery: A Randomized Controlled Trial. The Journal of Bone and Joint Surgery-American Volume. 2010 Apr;92(4):814–22.
