## Appendix 8 for "Registration and reporting characteristics of trials investigating exercise therapy following total knee arthroplasty: A systematic review"

**Sensitivity analysis – Extended forest plot – trials recruiting before July 1st 2005 omitted.**

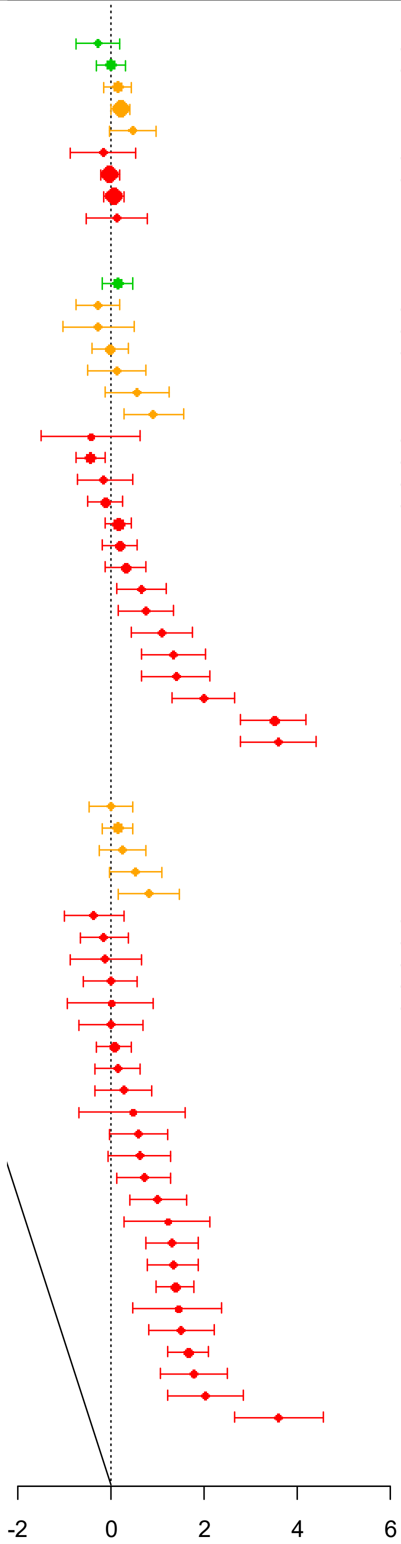


| Registration | Outcome hierarchy level | Estimate (95% CI) |
|---|---|---|
| **Prospective** | | |
| Larsen (2024) | 1 | -0.30 (-0.77 : 0.2) |
| Buhagiar (2017) | 1 | -0.01 (-0.31 : 0.3) |
| Lenguerrand (2020) | 1 | 0.14 (-0.16 : 0.4) |
| Hamilton (2020) | 1 | 0.20 (-0.01 : 0.4) |
| Sattler (2019) | 2 | 0.47 (-0.04 : 1.0) |
| Cheng (2022) | 2 | -0.17 (-0.87 : 0.5) |
| Fransen (2017) | 1 | -0.03 (-0.24 : 0.2) |
| Han (2015) | 1 | 0.05 (-0.15 : 0.3) |
| Do and Yim (2020) | 6 | 0.12 (-0.53 : 0.8) |
| **Retrospective** | | |
| Bade (2017) | 1 | 0.14 (-0.18 : 0.4) |
| Jakobsen (2014) | 1 | -0.29 (-0.75 : 0.2) |
| Christiansen (2015) | 1 | -0.28 (-1.05 : 0.5) |
| Harmer (2009) | 4 | -0.03 (-0.42 : 0.4) |
| Kelly (2015) | 1 | 0.11 (-0.52 : 0.7) |
| Alsayani (2024) | 6 | 0.55 (-0.14 : 1.2) |
| Carozzo (2022) | 3 | 0.91 ( 0.27 : 1.6) |
| Trudelle-Jackson (2020) | 3 | -0.46 (-1.52 : 0.6) |
| Ko (2013) | 2 | -0.45 (-0.75 : -0.1) |
| Tanaka (2019) | 6 | -0.15 (-0.74 : 0.4) |
| Xu (2021) | 6 | -0.12 (-0.50 : 0.2) |
| Moffet (2015) | 1 | 0.15 (-0.14 : 0.4) |
| Heikkilä (2017) | 4 | 0.18 (-0.20 : 0.6) |
| Maeda (2024) | 6 | 0.31 (-0.14 : 0.8) |
| Bruun-Olsen (2013) | 1 | 0.64 ( 0.11 : 1.2) |
| Jacksteit (2021) | 2 | 0.75 ( 0.15 : 1.4) |
| An (2023) | 6 | 1.10 ( 0.44 : 1.8) |
| Lee (2021) | 6 | 1.34 ( 0.65 : 2.0) |
| Husby (2018) | 2 | 1.40 ( 0.66 : 2.1) |
| Moutzouri (2018) | 2 | 1.99 ( 1.32 : 2.6) |
| Jiao (2023) | 2 | 3.50 ( 2.79 : 4.2) |
| Cetinkaya (2022) | 4 | 3.60 ( 2.78 : 4.4) |
| **Not registered** | | |
| Chow (2010) | 6 | 0.00 (-0.47 : 0.5) |
| den Hertog (2012) | 2 | 0.15 (-0.18 : 0.5) |
| Zietek (2015) | 6 | 0.24 (-0.25 : 0.7) |
| Schulz (2018) | 6 | 0.52 (-0.03 : 1.1) |
| Mau-Moeller (2014) | 2 | 0.81 ( 0.16 : 1.5) |
| Can (2023) | 6 | -0.37 (-1.01 : 0.3) |
| Levine (2013) | 5 | -0.15 (-0.67 : 0.4) |
| Yang (2013) | 4 | -0.12 (-0.90 : 0.6) |
| Dujin Park (2012) | 4 | -0.02 (-0.60 : 0.6) |
| Johnson (2010) | 4 | -0.01 (-0.94 : 0.9) |
| Pastore (2015) | 6 | 0.00 (-0.69 : 0.7) |
| Eymir (2021) | 3 | 0.06 (-0.31 : 0.4) |
| Shabbir (2017) | 2 | 0.14 (-0.35 : 0.6) |
| Tousignant (2011) | 6 | 0.26 (-0.34 : 0.9) |
| Park (2024) | 6 | 0.45 (-0.69 : 1.6) |
| Tsukada (2020) | 6 | 0.59 (-0.03 : 1.2) |
| Suh (2017) | 4 | 0.62 (-0.06 : 1.3) |
| Hardt (2018) | 3 | 0.70 ( 0.12 : 1.3) |
| Hepperger (2016) | 6 | 1.00 ( 0.40 : 1.6) |
| Teissier (2020) | 6 | 1.21 ( 0.29 : 2.1) |
| Bily (2016) | 4 | 1.30 ( 0.73 : 1.9) |
| Unver (2016) | 4 | 1.34 ( 0.78 : 1.9) |
| Liao (2013) | 5 | 1.36 ( 0.95 : 1.8) |
| Papotto (2012) | 3 | 1.42 ( 0.47 : 2.4) |
| Akbaba (2014) | 4 | 1.51 ( 0.82 : 2.2) |
| Li (2019) | 6 | 1.65 ( 1.21 : 2.1) |
| Molla (2017) | 4 | 1.77 ( 1.05 : 2.5) |
| Karaman (2017) | 3 | 2.04 ( 1.23 : 2.9) |
| Warner (2020) | 2 | 3.61 ( 2.66 : 4.6) |

### Sensitivity analysis – Funnel plot – trials recruiting before July 1st 2005 omitted.

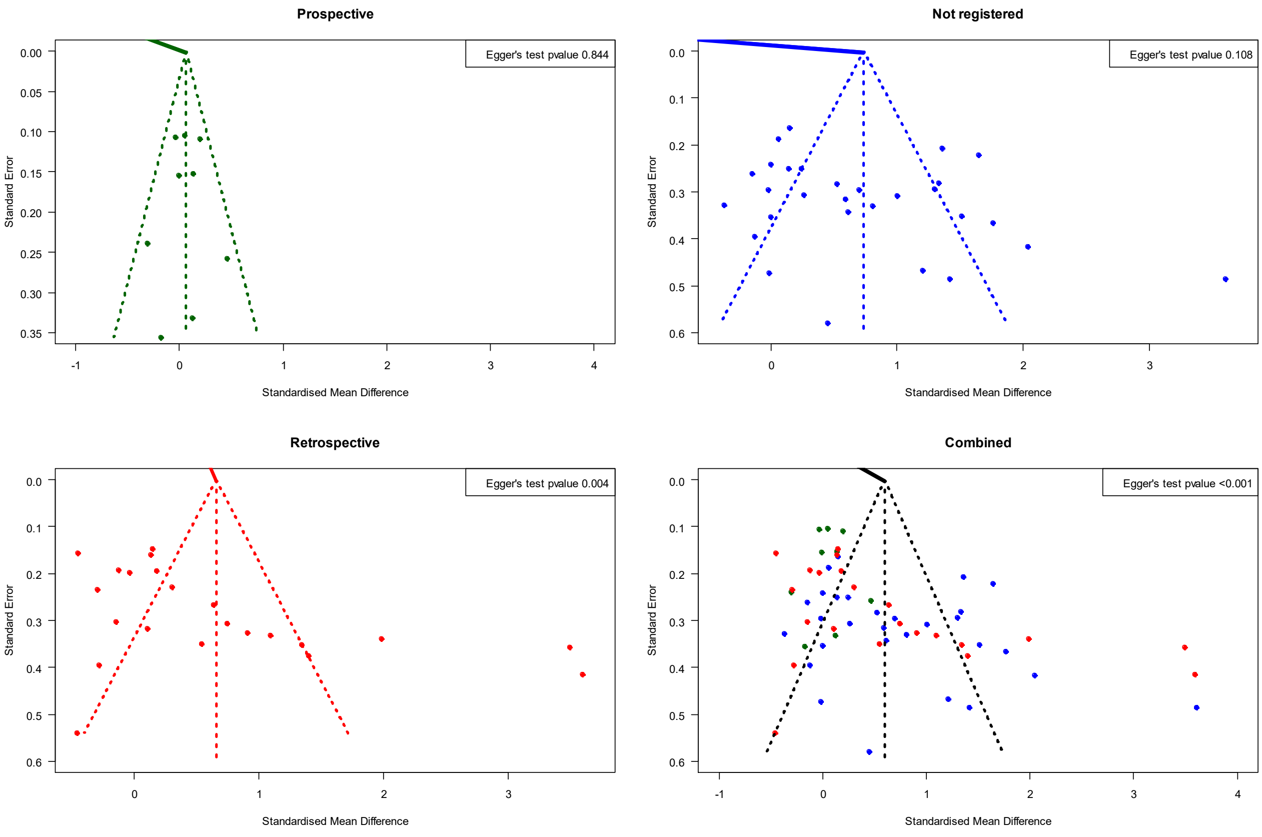


### Sensitivity analysis – Combined forest plot – trials recruiting before July 1st 2005 omitted.

| Registration | Number of studies | Number of participants | $I^2$ | Estimate (95% CI) |
|---|---|---|---|---|
| Prospective | 9 | 1568 | 0.01 | 0.06 (-0.03 : 0.16) |
| Retrospective | 22 | 1549 | 0.91 | 0.66 ( 0.16 : 1.15) |
| Not registered | 29 | 1498 | 0.84 | 0.74 ( 0.42 : 1.05) |
| Total | 60 | 4615 | 0.88 | 0.60 ( 0.36 : 0.83) |

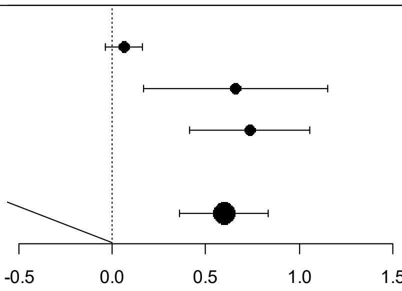
